# The implantable system that restores hemodynamic stability after spinal cord injury

**DOI:** 10.1101/2024.05.10.24306826

**Authors:** Aaron A. Phillips, Aasta P. Gandhi, Nicolas Hankov, Sergio D. Hernandez-Charpak, Julien Rimok, Anthony Incognito, Anouk E. J. Nijland, Marina D’Ercole, Anne Watrin, Maxime Berney, Aikaterini Damianaki, Grégory Dumont, Nicolò Macellari, Laura De Herde, Nadine Intering, Donovan Smith, Ryan Miller, Meagan N. Smith, Jordan Lee, Edeny Baaklini, Jean-Baptiste Ledoux, Javier G. Ordonnez, Taylor Newton, Ettore F. Meliadò, Léa Duguet, Charlotte Jacquet, Léa Bole-Feysot, Markus Rieger, Kristen Gelenitis, Yoann Dumeny, Miroslav Caban, Damien Ganty, Edoardo Paoles, Thomas Baumgartner, Clinical Study Team, Onward Team, Cathal Harte, Charles David Sasportes, Paul Romo, Tristan Vouga, Jemina Fasola, Jimmy Ravier, Matthieu Gautier, Frédéric Merlos, Rik Buschman, Tomislav Milekovic, Andreas Rowald, Stefano Mandija, Cornelis A.T. van den Berg, Niels Kuster, Esra Neufeld, Etienne Pralong, Lorenz Hirt, Stefano Carda, Fabio Becce, Etienne Aleton, Kyle Rogan, Patrick Schoettker, Grégoire Wuerzner, Nelleke Langerak, Noël L.W. Keijsers, Brian K. Kwon, James D. Guest, Erika Ross, John Murphy, Erkan Kurt, Steve Casha, Fady Girgis, Ilse van Nes, Kelly A. Larkin-Kaiser, Robin Demesmaeker, Léonie Asboth, Jordan W. Squair, Jocelyne Bloch, Grégoire Courtine

## Abstract

A spinal cord injury (SCI) causes immediate and sustained hemodynamic instability that threatens neurological recovery and impacts quality of life. Here, we establish the clinical burden of chronic hypotensive complications due to SCI in 1,479 participants, and expose the ineffective treatment of these complications with conservative measures. To address this clinical burden, we developed a purpose-built implantable system based on biomimetic epidural electrical stimulation (EES) of the spinal cord that immediately triggered robust pressor responses. The system durably reduced the severity of hypotensive complications in people with SCI, removed the necessity for conservative treatments, improved quality of life, and enabled engagement in activities of daily living. Central to the development of this therapy was the head-to-head demonstration in the same participants that EES must target the last three thoracic segments, and not the lumbosacral segments, to achieve the safe and effective regulation of blood pressure in people with SCI. These findings in 14 participants establish a path for a pivotal device trial that evaluates the safety and efficacy of EES to treat the underappreciated, treatment-resistant hypotensive complications due to SCI.

## Introduction

A spinal cord injury (SCI) disrupts the communication between supraspinal vasomotor centers and the regions of the spinal cord that regulate blood pressure^1,2^. The consequence is pernicious exposure to disabling and potentially life-threatening blood pressure instability that threatens neurological recovery^3–8^. A prominent feature of this hemodynamic instability is a spectrum of hypotensive complications that include orthostatic hypotension, resting hypotension, postprandial hypotension, and exertional hypotension^2^. These complications are clinically managed with conservative measures such as uncomfortable abdominal binders, pressure stockings, high sodium diets, and slow-acting pharmacological pressor agents^2^. Despite these measures, the majority of people who present with hemodynamic instability due to SCI remain chronically hypotensive, which impacts their quality of life and reduces engagement in social and professional activities^2,9–12^. This symptomatology^9,10,13^, coupled with the unexplained prevalence of stroke and heart disease in this population, compelled health institutions to encourage the development of new approaches to manage hemodynamic instability in people with SCI^14,15^.

Recent case studies documented transient increases in blood pressure in response to epidural electrical stimulation (EES) applied over the lumbosacral spinal cord of people with SCI^16–21^. Since the lumbosacral spinal cord contains a paucity of sympathetic preganglionic neurons that are required for the regulation of blood pressure^1,22^, these serendipitous observations appeared to be incongruent with the anatomy of the autonomic nervous system^22,23^. Consequently, the mechanisms underlying these pressor responses remain enigmatic, casting doubt that observations of blood pressure elevations triggered by EES applied over the lumbosacral spinal cord could translate into a safe and efficacious treatment to manage hemodynamic instability in people with SCI.

To address the unsatisfying management of hemodynamic instability in people with SCI, we previously developed a preclinical model of hemodynamic instability due to SCI with the aim of studying the mechanisms through which EES applied over the spinal cord can lead to increases in blood pressure^1,24^. We found that EES depolarizes large-diameter afferents where they bend to enter the spinal cord through the dorsal root entry zones^1,25^. The recruitment of these afferents leads to the activation of splanchnic neurons via the activation of Vsx2-expressing neurons that directly connect to sympathetic preganglionic neurons^1,25^. Importantly, the amplitude of pressor responses followed a Gaussian distribution wherein the largest responses were elicited when EES was applied over the lower thoracic spinal cord. This distribution correlated with the density of sympathetic preganglionic neurons, in line with the known anatomy of the autonomic nervous system^22^. We called this region the *hemodynamic hotspot*. We translated this understanding into a neuromodulation strategy operating in closed-loop that adjusts EES amplitudes delivered over the *hemodynamic hotspot* to regulate blood pressure in real time. This strategy maintained blood pressure stability during transient, varying, and sustained hemodynamic challenges in rat and nonhuman primate models of SCI, and in one human participant^1^.

Here, we aimed to establish the burden of hypotension and assess the effectiveness of current treatments in large populations of people living with SCI in order to justify new treatments involving neurosurgical interventions. We then sought to address the ambiguity on the optimal location of the spinal cord to apply EES to treat hemodynamic instability and exploited this understanding to develop a purpose-built implanted system that harnesses the mechanisms through which EES regulates blood pressure. We finally provide preliminary validation on the long-term efficacy of this new system and resulting improvements in quality of life, paving the way towards pivotal clinical trials.

## Results

### Underappreciated prevalence of medically-refractory hypotensive complications

EES is a promising therapy to address the lack of satisfying management options for hemodynamic instability due to SCI. However, EES requires a neurosurgical intervention that must be weighed against the risks and benefits of the procedure. Establishing this balance requires an understanding of the prevalence, symptomatology, and effectiveness of current management strategies. While hypotension is a recognized medically refractory complication of SCI^2^, these epidemiological factors have not been quantified conclusively.

To address this knowledge gap, we analyzed the Spinal Cord Injury Community Survey (SCICS)^26,27^, which includes self-reported information on symptoms of orthostatic hypotension and demographic information in 1,479 individuals living with chronic SCI (**Supplementary Fig. 1a-c** and **supplementary Note 1**). This analysis revealed that 78% of individuals with tetraplegia had been told by a medical practitioner that they have orthostatic hypotension. Of this subset of individuals who were diagnosed, 28% of them were being treated for orthostatic hypotension, yet 91% still experienced symptoms (**Fig. 1a**).

**Fig. 1.**
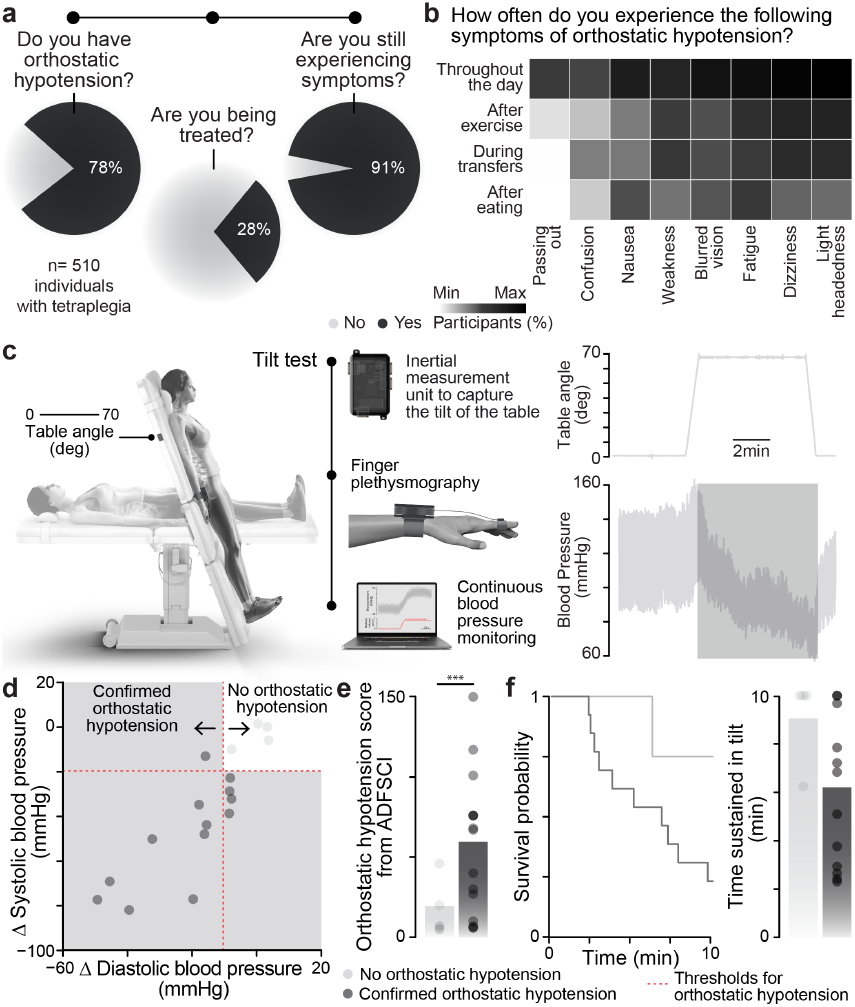
Orthostatic hypotension is a medically refractory condition in people with SCI. **a**, The prevalence of orthostatic hypotension and management efficacy in 1,479 individuals with tetraplegia (n= 510) from the Rick Hansen Spinal Cord Injury Registry^26,27^. **b**, Percentage of individuals with tetraplegia experiencing each symptom scored in the Autonomic Dysfunction Following Spinal Cord Injury survey (ADFSCI) across various daily activities (n = 107). **c**, Scheme of tilt-table test and representative blood pressure response. **d**, Drop in systolic and diastolic blood pressure during a tilt-table test (n = 17). **e**, ADFSCI total orthostatic hypotension score in individuals with and without clinically defined orthostatic hypotension (n = 4 with no orthostatic hypotension, n = 13 with confirmed orthostatic hypotension, independent samples two-tailed Welch’s t-test: t = 12.84; p-value = 2.37e-02). **f**, Kaplan-Meier plot of exposure status to time, segregated by the presence of clinically defined orthostatic hypotension. Bar graph shows the corresponding time to tilt end.

This analysis compelled us to characterize the pattern of hypotension-related symptoms due to SCI. For this purpose, we surveyed a diverse sample of 254 individuals with SCI, which included responses to the Autonomic Dysfunction Following Spinal Cord Injury (ADFSCI) rating scale. When considering individuals with tetraplegia, we found that every individual who responded to the survey experienced symptoms of hypotension throughout the day. These symptoms were primarily characterized by lightheadedness, dizziness, fatigue, blurred vision, and weakness throughout the day (**Fig. 1b**).

In a subset of these individuals, we found that participants who met the criteria for orthostatic hypotension during the tilt-table test reported significantly more hypotensive symptoms (**Fig. 1c-f, Supplementary Fig. 2a-i** and **Supplementary Table 3**).

These results established that i) the vast majority of individuals living with tetraplegia experience severe and persistent symptoms of hypotension; ii) hypotension is accompanied by quantifiable symptomatology that increases with the level and completeness of injury, and iii) people with SCI continue experiencing symptoms of orthostatic hypotension despite medical management.

We concluded that these outcomes, combined with the long-term consequences of medically refractory hypotension, justified the consideration of therapeutic strategies involving surgical interventions to manage hypotension due to SCI.

### EES is more effective when applied over lower thoracic than lumbosacral segments

Extensive preclinical evidence in mice, rats, and nonhuman primates as well as clinical case studies demonstrated that EES reduces the severity of orthostatic hypotension^1,16–19,28^. However, the optimal location to deliver EES has remained controversial. We previously showed that EES activates large-diameter afferents where they bend to enter the spinal cord through the dorsal root entry zones and that this recruitment engages sympathetic preganglionic neurons indirectly via Vsx2-expressing excitatory neurons nested in the intermediate lamina of the stimulated region of the spinal cord^1,25^ (**Fig. 2a**). We also confirmed an enrichment of these sympathetic preganglionic neurons in the lower thoracic spinal cord (**Fig. 2a**), and showed that this enrichment coincided with the maximal pressor response to EES. This location was thus termed the *hemodynamic hotspot*^1^. Despite this evidence, many studies reported unexpected pressor responses during the application of EES over the lumbosacral spinal cord^16–19^. The incongruence of these observations with the anatomy of the autonomic nervous system^22,23^ compelled us to perform a head-to-head comparison in the same participants to establish the optimal location to deliver EES for the regulation of blood pressure in humans with SCI.

**Fig. 2.**
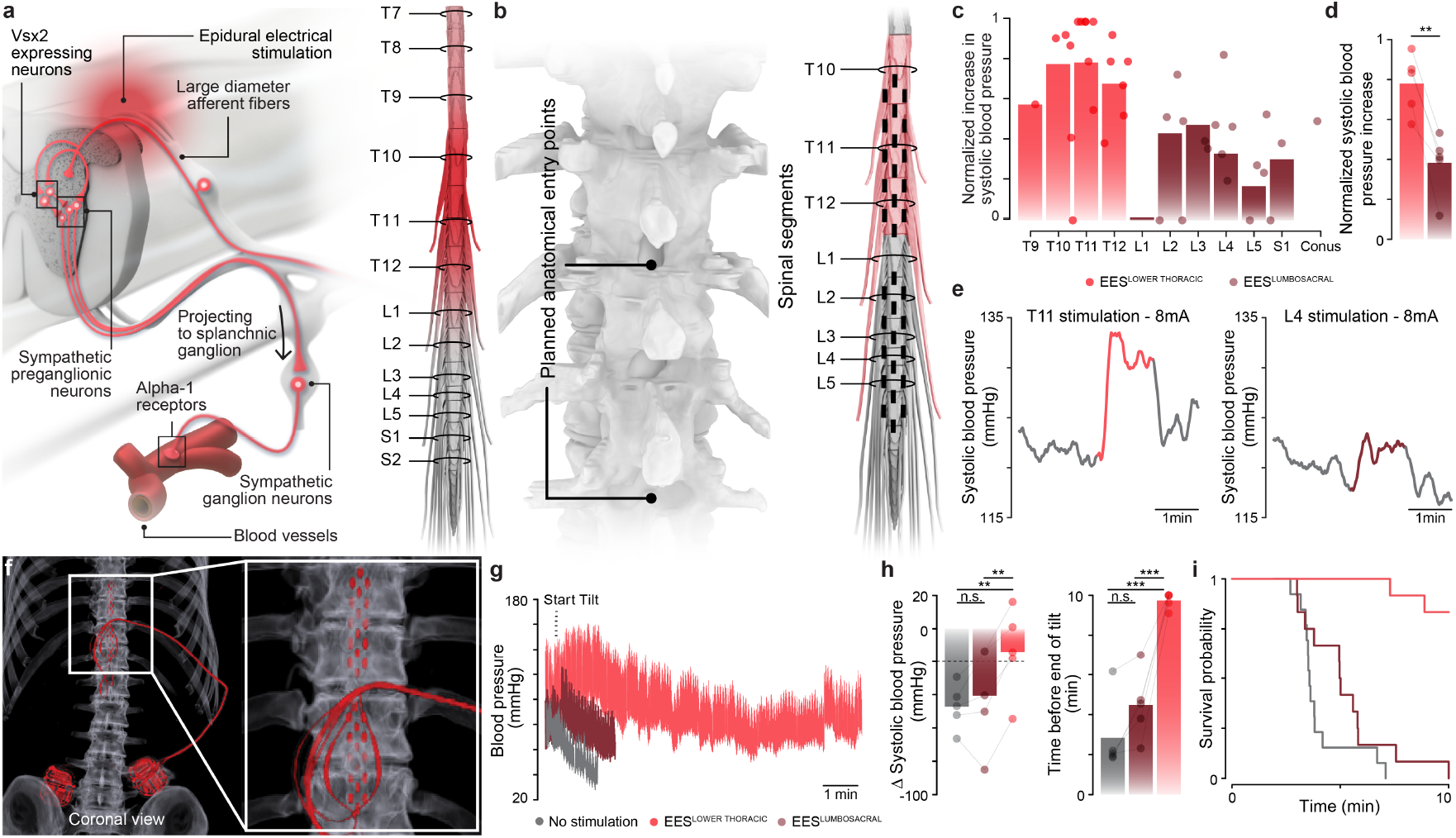
Validation of the location of the *hemodynamic hotspot*. **a**, Overview of the neuronal architecture recruited EES to trigger pressor responses. **b**, Anatomical planning of laminotomies for inserting the paddle lead (*left*) based on the targeted location of the paddle leads (*right*) over the thoracic and lumbosacral segments. **c**, Amplitude of systolic pressor response per spinal segment when delivering EES intraoperatively. Dots (n = 5 participants) denote pressure responses to EES applied over specific segments, the location of which was reconstructed postoperatively. **d**, Bar graph shows averaged changes in systolic blood pressure in response to EES applied over thoracic or lumbar spinal segments (n = 5, paired samples two-tailed t-test; t = 6.46; p = 3.00e-03). **e**, Systolic blood pressure response measured intraoperatively while EES was applied over the T11 versus L4 spinal segments for a representative participant. **f**, Sagittal and coronal reconstructions from a postoperative computed tomography scan. **g**, Changes in blood pressure during a tilt-table test without EES, and with EES applied over either the lower thoracic or lumbosacral spinal segments. **h**, Drop in systolic blood pressure during a 10-minute tilt-table test, and corresponding tilt duration (n = 5, repeated measures ANOVA with Tukey’s HSD). **i**, Kaplan-Meier plot of exposure status to time until end of tilt, segregated by the location over which EES was applied (n = 37 tilts; mixed model Cox regression with likelihood ratio test estimate = 36.2; p-value = 2.48e-06).

Consequently, we initiated two clinical studies in Canada and Switzerland to compare the effect of EES applied over the lower thoracic versus lumbosacral spinal cord on hemodynamic instability (ClinicalTrials.gov ID: NCT04994886, NCT05044923 We enrolled 6 individuals with medically-refractory orthostatic hypotension secondary to their SCI (**Supplementary Fig. 3a** and **Supplementary Table 2**). The six participants included in these clinical study met the established clinical criteria for orthostatic hypotension since none of these participants were able to sustain the orthostatic challenge imposed by verticalization on a tilt table test, showing an average drop of 50 mmHg in systolic blood pressure within 3 minutes (**Supplementary Fig. 3b-f**). This drop far exceeds the international criteria for orthostatic hypotension, set at a 20 mmHg drop of systolic blood pressure within 3 minutes^29^.

Since only repurposed technologies originally designed to alleviate neuropathic pain were available to target both locations (**Fig. 2b** and **Supplementary Fig. 3g**), we sought to optimize the surgical implantation of two separate paddle leads over the lower thoracic spinal cord and the lumbosacral spinal cord. To facilitate this placement, we generated computational models that we personalized with computed tomography and magnetic resonance imaging sequences that we previously optimized to enable the identification of the dorsal root entry zones targeted by EES^30^ (**Fig. 2b** and **Supplementary Fig. 4**). We then injected these coordinates into our preoperative planning software that we consolidated with intraoperative imaging of the spinal column. Under general anesthesia, we performed laminotomies to enable the insertion of both leads over the planned anatomical locations. We then fine-tuned the final position of both leads based on the muscle responses to EES that are expected from the known rostrocaudal distribution of motor neuron pools innervating trunk and leg muscles^30^ (**Supplementary Fig. 3h-j**).

Once the location of both leads was secured, we applied EES (120 Hz, 1-10 mA, 300 µs) over each pair of electrodes on each row of both paddles to assess pressor responses that we monitored by measuring beat-by-beat blood pressure through an arterial line. We mapped pressor responses to post-operative reconstructions of the spinal cord regions activated by each pair of electrodes. As previously established in rat and nonhuman primate models of SCI^1^, we found that pressor responses followed a Gaussian distribution, wherein the peak pressor responses were centered over the penultimate segment of the thoracic spinal cord (**Fig. 2c-e** and **Supplementary Fig. 3k-l**).

Postoperative computed tomography acquisitions confirmed that both leads were positioned over the planned locations (**Fig. 2f**). Intraoperative assessments confirmed that the optimal location to elicit pressor responses with EES was centered over the lower thoracic spinal cord—the *hemodynamic hotspot*. Therefore, we next verified whether EES applied over the *hemodynamic hotspot* was superior to EES applied over the lumbosacral spinal cord to stabilize blood pressure during formal tilt-table testing post-operatively, since previous case studies^16–19^ reported a mitigation of orthostatic hypotension when EES was applied over the lumbosacral segments (**Fig. 2g-i** and **Supplementary Fig. 5a-g**).

We conducted comprehensive mapping sessions to identify optimal configurations of electrodes and stimulation parameters that could trigger pressor responses when EES was delivered over the lumbosacral segments (**Supplementary Fig. 5a-d**). Despite this mapping, we failed to identify parameters of EES that could trigger robust and reproducible pressor responses without eliciting spasms in leg muscles. EES applied over the lumbosacral segments marginally reduced the severity of orthostatic hypotension (**Fig. 2g-i** and **Supplementary Fig. 5e-g**).

This failure contrasted with the robust pressor responses elicited when EES was applied over the lower thoracic spinal cord, which enabled all the participants to improve their tolerance to orthostatic challenges induced by the verticalization during tilt-table tests in the absence of undesirable sensations (**Fig. 2g-i** and **Supplementary Fig. 5a-g**).

These results demonstrate that EES applied over the lumbosacral spinal cord failed to consistently trigger meaningful pressor responses, as expected based on the incongruence between this location and the anatomy and physiology of the autonomic nervous system. Instead, EES applied over the *hemodynamic hotspot*, wherein an enrichment of sympathetic preganglionic neurons involved in the modulation of blood pressure reside, consistently triggered robust pressor responses that enabled all six participants to tolerate severe orthostatic challenges.

These quantifications provide important results for the ongoing debated optimal location of EES for effective treatment of hemodynamic instability in people with SCI. In addition, recent preclinical experiments demonstrated that longterm application of EES over the lumbosacral spinal cord augmented the density of maladaptive synaptic connections from Vsx2-expressing neurons of the lumbosacral to Vsx2-expressing neurons of the lower thoracic spinal cord, which reinforces the severity of autonomic dysreflexia^25^. Given the discrepancy in safety and efficacy of EES applied over the *hemodynamic hotspot* versus lumbosacral segments, these results establish a safe path forward on how to deliver EES to treat hemodynamic instability due to SCI.

### Purpose-built neurostimulation platform to restore hemodynamic stability after SCI

We aimed to develop a clinical-grade implantable neurostimulation platform that addresses the limitations of existing systems and meets the identified requirements for the safe and effective management of hemodynamic instability in people with SCI (**Supplementary Note 2**).

Since the possibility to adjust EES parameters in closed-loop was a desired feature of the platform, we designed an implantable neurostimulation platform that continuously communicates with a wearable hub through near field magnetic induction (NFMI) (**Fig. 3a**). This hub can acquire external signals, including continuous measurements of blood pressure, and embeds computing power to support closed-loop control algorithms. This configuration enables closed-loop control of EES parameters with latencies as low as 25 ms. We programmed the firmware to support any configuration of anodes and cathodes, and to resolve the limited range of programmable EES parameters in current neurostimulation platforms.

**Fig. 3.**
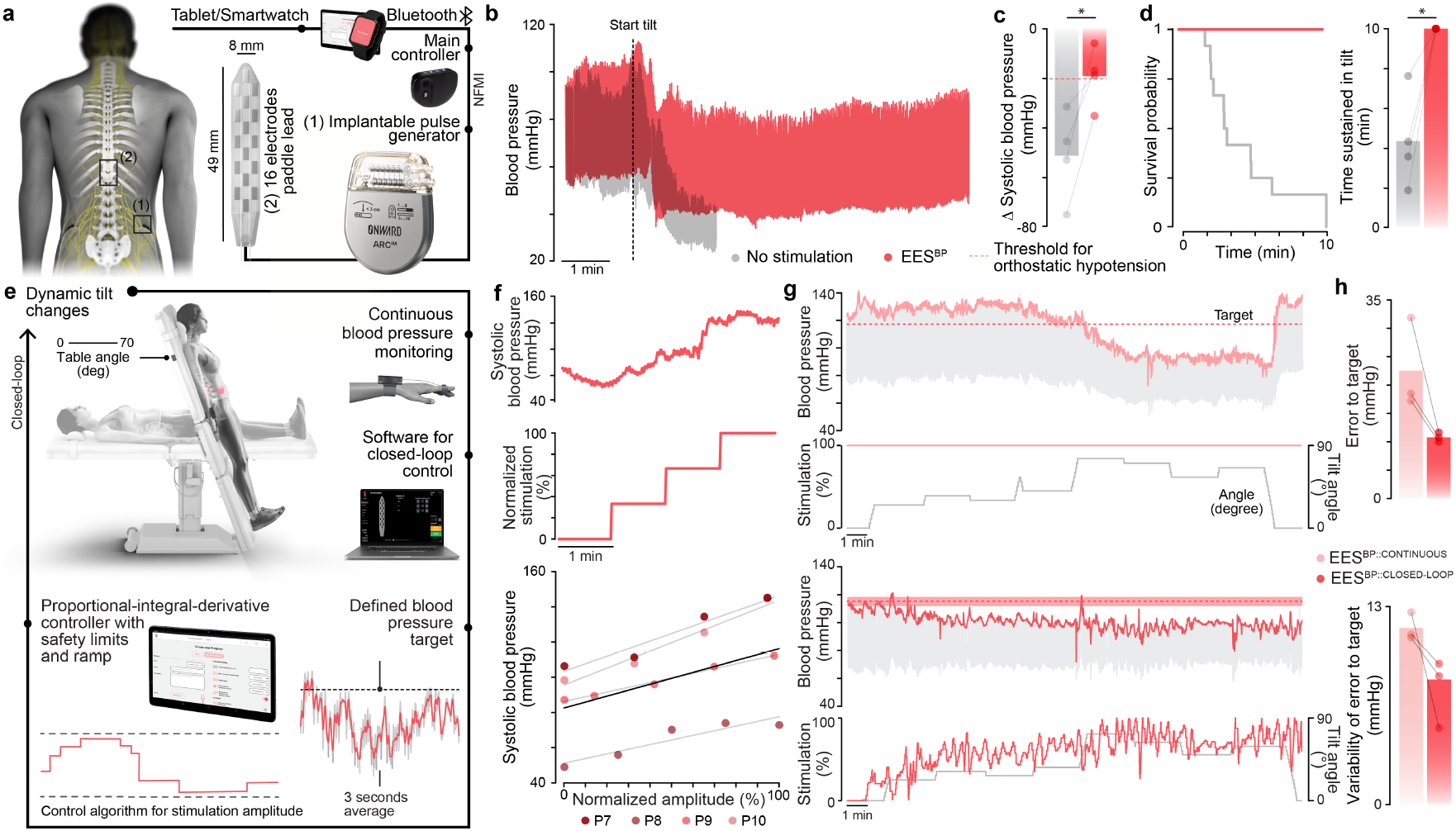
Validation of the new, purpose-built implantable neurostimulation platform. **a**, Scheme of the investigational system, including a clinician controller (tablet), patient programmer (external smartwatch), a main controller (hub), implantable pulse generator (IPG), and a Medtronic 5-6-5 Surescan Specify paddle lead. **b**, Changes in blood pressure from a representative participant during an orthostatic challenge without and with EES applied over the hemodynamic hotspot (EES^BP^). **c**, Average drop in systolic blood pressure during orthostatic challenge without and with EES (n = 4, paired samples two-tailed t-test; t = 4.27; p = 2.4e-02). **d**, Kaplan-Meier plot of exposure status to time, segregated by the presence or absence of EES (n = 24 tilts; mixed model Cox regression with likelihood ratio test estimate = 15.36; p-value = 4.00e-04) and tilt duration (n = 4, paired samples two-tailed t-test; t = 4.69; p = 1.8e-02) without EES and with EES applied over the hemodynamic hotspot. **e**, Closed-loop control of blood pressure consisting of a proportional-integral-derivative (PID) controller that adjusts the amplitude of EES applied over the hemodynamic hotspot to maintain blood pressure within a range of predefined blood pressure targets. **f**, Stepwise increase in systolic blood pressure in response to gradual increases of EES amplitude (1 mA per min) for a representative participant. Relationship between EES amplitude and systolic blood pressure (n = 4; black line represents model fit of a mixed effects linear regression R^2^ = 0.97, p-value = 8.83e-09). **g**, Changes in systolic blood pressure during a dynamic orthostatic challenge while EES is delivered continuously (top) or controlled in closed-loop (*bottom*) to maintain the systolic blood pressure within a target range. **h**, Error between the predefined blood pressure target (*top*) and variability of error to target (*bottom*) when EES was delivered continuously or controlled in closed-loop (n = 3/4 - Trial 3, 1 participant did not complete assessment due to spasticity).

The hub communicates wirelessly with a touch-screen tablet and a smartwatch, from which therapeutic programs can be selected and parameterized (**Fig. 3a**). We optimized these user interfaces to comply with usability assessments and user needs. We also endowed the user interface with practical features that reduce the impact of impaired arm and hand functions for operating the therapy independently and safely. In addition, customizable algorithms enable real time adjustment of set-point amplitudes based on hemodynamic requirements that are well known to differ throughout the day, such as after meals and during sleep and exercise^2,31–33^.

After meeting the requirements for all applicable standards for active medical device development, we tested the preliminary safety and efficacy of this new platform under the protocol of a new clinical study conducted in Switzerland (ClinicalTrials.gov ID: NCT05111093) (**Supplementary Fig. 6a**). We enrolled 4 new participants who presented with debilitating medically-refractory hypotension (**Supplementary Fig. 6b-f** and **Supplementary Table 2**) and implanted the purpose-built neurostimulation platform, but maintained every other aspect of our preoperative and operative procedures, including the implantation of the same repurposed paddle lead over the *hemodynamic hotspot* as in our previous experiments (**Supplementary Fig. 6g-h**).

In all the participants, the implanted neurostimulation platform met the requirements to deliver EES waveforms that elicit robust pressor responses (**Supplementary Fig. 6i-l**) that enabled all the participants to improve their tolerance to orthostatic challenges (**Fig. 3b-d** and **Supplementary Fig. 6m-s**). As we previously reported in rats and nonhuman primates^1^, we confirmed in all four participants that the magnitude of pressor responses scaled linearly with increasing amplitudes of EES (**Fig. 3f** and **Supplementary Fig. 7a**). This linear relationship suggested that blood pressure could be regulated in real time based on a proportional-integral-differential controller regulating EES amplitudes to maintain blood pressure within user-defined levels.

To test this possibility, we leveraged the closed-loop capabilities and versatile software environment of the new neurostimulation platform to implement this proportional-integral-differential controller (**Fig. 3e**). We then evaluated the performance of this controller during formal and dynamic tilt-tests. As expected based on previous experiments (**Fig. 2g-i**), we found that the delivery of continuous EES decreased the severity of orthostatic hypotension during formal tilt-tests wherein the amplitude of EES remained constant (**Fig. 3b-d**). However, dynamic tilt-testing sessions exposed the limitations of continuous EES, since pseudorandom changes in body orientation with respect to the direction of gravity led to dynamic perturbations of blood pressure that could not be managed satisfactorily with continuous EES. In contrast, closed-loop control of EES led to rapid and precise updates to EES amplitudes that translated into finely-tuned levels of blood pressure during dynamic tilt-tests (**Fig. 3g** and **Supplementary Fig. 7b**). Closed-loop control of EES applied over the *hemodynamic hotspot* maintained blood pressure levels within the desired target range for extended periods of time despite transient and dynamic changes in postural orientation (**Fig. 3g, Supplementary Fig. 7b** and **Supplementary Fig. 11n**). Quantifications also revealed the superiority of EES controlled in closed-loop over both continuous EES and alternative closed-loop approaches relying on inertial measurement units or manual adjustments of EES (**Fig. 3h, Supplementary Fig. 7c** and **Supplementary Fig.11o**).

These results confirmed that the newly developed neurostimulation platform fulfilled the technical requirements to modulate blood pressure with EES, and established the possibility of achieving precise control of blood pressure in closed-loop despite transient, varying, and sustained hemodynamic challenges in people with orthostatic hypotension due to SCI.

### A new paddle lead targets the *hemodynamic hotspot*

We next aimed to develop a clinical-grade paddle lead that combines the necessary features to target the large-diameter afferents projecting to the *hemodynamic hotspot* while avoiding the undesired side effects of non-selective stimulation.

Our mapping experiments showed that the *hemodynamic hotspot* is centered over the last three segments of the lower thoracic spinal cord. Moreover, the same experiments revealed that EES applied over multiple segments leads to cumulative increases in the amplitude of pressor responses compared to when the same amplitude of EES is applied over a single spinal segment (**Fig. 4a**).

**Fig. 4.**
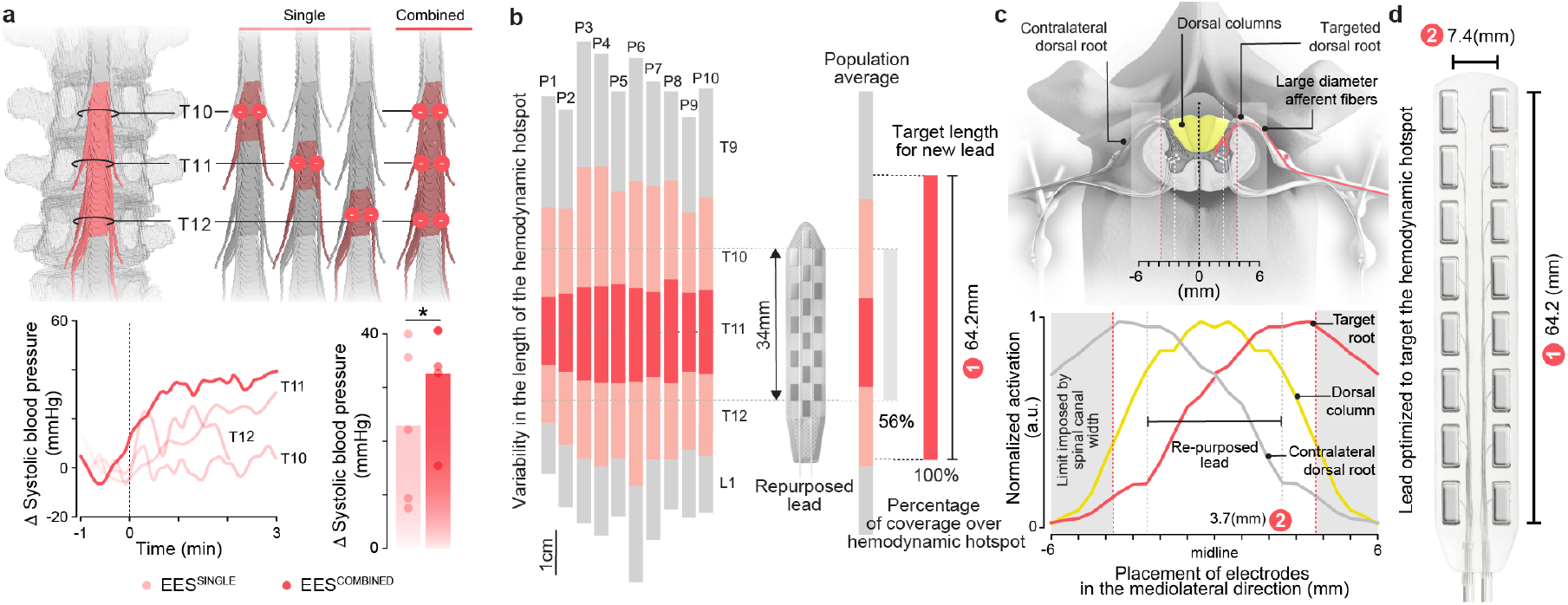
Purpose-built paddle lead to target the hemodynamic hotspot. **a**, EES applied over multiple segments of the hemodynamic hotspot leads to superior increase in blood pressure compared to EES applied over the median of a single segment (paired samples, one-tailed t-test, n = 5, t = 2.69, p = 2.72e-02). **b**, Quantification of the length of the lower thoracic spinal cord in the study participants (n=10) (*left*), and average length of the lower thoracic spinal cord quantified from published data (*right*). The coverage of the hemodynamic hotspot by the Medtronic 5-6-5 lead and expected from the newly-designed paddle lead are reported. **c**, Illustrative drawing of the anatomical model implemented in-silico to predict the relationships between the lateral position of EES electrodes and the relative recruitment of the targeted dorsal root, contralateral dorsal root, and dorsal columns, as shown in the plot. **d**, Photograph of the newly-designed purpose-built paddle lead that combines 2 parallel columns of 8 equally spaced electrodes that aim to cover the entire length of the hemodynamic hotspot while avoiding the recruitment of the dorsal columns.

We thus reasoned that the optimal paddle lead to regulate blood pressure with EES must maximize the coverage of the last three thoracic segments of the spinal cord. However, our analysis of the 10 individuals who were implanted with the repurposed paddle revealed that the coverage of these segments only reached 56% over the targeted region (**Fig. 4b** and **Supplementary Fig. 8a**). Moreover, 6 of the 16 electrodes were located over the midline of the spinal cord. These electrodes elicited undesired muscle spasms in the legs through antidromic volleys elicited along sensory axons ascending the dorsal columns (**Supplementary Fig. 8b**). This analysis emphasized the necessity to design a new paddle lead with a configuration of electrodes that meets the requirements to modulate blood pressure with EES.

To guide the informed design of this lead, we leveraged our digital library of human thoracic spinal cords that were reconstructed from magnetic resonance imaging acquisitions and combined these reconstructions with anatomical values from the literature^30,34–39^. These anatomical quantifications uncovered the requirements to arrange two columns of equally distributed electrodes that could cover 100% of the last three segments of the human thoracic spinal cord in more than 95% of the population (**Fig. 4b**). In addition, we conducted computer simulations to identify the optimal mediolateral location of electrodes that would target the dorsal root entry zones innervating the lower thoracic spinal cord while avoiding the undesired recruitment of the sensory axons in the dorsal columns. These simulations combined with quantifications from our anatomical models of the spinal cord revealed that the optimal tradeoff for the placement of these electrodes within the anatomical constraints of the spinal column was 3.7 mm lateral to the dorsal root entry zones (**Fig. 4c** and **Supplementary Fig. 8c-d**). We embedded the identified configuration of electrodes into the design of a new paddle lead that we fabricated with conventional medical-grade technologies and validated for biocompatibility (**Fig. 4d**).

### The implanted system that regulates hemodynamics after SCI

Having developed the complete implantable system with the required features to regulate blood pressure with EES, we amended our existing clinical study (ClinicalTrials.gov ID: NCT05111093) to interface the new paddle lead with the purpose-built neurostimulation platform (**Supplementary Fig. 9a**).

We enrolled 3 new participants who presented with debilitating hypotension (**Supplementary Fig. 9b-f** and **Supplementary Table 2**) and implanted them with the complete new system following the same procedure as described above. The extended length of the lead and optimal coverage of the dorsal root entry zones innervating the *hemodynamic hotspot* simplified the preoperative planning and intraoperative placement of the lead (**Supplementary Fig. 9g-h**). Indeed, the extended length avoided compromises on whether the rostral or caudal regions of the *hemodynamic hotspot* had to be targeted with shorter leads, while the optimized mediolateral electrode locations ensured the selective recruitment of the left versus right dorsal root entry zones (**Fig. 4c** and **Supplementary Fig. 8c-d**). These improved features of the paddle lead minimized the need for intraoperative fine-tuning of the paddle lead position. Updates to the personalized computational models of the spinal cord with postoperative imaging data confirmed that the lead covered the entire extent of the *hemodynamic hotspot* and predicted selective activation of the dorsal root entry zones in all participants (**Fig. 5b**).

**Fig. 5.**
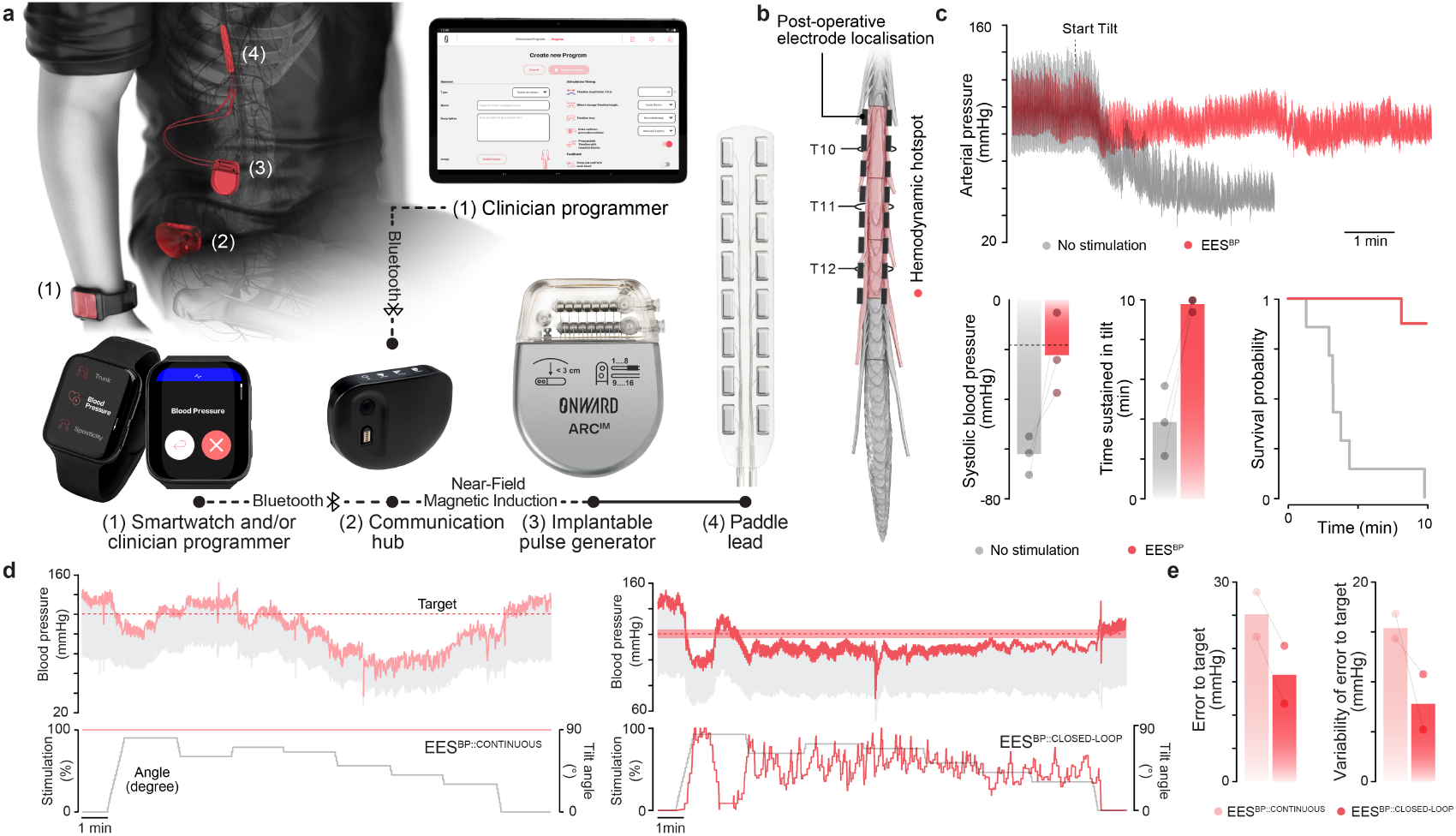
Complete purpose-built system to regulate blood pressure. **a**, Description of the complete system, including the paddle lead, implantable pulse generator, communication hub, and external smartwatch to operate the various programs of the therapy. **b**, Postoperative reconstruction of the final position of the paddle lead. **c**, Changes in blood pressure from a representative participant during an orthostatic challenge without EES and with continuous EES applied over the hemodynamic hotspot. The bar plots report the average drop in systolic blood pressure during orthostatic challenge and average tilt duration without EES and with EES applied over the hemodynamic hotspot. Kaplan-Meier plot of exposure status to time, segregated by the presence or absence of EES. **d**, Changes in diastolic and systolic blood pressure during a dynamic orthostatic challenge while EES is applied continuously (*left*) or in closed-loop (*right*) to maintain the systolic blood pressure within a target range (red shaded area) in P11. **e**, Error between the predefined blood pressure target and variability of error to target when EES was delivered continuously or controlled in closed-loop (n = 2/3 - Trial 3, 1 participant did not complete the assessment due to spasticity).

The optimized location of the electrodes also eliminated the need for extensive and cumbersome mapping sessions to identify the optimal configurations and parameters to elicit pressor responses^40^. Instead, minimal adjustments of predetermined electrode configurations (three pairs of electrodes covering the *hemodynamic hotspot*) and parameters (120 Hz frequency, 0.3 ms pulse duration, up to 20 mA amplitude) were sufficient to trigger robust, safe, and well-controlled increases in blood pressure during the first postoperative stimulation session (**Supplementary Fig. 9i-l**).

As observed in the previous 10 participants, EES applied over the *hemodynamic hotspot* led to pressor responses that scaled linearly with increasing amplitudes of EES (**Supplementary Fig. 9u**) and reduced the severity of orthostatic hypotension during a formal 10-minute tilt-table test (**Fig 5c** and **Supplementary Fig. 9m-t**). We next assessed the possibility of controlling blood pressure in closed-loop during formal and dynamic tilt-table testing sessions (**Fig. 5d**). We found that the complete purpose-built system maintained the desired blood pressure levels with high-fidelity despite varying orthostatic challenges (**Fig. 5e**).

### Independent proof of concept of the implanted system at an external site

We next asked whether our therapeutic strategy could be deployed independently by a center that had never been exposed to the implanted system and therapy. To assess this scalability, we conducted a new clinical study in the Netherlands (ClinicalTrials.gov ID: NCT05941819) (**Supplementary Fig. 10a**).

A neurosurgeon who had not been previously involved in the studies followed our procedures to implant the complete system in a participant who presented with severe orthostatic hypotension (**Supplementary Fig. 10b-f** and **Supplementary Table 2**). As observed in all the previous participants, intraoperative delivery of EES over the *hemodynamic hotspot* triggered robust pressor responses. Postoperative imaging confirmed the location of the lead over the *hemodynamic hotspot* (**Supplementary Fig. 10g-h**).

During the first postoperative session, we asked a physician without prior experience with EES to program the therapy based on anatomical principles to determine effective electrode configurations and parameters to modulate blood pressure with the system. The non-expert was able to program the therapy in only a few minutes. As early as the first session postoperatively, EES delivered over the *hemodynamic hotspot* in a seated position elicited robust pressor responses that increased the systolic blood pressure from 86 mmHg to 105 mmHg (**Supplementary Fig. 10i**). With EES, the participant was able to complete the 10-min tilt-table test, which was not possible without EES (**Supplementary Fig. 10j-n**). Closed-loop control of EES further improved the maintenance of blood pressure despite orthostatic challenges of varying severities (**Supplementary Fig. 10o-q**).

### Independent use of the system for blood pressure management

We concluded that the safety and efficacy profile of the new purpose-built implantable system and therapy were sufficient to enable the participants (n = 8 participants) to use the therapy independently at home, and thus to evaluate the long-term consequences of this therapy for the management of blood pressure after SCI. To support this evaluation, the participants were sent home with stimulation programs that were tailored to their individual pattern of hypotensive symptoms by a trained physician.

### Long-term efficacy of EES applied over the *hemodynamic hotspot*

Although the various participants included in our clinical studies were implanted with successively improved technologies, this diverse cohort of female and male participants spanning 18 to 74 years of age (**Supplementary Tables 1-2**) across 3 sites provided the opportunity to conduct a preliminary evaluation of the safety and effectiveness of EES applied over the *hemodynamic hotspot* to manage hemodynamic instability in people living with chronic SCI.

When evaluated up to two years after implantation, participants still exhibited severe orthostatic hypotension during a formal tilt table test when EES was turned off. They also continued exhibiting robust hemodynamic stability when EES was turned on, which allowed them to complete the tilt-table test (**Fig 6a-b** and **Supplementary Fig. 11f-k**). Moreover, formal evaluations confirmed the persistence of blood pressure stabilization evaluations during constant-varying orthostatic challenges when EES was controlled in closed-loop (**Supplementary Fig. 11m-o**). Pressor responses during tilt test with EES were associated with increased concentrations of circulating norepinephrine (**Supplementary Fig. 11l**), augmented splanchnic volume (**Supplementary Fig. 12i**), and increased sympathetic nerve activity (**Supplementary Fig. 12j**), which confirmed the activation of the sympathetic nervous system when EES was applied over the *hemodynamic hotspot*.

**Fig. 6.**
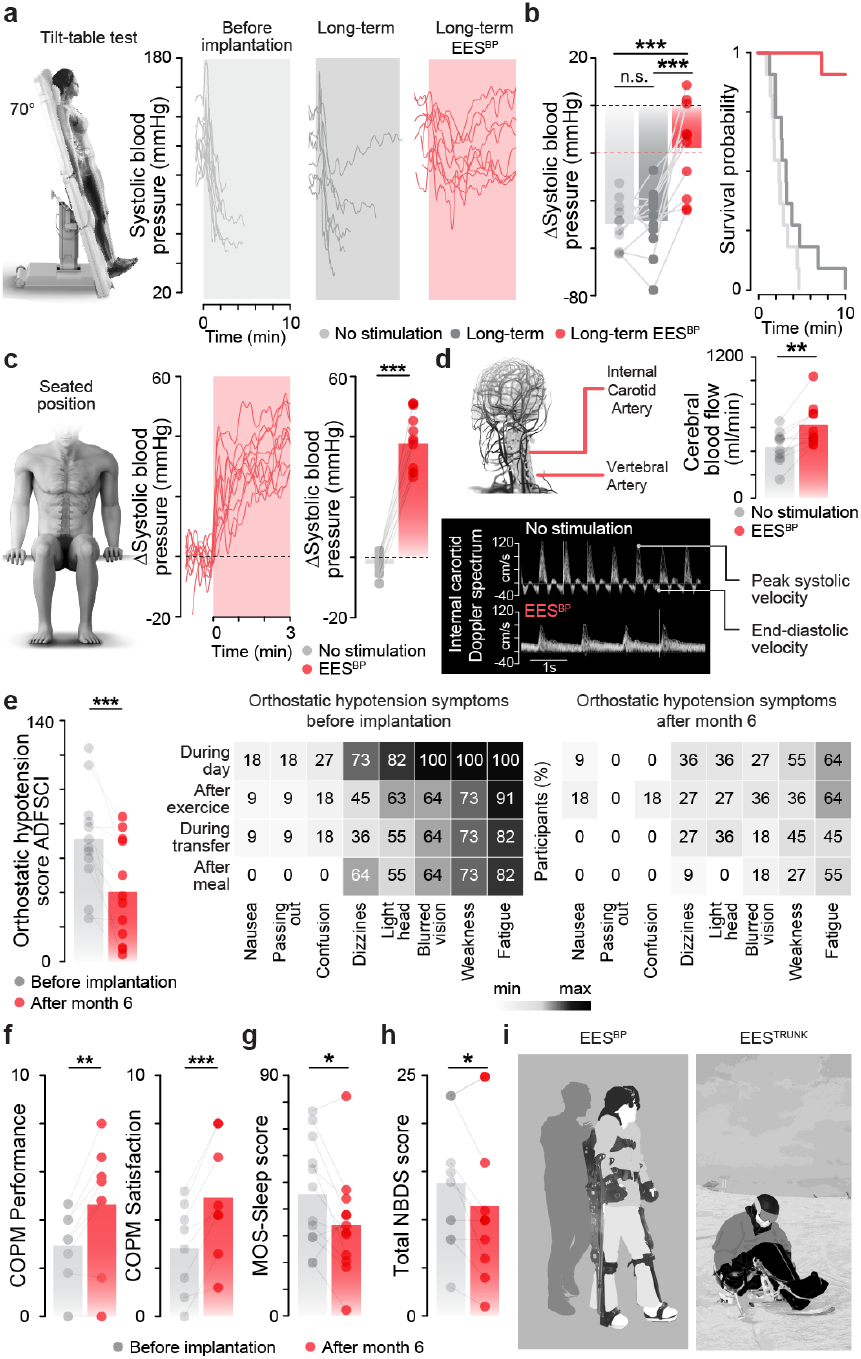
Long-term management of blood pressure improves quality of life. **a**, Changes in systolic blood pressure during a formal tilt-table test before implantation (*left*), without EES at least 3 and up to 24 months after implantation of the system (*middle*), and at the same time-point with EES (*right*) (n = 11/11 - Trials 1,3,4; n = 7 measured after at least 6 months of daily management of blood pressure with the system; n = 4 after at least 3 months). **b**, Bar plot reporting average changes in systolic blood pressure (*left*) during a format tilt-table test performed under the conditions as described in (**a**) (n = 11; repeated measures ANOVA with Tukey’s HSD). Corresponding Kaplan-Meier plot of exposure status to time sustained in tilt, segregated by the same condition as described in (**a**) (n = 11 tilts; mixed model Cox regression with likelihood ratio test estimate = 36.36; p-value = 3.0e-04). **c**, Changes in systolic blood pressure when delivering EES in seated position. Each line reports responses from a single participant to EES with default parameters for blood pressure regulation (n = 11/11 - Trials 1,3,4; n = 9 participants measured after at least 6 months of daily management of blood pressure with the system, n = 2 with at least 3 months). Bar plots report the average change in systolic blood pressure in a seated position that was quantified before and at 2 minutes after the onset of EES (n = 9 post-implant tilt measured with at least 6 months of EES, n = 2 post-implant tilt measured with at least 3 months of EES use). **d**, Quantification of the total cerebral blood flow without and with EES using a Doppler spectrum of the internal carotid without and with EES. The bar graph reports the total change in cerebral blood flow measured in a seated position without and with EES (n = 10, paired samples t-test, t = -4.32, p-value = 1.90e-03). **e**, Bar graph reporting Autonomic Dysfunction Following Spinal Cord Injury (ADFSCI) orthostatic hypotension score quantified before implantation of the system and after 6 months of management of blood pressure with the system (n = 12/14 - Trials 1-4, 2 participant explanted prior to 6 months; paired samples two-tailed t-test; t = 4.54; p-value = 8.45e-04), including a color-coded representation of the percentage of participants who experience each symptom described in this scoring system. **f**, Bar graph reporting the Performance (n = 8/8 - Trials 3-4, paired samples two-tailed t-test; t = -3.92; p-value = 5.7e-03) and Satisfaction (n = 8/8 - Trials 3-4, paired samples two-tailed t-test; t = -5.54; p-value = 8.7e-04) scores of the Canadian Occupational Performance Measurement (COPM) based on the baseline goals set by occupational therapists with the participants before the implantation of the system and after 6 months of management of blood pressure with the system. **g**, Bar graph reporting scores on the MOS-S - Sleep Problem Index II measured before implantation of the system and after 6 months of management of blood pressure with the system (n = 12/14 - Trials 1-4, 2 participants explanted prior to 6 months; paired samples t-test; t = 2.58; p-value = 2.52e-02). **h**, Bar graph reporting Neurogenic Bowel Dysfunction Score (NBDS) before implantation of the system and after 6 months of management of blood pressure with the system (n = 11/11 - Trials 1-3; paired samples t-test; t = 2.23; p-value = 4.95e-02). **i**, Usage of EES programs to support blood pressure stability during upright rehabilitation and maintain trunk stability during recreation activities such as sit-skiing.

Every tested participant continued demonstrating controllable pressor responses when EES was delivered over the *hemodynamic hotspot* while seated in their wheelchair, corresponding to the conditions of their daily life (**Fig. 6c** and **Supplementary Fig. 11a-e**). Pressor responses in seated conditions coincided with an increase in cerebral blood flow (**Fig. 6d**).

These objective physiological outcomes also concurred with a reduction in self-reported measures of orthostatic hypotension severity (**Fig. 6e**). These results showed that EES applied over the *hemodynamic hotspot* led to an immediate and durable reduction in the severity of hypotension in this cohort of individuals with SCI.

### Effective management of blood pressure improves quality of life

Our clinical outcomes thus far highlighted the durability of EES applied over the *hemodynamic hotspot* to manage blood pressure instability in people with SCI, but whether the resulting long-term improvement in blood pressure instability translated into an improvement in quality of life and an increased possibility to participate in activities of daily living remained unclear.

To address this question, we asked whether the participants relied on the therapy at home to manage their blood pressure throughout their daily life. We quantified the use of the therapy from the stimulation logs, and found that the participants turned the stimulation on throughout their waking hours, with increased usage to facilitate verticalization in the morning routine and to reduce postprandial hypotension after meals (**Supplementary Fig. 12a**). We confirmed the improvement in postprandial hypotension during in-clinic evaluations with controlled food intake (**Supplementary Fig. 12b**). The daily use of the therapy was accompanied by the medically-supervised cessation of all other conservative treatment options that participants had been relying on to manage hypotension, including abdominal binders and midodrine (**Supplementary Fig. 12d**). However, these improvements only manifested in the presence of the stimulation, since none of the participants demonstrated a reduction in the severity of orthostatic hypotension during formal tilt-tests when EES was turned off (**Fig. 6a-b** and **Supplementary Fig. 11g-k**). Evaluations of system usability^41^ confirmed that the participants could operate the therapy safely and effectively despite impairements in arm and hand movements (**Supplementary Fig. 12c**).

These multifaceted ameliorations translated into improvements in quality of life, as measured by formal outcome measures (**Fig. 6e**). Quantified increases in quality of life coincided with other subjective improvements that were reported to the study team (**Supplementary Fig. 12k**).

The beneficial impact of the therapy also extended to other aspects of life that could be potentially influenced by improved management of blood pressure and autonomic functions. For example, longitudinal outcome measures revealed improvements in sleep quality, bowel management, spasticity, and engagement in social activities (**Fig. 6f-i** and **Supplementary Fig. 12e**).

Since the participants regained the ability to sustain verticalization for extended periods of time when the system was turned on, they could engage in rehabilitation exercises that involved standing or walking in a motorized exoskeleton^42^ (**Fig. 6i** and **Supplementary Fig. 12f**). These tasks were not accessible with the system turned off since they experienced hypotensive symptoms in less than 2 minutes during verticalization.

The last three thoracic segments contain motor neurons innervating the trunk musculature. Consequently, EES applied over these segments activated trunk muscles. We reasoned that this added benefit of applying EES over thoracic segments could be leveraged to configure stimulation programs that concomitantly improve hemodynamic instability and neurological functions dependent on trunk and abdominal musculature. We mapped the optimal locations of EES to activate specific ensembles of trunk and abdominal muscles, which allowed us to customize stimulation programs to reinforce the activation of the trunk and abdominal musculature, aiming at improving trunk stability and coughing (**Supplementary Fig. 12g-h**). Quantifications of these neurological functions in well-controlled laboratory settings confirmed that these stimulation programs led to significant improvements (**Fig. 6i** and **Supplementary Fig. 12g-h**).

Participants leveraged these new stimulation programs to support activities of daily living. For example, one participant turned on trunk stability stimulation programs to facilitate the upright position of his trunk during meals, which freed his hands to manipulate his fork and knife without experiencing instability (**Fig. 6f**). Another participant used these programs to engage in recreational skiing activities since both his blood pressure and trunk stability were sufficient to enjoy these rides safely (**Fig. 6i**).

### Safety of the newly-developed system and therapy

Despite decades of experience in the management of neuropathic pain that have established the safety profile of EES, we systematically monitored for device- and procedure-related adverse events throughout the clinical studies.

All the participants have been operating the system to regulate their hemodynamic instability daily for up to two years. They were asked to report any safety event, including during home-use. All the serious and non-serious adverse device effects (S/ADE) reported across the 5 clinical studies for each participant are quantified and described in **Table 1** and **Supplementary Tables 4 - 7**.

**Table 1.** Overview of (serious) adverse events. Serious and non-serious adverse events and relatedness to investigational device or study procedure reported in all four clinical trials: Trial 1 (STIMO-HEMO, NCT04994886), Trial 2 (HEMO, NCT05044923), Trial 3 (HemON, NCT05111093), and Trial 4 (HemON-NL, NCT05941819).

| Event | Trial 1<br>NCT04994886 | Trial 2<br>NCT05044923 | Trial 3<br>NCT05111093 | Trial 4<br>NCT05941819 |
| --- | --- | --- | --- | --- |
| Number of implanted participants | 3 | 3 | 7 | 1 |
| **Serious adverse events** | **3** | **4** | **3** | **0** |
| *not related to device nor procedure* | 1 | 1 | 2 | 0 |
| *related to device* | 0 | 2 | 0 | 0 |
| *related to study procedure* | 2 | 1 | 1 | 0 |
| **Non-serious adverse events** | **16** | **4** | **29** | **5** |
| *not related to device nor procedure* | 3 | 3 | 16 | 3 |
| *related to device* | 5 | 1 | 4 | 2 |
| *related to study procedure* | 8 | 0 | 9 | 0 |

Across the 8 participants implanted with the newly-developed system or components of this system, no serious adverse events related to the device were reported. Only one serious adverse event involving an uncontrolled increase in blood pressure due to the inadvertent removal of a urinary probe occurred while the system was turned off (**Supplementary Table 7**). Uncontrolled increases in blood pressure during EES occurred only once across hundreds of testing sessions conducted over a period that spanned several months and involved the delivery of EES across a broad range of parameters. This non-serious device-related adverse event occurred while targeting the trunk musculature, but ceased as soon as EES was switched off.

## Discussion

We conducted two epidemiological analyses and five complementary clinical studies involving implantable medical devices that i) established the burden of hypotension complications in people with SCI, ii) provide data that supports the lower thoracic spinal cord as the optimal location to apply EES for treating hemodynamic instability, iii) enabled the stepwise development of a purpose-built system harnessing the understanding of the mechanisms through which EES regulates hemodynamics in people with SCI, and iv) provides preliminary safety and efficacy data of this system to reduce hypotensive complications throughout the day and thus improve quality of life.

Before considering the development of a therapy based on EES to improve hemodynamic instability after SCI, we thought that it was essential to address the controversy on the optimal location to restore hemodynamics with EES^16–21^. We found that EES applied to the *hemodynamic hotspot* is more effective compared to EES applied to the lumbosacral spinal cord. This result was not surprising since attempts to control hemodynamics by stimulating the lumbar region of the spinal cord are incongruent with the basic and well-conserved anatomy of the ancestral autonomic nervous system. To date, the mechanisms underlying the fortuitous observations of pressor responses with EES applied over the lumbosacral spinal cord remain unknown. We surmised that two mechanisms may explain these observations.

The first mechanism that we previously exposed in pre-clinical models is the off-target activation of the dorsal roots innervating the last thoracic segment as they exit the spinal column through the T12 intervertebral foramen^1^, which roughly coincides with upper lumbar spinal cord segments. Our mapping experiments demonstrate that the optimal coverage of the last three segments of the thoracic spinal cord, especially the penultimate thoracic segment, is essential to maximize pressor responses to EES. Consequently, the inefficient recruitment of only the last thoracic root is unlikely to be an effective and reproducible methodology to manage hemodynamic instability across the population of people with SCI.

The second mechanism, which is the most concerning for the safety of people with SCI, is the risk that EES applied over the lower aspect of the lumbosacral spinal cord directly activates the pathways that cause life-threatening autonomic dysreflexia^2,43^. Sensory information originating from the overstimulated afferents of the bladder and bowel are the most common triggers of autonomic dysreflexia in people with SCI. Afferents from these visceral organs project to the spinal cord through the lower lumbar and upper sacral dorsal roots, which are directly recruited when applying EES over the lumbosacral spinal cord, especially with configurations targeting the lower lumbar dorsal roots^18,21,40^. Since no other neuronal populations that contribute to the control of blood pressure exist in this region of the spinal cord^1,22^, the only explanation for pressor responses elicited by the application of EES over the lower lumbar and sacral spinal cord is the activation of the pathways that can cause autonomic dysreflexia. Indeed, preclinical evidence demonstrated that daily application of EES over the lower lumbosacral segments in rodents with complete upper thoracic SCI augments the density of axonal projections from Vsx2-expressing neurons in the lumbosacral spinal cord onto Vsx2-expressing neurons in the lower thoracic spinal cord^25^. These neurons are embedded in the neuronal architecture that causes autonomic dysreflexia. Consequently, this aberrant exacerbation of the neuronal architecture responsible for autonomic dysreflexia doubled the severity of pressor responses that characterize this life-threatening condition^25^. Although the implication of Vsx2-expressing neurons has not been confirmed in humans, single-cell and spatial transcriptomic technologies^44^ have advanced sufficiently to enable the exploration of this mechanism, even in human tissue. With this limitation in mind, this mechanism appears to be the most likely explanation for the reported improvements of resting hypotension and orthostatic hypotension in individuals with SCI who underwent extensive stimulation of the lower lumbosacral segments.

Together, consistent results obtained in a total of 14 participants in 4 clinical studies across 3 countries conducted at 3 independent clinical centers with independent clinical teams provide robust preliminary data on the safety and efficacy of EES applied over the hemodynamic hotspot to treat hemodynamic instability in people living with chronic SCI. These findings establish a path forward to treat the underappreciated, treatment-resistant hypotensive complications due to SCI. Completing this path will require a pivotal device trial that evaluates the safety and efficacy of this therapy.

Neurodegenerative disorders such as typical and atypical Parkinson’s disease lead to comparable hypotensive complications, which are due to the loss of sympathetic neurons in the brainstem, spinal cord, and peripheral ganglia. We recently showed that EES durably reduced the severity of orthostatic hypotension in a patient with multiple system atrophy^28^. While the regulation of blood pressure with EES is contingent on the partial preservation of both preganglionic and ganglionic neurons^1,22,25^ 1,20,25, these results open the possibility that our implantable system may also contribute to the management of severe orthostatic hypotension conditions in people with neurodegenerative disorders.

## Methods

### Epidemological analysis

#### Spinal Cord Injury Community Survey (SCICS)

Ethical approval was obtained from an independent ethics board (Veritas Independent Review Board) and the Research Ethics Board of Université Laval (principal investigator’s institution). Ethical approval from local research ethics boards was also obtained to recruit from spinal cord injury centers across Canada. Individuals with SCI (n = 1,479) across Canada were recruited using a national consumer awareness campaign and provided written informed consent to participate^26,27^. The survey consisted of a series of variables identified by healthcare and service providers, researchers, as well as individuals with SCI, including demographics, secondary health complications, comorbidities, SCI-related needs, healthcare utilization, community participation, quality of life, as well as overall health ratings^26,27^. Participants were asked how often they experienced symptoms such as light-headedness and/or dizziness consistent with symptoms of orthostatic hypotension in the past 12 months. Responses were ranked on a 6-point ordinal severity scale ranging from zero (i.e., “Never”) to 5 (i.e., “Every day”). Participants were also asked if they received or sought out treatment concerning these symptoms on a two-point scale (i.e., “Yes” or “No”), along with the degree to which it limited activities from zero (i.e., “Never”) to 5 (i.e., “Every day”). Participants were also asked if they had experienced specific problems, such as heart disease, in the past 12 months. Participants’ American Spinal Injury Association Impairment Scale (ASIA) was estimated using responses to questions about lesion level and sensorimotor/mobility capabilities. A binary approach was used for the evaluation of outcome variables including the level of injury (i.e., cervical SCI vs. non-cervical SCI), the severity of injury (i.e., functionally complete vs. incomplete), presence of orthostatic hypotension (i.e., yes vs. no), and orthostatic hypotensive symptoms (i.e., yes vs. no). For variables ranked on a 6-point ordinal scale, lower scores (i.e., 0-3) were categorized as “No” and higher scores (i.e., 4-5) were categorized as “Yes.”

#### Autonomic dysfunction in a sample of individuals living with SCI

254 participants were surveyed with the ADFSCI questionnaire (see Autonomic Dysfunction Following Spinal Cord Injury) to measure the severity and frequency of orthostatic hypotension in a diverse sample of individuals with SCI. From this population, 17 individuals were pseudo-randomly selected to participate in a single formal, 10-minute tilt-table test (see Orthostatic challenge with tilt-table test). Hemodynamic measurements were taken via finger plethysmography (see Post-operative hemodynamic monitoring), and data was analogously acquired with the LabChart Software (ADInstruments, Dunedin, New Zealand) and upsampled to 1000 Hz.

### Clinical trials

#### Study designs and objectives

All experiments were carried out as part of 4 clinical safety (primary objective) and preliminary efficacy (secondary objectives) trials: **STIMO-HEMO** (NCT04994886, CHUV, Lausanne, Switzerland), **HEMO** (NCT05044923, University of Calgary, C algary, Canada), **HemON** (NCT05111093, CHUV, Lausanne, Switzerland), Amendement of HemON (NCT05111093, CHUV, Lausanne, Switzerland) and **HemON-NL** (NCT05941819, Sint Maartenskliniek, Ubbergen, The Netherlands). The HemON trial was amended after 4 participants to include the new thoracic paddle lead. All 4 trials and amendments received approval by the local ethical committees and national competent authorities. A summary of information related to each study is provided in **Supplementary Table 1**. All the participants signed a written informed consent at enrollment in the trial. All participants had the option to indicate consent for the publication of identifiable images or videos. All surgical and experimental procedures were performed at the investigational hospital sites, including the neurosurgery department of Lausanne University Hospital (CHUV) in Lausanne, Switzerland, the neurosurgery department of Foothills Medical Center in Calgary, Canada, and the neurosurgery department of the Radboud University Medical Center in Nijmegen, The Netherlands. The study involved eligibility and baseline assessments conducted before the surgery, the neurosurgical implantation of an investigational device, a postoperative period during which EES protocols were configured, and long-term follow-up periods, including independent use of EES at home. To date, 13 individuals completed the intensive part of the study.

#### Study participants

Fourteen individuals who had suffered a traumatic SCI participated in one of the 5 studies. Demographic data and neurological status were evaluated according to the International Standards for Neurological Classification of Spinal Cord Injury^45^, and are reported in **Supplementary Table 2**.

For the STIMO-HEMO and HEMO clinical trials, participants were eligible if they were aged between 18 and 70 years old, had a radiologically confirmed SCI located between C3 and T6 (inclusive), were neurologically classified as A or B on the American Spinal Cord Injury Association (ASIA) Impairment Scale (AIS), had stable medical, physical and psychological condition under the discretion of the investigators, had the injury for at least 1 year, did not have any required spinal instrumentation within the past 6 months, were able and willing to attend all scheduled appointments, and had confirmed orthostatic hypotension and autonomic dysreflexia. Exclusion criteria included individuals in any emergency situations, participants with any diseases and conditions that would increase the morbidity and mortality of the spinal cord injury surgery, participants were unable to withhold antiplatelet, or anticoagulation agents perioperatively, had history of myocardial infarction or cerebrovascular events within the past 6 months, had a current, or anticipated need for opiod pain medications, had pain that would prevent full participation in the rehabilitation program in the judgment of the investigators, had a clinically significant mental illness in the judgment of the investigators, received botulinum toxin injections within 6 months of enrollment, presence of significant pressure ulcers, recurrent urinary tract infection refractory to antibiotics, unhealed spinal fractures, known or suspected drug or alcohol abuse, presence of indwelling baclofen or insulin pump, and any other conditions that would make the subject unable to participate in testing in the judgment of the investigators. Women pregnant during screening, or breastfeeding were also excluded.

For the HemON clinical trial, eligible participants were 18 years of age or older, had a SCI located between C3 and T6 (inclusive), had their injury for at least 1 month, had stable medical, physical, and psychological conditions as considered by the investigators, spoke the French or English language to interact with the study team, available to participate in good faith and attend scheduled appointments per study conditions, and had confirmed orthostatic hypotension. Exclusion criteria include SCI related to neurodegenerative disease, diseases and conditions that would increase the morbidity and mortality of the surgery, the inability to withhold antiplatelet, or anticoagulation agents perioperatively, history of myocardial infarction or cerebrovascular events within the past 6 months, other conditions that would make the subject unable to participate in testing in the judgment of the investigators, clinically significant mental illness in the judgment of the investigators, having received botulinum toxin vesical and non-vesical injections within 3 months of enrollment, presence of significant pressure ulcers, recurrent urinary tract infections refractory to antibiotics, presence of indwelling baclofen or insulin pumps, other clinically significant concomitant disease states including but not limited to renal failure, hepatic dysfunction, cardiovascular disease, etc., inability to follow study procedures due to language problems, psychological disorders, or dementia, participation in another study with investigational drugs within the 30 days preceding and during the present study, and any relation to the investigator, his/her family members, employees, and other dependent persons. Women who are pregnant or breastfeeding, lack safe contraception if with childbearing capacity, or had the intention to become pregnant during the course of the study were excluded.

For the HemON-NL trial, eligible participants were 18 years of age or older had a SCI located between C3 and T6 (inclusive), had a traumatic SCI, had their injury for at least 1 month, were neurologically classified as AIS A, B, C or D, had stable medical, physical and psychological conditions as considered by the investigators, spoke the Dutch or English language to interact and understand the study team, available to participate in good faith and attend scheduled appointments per study conditions, had if needed continuous support from personal caregiver in daily life during the study visit (including independent transport) and had confirmed orthostatic hypotension. Exclusion criteria include SCI related to diseases and conditions that would increase the morbidity and mortality of the surgery, diseases and conditions that would require regular magnetic resonance imaging (MRI), the inability to perform an MRI due to metal, magnetic or electrical device in the body (e.g. oral implant with magnet, metal splinter, neurostimulator, artificial heart valve, clips, stents etc.) as assessed by the MRI form of Sint Maartenskliniek, the inability to withhold antiplatelet, or anticoagulation agents perioperatively, history of myocardial infarction or cerebrovascular events within the past 6 months, other conditions that would make the subject unable to participate in testing in the judgment of the investigators, clinically significant mental illness in the judgment of the investigators, having received botulinum toxin vesical and non-vesical injections within 3 months of enrollment, presence of significant pressure ulcers, recurrent urinary tract infections refractory to antibiotics, presence of indwelling baclofen or insulin pumps, other clinically significant concomitant disease states including but not limited to renal failure, hepatic dysfunction, cardiovascular disease, etc., inability to follow study procedures due to language problems, psychological disorders, or dementia, participation in another study with investigational drugs within the 30 days preceding and during the present study, and any relation to the investigator, his/her family members, employees, and other dependent persons. Women who are pregnant or breastfeeding, lack safe contraception if with childbearing capacity, or had the intention to become pregnant during the course of the study were excluded.

#### Safety outcomes

All the feasibility clinical trials outlined in **Supplementary Table 1** evaluated the safety and preliminary efficacy of the investigational system and/or therapy to restore hemodynamic stability. The primary outcome measures assess the occurrences of serious adverse events and adverse events that are deemed related or possibly related to the investigational system or study procedure during the course of studies. All reportable safety events have been reported per local regulation to the respective authorities (Health Research Ethics Board at the University of Calgary, Ethics Committee CER-VD, and competent authorities Swissmedic in Switzerland and METC Oost-Nederland in Nijmegen). Data safety monitoring boards have been appointed in all studies and consulted on a yearly basis or adHoc in case of reportable events. All adverse device effects for the 4 clinical trials are summarized in **Supplementary Tables 4-7**. Two serious adverse events related to the device leading to explantation occurred during the course of the HEMO study due to postoperative infections. Device explantations were reported to local authorities and to the manufacturer. Both were resolved without sequelae following explantation surgery. In studies evaluating the safety of the new implanted device, one reportable serious adverse event related to study procedure occurred (subpubic catheter came out during a rehabilitation session while the stimulation was turned off).

#### Imaging Data

All the participants underwent structural 3T magnetic resonance imaging (MRI) and computed tomography (CT) imaging of the thoracolumbar spine to enable the elaboration of a personalized anatomical model of the spine and guide the preoperative planning. MRI acquisitions were conducted in a supine position with both arms on the sides. Prior to image acquisition, shim boxes were applied to correct for magnetic field inhomogeneities. All sequences were performed without gadolinium-based contrast agent administration. In the STIMO-HEMO and HemON clinical trials, MRI was performed on a Magnetom PrismaFit (Siemens Healthineers, Erlangen, Germany) with 18-channel body and 32-channel spine array coils. The standard MRI protocol comprised the following four pulse sequences: a) 2D sagittal T2-weighted turbo spin-echo (repetition time (TR), 3080 msec; echo time (TE), 98 msec; voxel size, 0.6 × 0.6 × 3 mm^3^); b) T2-weighted SPACE (Sampling Perfection with Application-optimized Contrasts using different flip angle Evolution) sequence (TR, 1500 msec; TE, 135 msec; interpolated voxel size, 0.4 × 0.4 × 0.8 mm^3^); c) 3D axial T2-weighted SPACE with ZOOMit (dynamic excitation pulses to achieve selective/zoomed field-of-view) software (TR, 2500 msec; TE, 106 msec; interpolated voxel size, 0.3 × 0.3 × 0.5 mm^3^); and d) 3D coronal T2-weighted TrueFISP (True Fast Imaging with Steady state Precession) (TR, 6.04 msec; TE, 3.02 msec; interpolated voxel size, 0.3 × 0.3 × 0.6 mm^3^). The total scan time was less than 35 minutes overall. The HemON trial MRI protocol consisted of a subset of these sequences (no 3D axial T2-weighted SPACE with ZOOMit as this trial does not include an implant at the lumbosacral level). In the HEMO trial, a similar protocol was performed on a Discovery MR750 (GE Healthcare, Waukesha, WI, USA) with an 8-channel cervical thoracic lumbar coil. The following two sequences were performed: a) 3D coronal T2-weighted FIESTA-C (Fast Imaging Employing Steady-state Acquisition) (TR, 7.65 msec; TE, 3.48 msec; voxel size, 0.4×0.4×0.4 mm^3^); and b) 3D sagittal T2-weighted CUBE sequence (TR, 2000 msec; TE, 90 msec; interpolated voxel size, 0.8×1.25×0.8 mm^3^). The total scan time was 84 min overall. Finally, in the HemON-NL trial, the standard protocol was translated to an Ingenia Omega (Philips Healthcare, Best, The Netherlands) with a 32-channel coil. The protocol comprised the following two sequences: a) sagittal T2-weighted FFE (Fast Field Echo) (TR, 3.34 msec; TE, 8.1 msec; interpolated voxel size, 0.75×0.75×3 mm^3^); and b) coronal Balanced FFE (TR, 7.0 msec; TE, 3.5 msec; interpolated voxel size, 0.75×0.75×3 mm^3^). The total duration of the acquisition was approximately 20 minutes overall.

CT scans were acquired in a supine position with both arms on the sides. No contrast agents were administered. In the STIMO-HEMO and HemON clinical trials, CT of the trunk including the pelvis was performed on a 256-detector row CT system (Revolution Apex, GE Healthcare) using the following acquisition parameters: tube potential, 120 kVp; tube current, 115-200 mA, by enabling automatic tube current modulation; gantry revolution time, 0.35 sec; beam collimation, 128 × 0.625 mm; pitch, 0.508. CT images were reconstructed with a field of view of 150 × 150 and 400 × 400 mm^2^ for the spine and pelvis, respectively; at a section thickness/interval of 1.25/0.625 and 1.25/1.25 mm, respectively, with both smooth (Standard) and sharp (Bone plus) convolution kernels and a deep learning image reconstruction algorithm (TrueFidelity, GE Healthcare; medium strength level) for image noise reduction. The nominal voxel size was 0.29 × 0.29 × 0.625 and 0.78 × 0.78 × 1.25 mm^3^ for the spine and pelvis, respectively. In the HEMO trial, a Revolution Discovery scanner (GE, Boston, MA, USA) was used to collect Standard and Bone Plus reconstructions (pitch 0.531:1, slice thickness 1.25 mm, interval: 1.25, 120 kV, 115-200 mA, rotation time: 0.4 s) of the thoracolumbar spine. In the HemON trial, an Ingenuity Core 128 scanner (Philips) was used to collect standard iDose and IMR reconstructions.

#### Personalized anatomical models of the spine

We trained two different nnU-net networks^46^ on a labeled data set of 15 healthy participants from a previous study^30^. The first network was trained on high-resolution MRI to segment the cerebrospinal fluid, the white matter, and spinal root tissues. The second network was trained on low-resolution MRI data to segment vertebral bodies, intervertebral discs, and the spinal canal. We trained the networks on 12 participants and tested on 3 subjects. We achieved high dice scores for each tissue (cerebrospinal fluid: 95.50; white matter: 90.55; roots: 71.88; vertebrae: 89.90; intervertebral discs: 91.22). When applying the networks to a new data set, we corrected the segmentation manually if needed. The segmentation of the roots was used as guidance to trace each spinal root individually. The rostral location of each root was constrained within the white matter. For each spinal quadrant (dorsal/ventral left/right), the end-of-root within the white matter points was cut 1 mm within the white matter and a spline was made with them. These end-of-root lines were then guidelines to force rootlets to attach to them, forcing them to end within the white matter. The procedure to generate the rootlets was described previously^30^. By forcing each rootlet to end in the end-of-roots guidelines, we made sure the rootlets followed the shape of the spine, thereby increasing robustness to scoliosis. We cut the white matter with respect to the rootlet divisions and obtained henceforth the spinal levels.

We used an anatomically informed framework trained on the VerSE datasets^47^ to segment the different vertebrae on CT imaging^48^. The authors of the framework showed a dice score of 91.04 on a large test set. Having both segmentations from MRI and CT spaces, we automatically placed labels on each vertebrae segmentation on MRI data. We then co-registered the CT data to the MRI space automatically using the Spinal Cord Toolbox^49^.

A computer-aided design (CAD) model of the 16 electrodes composing the paddle lead was positioned over the targeted dorsal root entry zones. Several visualizations of the 3D personalized anatomical model were then generated and shared with the clinical and neurosurgical team to guide preoperative surgical planning. Finally, a virtual paddle lead was placed over the selected location of the spinal cord and, and then projected onto a large screen in the operation room to guide the neurosurgeon in the insertion of the lead.

#### Neurosurgical intervention

The participants were put under protocolized propofol-based total intravenous general anesthesia and were placed in a prone position. Preoperative surgical planning informed the neurosurgeon about the vertebral entry-level and predicted optimal paddle lead position. Based on this knowledge, lateral and anteroposterior fluoroscopy x-rays were performed intra-operatively to identify the location of the laminotomies. A midline skin incision of approximately 5 cm on the back was performed, the fascia opened and the muscles retracted bilaterally. Excision of the midline ligamentous structures and a laminotomy at the desired entry level enabled the insertion of the paddle lead below the vertebra. For participants of the STIMO-HEMO and HEMO trials, a second skin incision or extended opening caudally was made and a second laminotomy was performed in the lumbar region based on the preoperative planning to allow for the insertion of the paddle lead over the lumbosacral spinal cord. The paddle lead(s) (Specify 5-6-5, Medtronic Inc, Minneapolis, MN, USA or ARC^IM^ Thoracic Lead, ONWARD Medical N.V, Eindhoven, Netherlands) are inserted and placed over the midline of the dura-mater and advanced rostrally to the target position under repeated fluoroscopies. Electrophysiological recordings were acquired using standard neuromonitoring systems (Cascade Elite, Cadwell Industries, Kennewick, USA or ISIS Xpress, Inomed Medizintechnik, Emmendingen, Germany). Single-pulses of EES were delivered at 0.5 Hz and increasing amplitude to elicit muscle responses that were recorded from subdermal (Neuroline Twisted Pair Subdermal, 12 × 0.4 mm, Ambu A/S, Ballerup, Denmark) or intramuscular needle electrodes (Inomed SDN electrodes, 40 × 0.45 mm, Inomed Medizintechnik, Emmendingen, Germany). The symmetry of the responses and the expected rostrocaudal distribution of muscle responses informed the corrections for lateral and rostrocaudal positioning. When the paddles deviated from a straight midline position, small additional laminotomies were made to remove bony protrusions and guide the paddle to a midline placement. Once the final position was determined, the paddle lead(s) was (were) anchored to the muscular fascia. In the STIMO-HEMO, HemON, and HemON-NL trials, the back opening was temporarily closed and the participants were put in lateral decubitus. Subsequently, the back incision was reopened and an abdominal incision of about 5 cm was made to implant each pulse generator in a subcutaneous pocket. In the HEMO trial, incisions of about 5 cm were made bilaterally in the upper buttocks region and subcutaneous pockets were created. The cables of the paddle lead were then tunneled between the back opening and subcutaneous pockets to be connected to the implantable pulse generator (Intellis, Medtronic Inc, Minneapolis, MN, USA or ARC^IM^ IPG, ONWARD Medical N.V., Eindhoven, Netherlands). The implantable pulse generator(s) was (were) implanted in the subcutaneous pocket(s) and all incisions were finally closed. In the STIMO-HEMO trial, both paddle lead cables were tunneled from the same flank side and tunneling was performed between the 2 abdominal subcutaneous pockets.

### Technological framework

#### Stimulation optimization

The configuration of the electrodes for EES^XX^ programs was guided by the understanding of the mechanisms through which EES regulates blood pressure (**Fig. 2a**). This configuration involved three steps. First, an intraoperative mapping was conducted to identify the relevant rows of electrodes to target the hemodynamic hotspot^1^ and to elicit the largest pressor response. Second, postoperative imaging was used to update the anatomical models and thus estimate the location of the electrodes that maximized recruitment of the dorsal root entry zones projecting to the hemodynamic hotspot. Third, a single postoperative session was conducted to quantify pressor responses elicited by each row of electrodes. Both monopolar and multipolar configurations of electrodes were tested to ensure the maximal activation of the hemodynamic hotspot. Stimulation frequency was set at 120 Hz, since we established that this frequency maximized pressor responses^1^. Multiple electrode configurations that each targeted a specific region of the hemodynamic hotspot. Each electrode configuration was delivered with 2 ms delay between each pulse, forming a traveling wave over the spinal cord. In the STIMO-HEMO and HEMO trials, pulse-width was mapped between 100 µs - 500 µs. Five out of six participants had a pulse width of 300 µs, while one participant (P3) had a pulse width of 500 µs. For the HemON and HemON-NL trials, the pulse width was set at 300 µs. The amplitude was set based on the increase in the current delivered through each electrode configuration until the systolic pressure increased by 20 mmHg, the diastolic pressure increased by 10 mmHg, or the participant reported any discomfort such as muscle contractions, or unpleasant sensation such as strong tingling. To validate the combined effect of multiple electrode configurations, we compared the amplitude of the pressor responses when EES was delivered with a configuration targeting a single level of the hemodynamic hotspot, versus all the levels embedded in the hemodynamic hotspot. All other stimulation parameters were maintained unchanged.

#### Intraoperative hemodynamic monitoring

Intraoperative beat-to-beat hemodynamic data was obtained from a pressure transducer connected to an arterial line cannula placed in the distal radial artery. The arterial line system was zeroed to atmospheric pressure. Arterial cannulation was performed either by palpation or with ultrasound guidance under sterile conditions. Adjustments in the anesthetic depth as well as administration of vasoactive medications or significant volumes of intravenous fluids were avoided during hotspot testing. All data was recorded using VitalRecorder software (VitalDB, Department of Anesthesiology and Pain Medicine, Seoul National University College of Medicine, Seoul, Korea) at a sampling frequency of 125 Hz. Alternatively, the arterial line was recorded with LabChart^24^ (ADInstruments, Dunedin, New Zealand).

#### Postoperative hemodynamic monitoring

Beat-to-beat blood pressure and heart rate were acquired using finger plethysmography (Finometer, Finapres Medical Systems; Amsterdam, Netherlands). Beat-by-beat blood pressure was calibrated to brachial artery blood pressure collected using an arm cuff embedded and synchronized with the Finometer^50–54^. Brachial arterial pressure was sampled at 200 Hz, while the systolic, diastolic, and mean arterial pressure were extracted from the calibrated arterial pressure at 1 Hz. The heart rate was also sampled at 1 Hz. Raw data and automatically extracted hemodynamic parameters were saved by and exported from the Finometer.

#### Off-label investigational system

The investigational system used in the STIMO-HEMO and HEMO clinical trials consisted of a set of CE-marked, FDA-approved medical devices used off-label. Two implantable pulse generators (lntellis™ with AdaptiveStim™, Medtronic Inc, Minneapolis, MN, USA) were connected to a paddle lead (Specify™ 5-6-5 SureScan™ MRI, Medtronic Inc, Minneapolis, MN, USA). These implants are indicated for chronic pain management. A tablet application with a wireless communicator device (Intellis™ clinician programmer, Medtronic Inc, Minneapolis, MN, USA) was used by the clinical team to set up the system and optimize the stimulation parameters. A remote control device and transcutaneous charger device (Patient Programmer and Recharger, Medtronic Inc, Minneapolis, MN, USA) were used by the participants to charge the implanted pulse generators and to turn EES programs on and off during their daily life, as well as adapt parameters of EES programs defined by the clinical team.

#### Purpose-built investigational system to treat hemodynamic instability

The investigational system used in the HemON and HemON-NL clinical trials were designed and built for treating hemodynamic instability based on the understanding of the mechanisms through which EES regulates blood pressure. A step-wise approach was followed. All participants were implanted with a purpose-built implantable pulse generator ARC^IM^ (ONWARD Medical N.V., Eindhoven, Netherlands) that communicates with a purpose-built ecosystem of control devices. The first 4 participants of the HemON clinical trial were implanted with the off-label paddle lead (Specify™ 5-6-5 SureScan™ MRI, Medtronic Inc, Minneapolis, MN, USA) as the participants of STIMO-HEMO and HEMO clinical trials, whereas all other participants were implanted with a newly-designed novel, purpose-built paddle lead (ARC^IM^ Thoracic Lead, ONWARD Medical N.V., Eindhoven, Netherlands).

#### Purpose-built implantable pulse generator and communication ecosystem to treat hemodynamic instability

The purpose-built ARC^IM^ implantable pulse generator is a new 16-channel neurostimulation platform that was specifically developed to regulate neurological functions with EES. The ARC^IM^ implantable pulse generator controls and delivers current-controlled stimulation pulses that can be predefined by the user, or controlled in real time. The system consists of a hermetically sealed, biocompatible can that surrounds the electrical components and a rechargeable battery that enables the delivery of electrical current. The external envelope is composed of two main components: the header containing the connector block that enables connection with 2 8-contact lead connectors and 2 coils for charging and communication, and the can with a rechargeable battery and electronics circuits. The ARC^IM^ implantable pulse generator was developed according to all applicable standards for medical device development. Conventional biomedical technologies were used to fabricate the system and extensive bench and in-vivo testing was performed to verify its performance.

The implantable pulse generator was implanted subcutaneously in a pocket created in the abdomen. The ARC^IM^ communication Hub ensured exchange of information with the implanted pulse generator using Near Field Magnetic Induction (NFMI), and also serves as a wireless charging station for the battery embedded in the implanted pulse generator. The communication hub is worn on a belt over or in proximity to the location of the implanted pulse generator. This Hub hosts a Bluetooth Low Energy (BLE) chip that enables fast, reliable wireless communication with external sensors or programmers. This versatility enables the acquisition of varying control signals and rapid exchange of information between the communication hub and the implanted pulse generator, which together establish the conditions for closed-loop control of stimulation parameters with latencies as low as 25ms between the generation of the stimulation command and the execution of this command. The Hub communicates with external programmers such as the ARC^IM^ Clinician Programmer onto which an Android App designed for clinicians allows the configuration of EES programs and evaluation of the system. When EES programs are deemed safe for personal use, the Clinician Programmer provides the possibility to make this stimulation program available to the participant.

The participants, or their caregivers, can control the system through the ARC^IM^ Personal Programmer. This Android Watch application allows users to select, start, and stop EES programs, as well as to tune stimulation amplitudes within predefined safety limits determined by the clinicians.

Device errors, paddle lead impedances, and daily stimulation utilization were extracted from usage logs across all devices. Furthermore, the Clinician Programmer includes an application programming interface, termed the ARC^IM^ API, that enables other programming softwares to control the stimulation, e.g. for closed-loop control of the stimulation.

All devices and softwares are adherent to the applicable standards and their performance was extensively tested. The entire system received the equivalent of an investigational device exemption from the competent Swiss authorities.

#### Purpose-built paddle lead

The ARC^IM^ Thoracic Lead developed by ONWARD Medical is a new 16-electrode paddle lead that incorporates a configuration of electrodes optimized to achieve the selective recruitment of the dorsal root entry zones projecting to the last 3 segments of the lower thoracic spinal cord. The configuration of the electrodes was determined based on a digital library of human thoracic spinal cords that we elaborated from CT and MRI acquisitions (**Supplementary Fig. 4**). The average length and width of spinal segments were calculated from our library and existing literature^30,34–39^. The combination of these 2 datasets generated global averages and standard deviations for each level of the lower thoracic spinal cord, which determined the total length of the hemodynamic hotspot. We created a normal distribution with the calculated global means and standard deviations and found that a length of 64.2 mm accurately represents the combined length of the 3 targeted spinal segments for over 95% of the population. This new paddle lead was designed following all applicable standards for medical implant development and fabricated using conventional biomedical technologies. Extensive bench and in-vivo testing was conducted to validate the mechanical, electrical, and biocompatibility robustness of the paddled lead. The equivalent of an investigational device exemption was granted by the competent Swiss competent authorities.

#### System for closed-loop control of blood pressure

The purpose-built investigational system was embedded in a framework for closed-loop control of systolic blood pressure based on real time monitoring of hemodynamic parameters. For this purpose, changes in systolic blood pressure were measured with the Finometer (see Postoperative hemodynamic monitoring). This signal was acquired using an analog-to-digital converter (Delsys Trigno Avanti Analog, Delsys Inc., Natick, Massachusetts) that wirelessly streamed the data to a sensor base station (Delsys Trigno Research+, Delsys Inc., Natick, Massachusetts). The 1 Hz systolic blood pressure measured with the Finometer was up-sampled to 1,257 Hz by the Trigno system, which was connected through USB to a Windows computer onto which a medical-grade software developed by EPFL was running. This software^30^ enables closed-loop control of the ARC^IM^ system through the ARC^IM^ API. The specific closed-loop algorithm used for the control of systolic blood pressure was implemented in a multi-threaded Python application that interfaces with the GDrive+ software via TCP/UDP communication enabling the reception of systolic blood pressure signals and delivery of stimulation updates.

#### Algorithm for closed-loop control of blood pressure

The systolic blood pressure signal was smoothed with an exponential weighted moving average over a 3-second window with a forgetting factor of 0.3. This elaborated signal was sent as an input to a Proportional-Integral-Derivative (PID) controller. To minimize steady-state errors and minimize system saturation, the integral term was penalized over time when the stimulation remained unchanged at the minimum or maximum amplitude of EES. Additionally, a forgetting factor on the integral term reduces error accumulation. A minimum (0 mA) and maximum amplitude (participant dependent) amplitude range was set to ensure the comfort and safety of the participant. The controller target was defined based on average blood pressure measurements from 10-minute tilt-table tests with EES (see Postoperative hemodynamic monitoring). A target range of +/- 3 mmHg was set around the user-defined target to account for intrinsic blood pressure variability. If the change in EES amplitude was greater than 0.5 mA, the controller incrementally ramped in steps of 500 ms per 0.5 mA for additional comfort. To calibrate the closed-loop control of EES, we found a linear operating range of stimulation amplitudes and systolic blood pressure. The minimum was the amplitude at which the first increase in systolic blood pressure occurred, and the maximum was the last amplitude at which the systolic blood pressure stopped increasing, or when the participant reported abdominal muscle contractions. Each participant remained in a seated position as the amplitude of EES increased by 1 mA every 1 minute to find the minimum (0 mA) and maximum parameters of stimulation (participant dependent). A regression between the average systolic blood pressure per stimulation amplitude resulted in a linear relationship between the systolic blood pressure and the amplitude of stimulation. The closed-loop was tuned manually using the heuristic Zieger-Nichols method^55^. The initial proportional constant was the slope of the regression derived from the calibration phase. The proportional, integral, and derivative constants were tuned over one 70-degree tilt. The same parameters were then used across days to test dynamic changes in the severity of orthostatic challenge. The dynamic changes in tilts consisted of a pseudo-random sequence of 8 changes in tilt angle spanning mild (20 degrees) to severe orthostatic challenge (80 degrees). Each tilt position was held for 60 to 120 seconds. The same sequence was run with continuous EES to enable comparison. An inertial measurement unit-based control algorithm was also developed using the same framework. The angle of the tilt table was derived from changes in 3D acceleration of an inertial measurement unit placed onto the tilt table (Delsys Trigno Avanti, Delsys Inc, Natick, Massachusetts). Data from the inertial measurement unit was recorded at 148 Hz. Accelerometer values in three dimensions were used to compute angular changes. Ranges of tilt positions were mapped to supine, seated, and standing positions, and corresponding amplitudes of EES.

#### Computer simulations

Numerical simulations were conducted using the computational life sciences platform Sim4Life (ZMT Zurich MedTech AG, Switzerland). First, a canonical spinal cord model was developed based on average anatomical measurements of the participant spinal cord anatomies. It was then used to simulate EES exposure throughout the spinal cord anatomy, as well as induced electrophysiological responses^30^. The dosimetric simulations involved finite element discretization using Sim4Life, which resulted in approximately 5 million volume elements (voxels). Each voxel was attributed dielectric properties corresponding to the assigned tissue types^56^ (i.e., spinal cord, spinal roots, cerebrospinal fluid, epidural fat, and vertebral bone). Anisotropic conductivity maps were assigned to the white matter and rootlets, leveraging existing Sim4Life functionalities (either diffusion-simulation- or DTI-imaging-based). By precomputing a suitable basis of electric potential distributions (one per epidural stimulation electrode contact) for the canonical bioelectrical spinal model, the electrical fields resulting from arbitrary EES pulses can rapidly be obtained^30^. For that purpose, the ohmic-current-dominated electro-quasistatic approximation of Maxwell’s equations was solved^57^. In addition to providing an average anatomical geometry, the spine model also encompassed trajectories of axons entering the spinal cord through the dorsal roots and ascending in the dorsal columns. Nerve fiber trajectories were generated inside the rootlets using Sim4Life’s IM-Safe tool, while the fibers in the dorsal column were uniformly distributed. Sim4Life includes functionality to estimate fiber recruitment using the generalized, Green’s function-based activating function, or detailed electrophysiological modeling. For the present work, the classic activating function (proportional to the discrete second derivative of the extracellular potential^57^) was used to assess relative stimulability for a given pulse shape, while electrophysiological simulations with the NEURON simulator incorporated in Sim4Life^58^ permitted conversion to absolute thresholds. For the latter, the predefined diameter-dependent spatially extended non-linear node (SENN) fiber model underlying low-frequency exposure safety standards was used. By assigning fiber diameters according to histology-based probability distributions, the percentage of recruited fibers per root could be estimated and the recruitment level across the different spinal segments and within the dorsal column could be predicted. Activation levels were normalized relative to maximal activation (i.e., recruitment of all dorsal root fibers). Various methodologies have been developed to optimize selective stimulation, including neural network backpropagation and hybrid surrogate modeling (SUMO) - multi-objective genetic algorithm (MOGA) optimization.

#### Electromyographic recordings

Electromyographic (EMG) activity of selected muscles was acquired at a 1,259kHz sample rate using the 16-channel wireless Delsys Trigno sensors (Delsys Inc, Natick, MA, USA) with bipolar surface electrodes. Skin hairs were shaved and abrasive gel was used (Nuprep gel) on the area of interest.

#### Trunk kinetics

Kinematics were captured by Physilog 5 sensors (Mindmaze, Lausanne, Switzerland) that were placed on the vertebral bodies of C3, T1, T6, T10, L1, and S1 of the participants. Each sensor is a stand-alone 10 degree of freedom Micro-Electro-Mechanical System (MEMS) inertial measurement unit includes 3D accelerometer, 3D gyroscope, 3D magnetometer, and a barometric pressure sensor at a sampling frequency of 256 Hz. These inertial measurement units were synchronized via a proprietary radiofrequency protocol.

### Clinical evaluations

#### Orthostatic challenge with tilt-table test

Participants were transferred to a supine position on a table capable of head-up tilt. We applied restraint straps to secure the participant on the knees, on the hips, and on the chest, with the feet stabilized. Resting supine blood pressure was recorded continuously for approximately 5-10 minutes to establish baseline values. Following this period, we tilted the participants upright up to a maximum of 70 degrees while recording hemodynamic values and symptoms of orthostatic tolerance. The time to reach the desired tilt angle from supine was achieved in less than 45 seconds. Participants were tilted until reaching their tolerance threshold or for a maximum duration of 10 minutes. They were asked not to talk during the test except to inform of and grade symptoms. The participant was asked to report any symptoms every 1-3 minutes. The participant was asked to rank the intensity of their symptoms between 1-10, 1 being no symptoms at all, and 10 being maximum feelings of dizziness, lightheadedness^59^, or nausea^19,59^. The participants were instructed to notify the research team if they needed to be returned to the supine position.

#### Blood draws and circulating norepinephrine

Blood samples were withdrawn at baseline after a period of rest in the supine position for 15 minutes and at 2 minutes (T2) after the onset of the tilt. During baseline testing, a total of 3 samples of blood were withdrawn. When testing with EES turned on, an additional sample of blood was withdrawn 5 minutes after the onset of EES in supine position. Blood samples allowed quantification of plasma catecholamines, including plasma norepinephrine, plasma epinephrine, and plasma dopamine. To avoid false positives, the participants were asked to abstain from any beta-blocker, dihydropyridine calcium channel blocker, phenoxybenzamine, anxiolytics, or decongestant medication. To ensure sample quality, prior to and immediately after, collection tubes were kept cold via immersion in crushed ice and kept in foil to reduce exposure to light. Samples were analyzed by the local Hospital Lab services at the University Hospital of Lausanne (Switzerland) and the Foothills Medical Center (Canada). Neuropathic pain prevented blood sampling in P8. Data and tests were not available at the time for P13 and were not part of the clinical protocol for P14.

#### Cerebral Blood Flow

Blood flow was measured in the seated position using Doppler ultrasound imaging equipment (Epiq Elite, Philips, United States). Measurements were conducted both without and with EES. The left and right proximal internal carotid arteries (ICA) and vertebral arteries (VA) were assessed using a 12-3 MHz broadband linear array probe (L12-3, Philips). Additionally, the right and left middle cerebral arteries (MCA) were examined using a transcranial 5-1MHz-pulsed TCD probe.

#### Postprandial blood pressure measurements

On 2 separate days at the same study time point, participants received an identical meal with equal caloric content and their blood pressure was monitored via an arm cuff prior to, during, and for 30 minutes to 1 hour following food intake. This assessment was performed 1 day without EES and 1 day with EES^BP^. For the assessment with EES^BP^, the stimulation was turned on during and after the meal. All points before starting the meal were averaged to determine a baseline value. Available cuff measures between 30 minutes and 60 minutes were averaged to quantify postprandial measures. The delta between baseline and postprandial averages was reported for the conditions without and with EES^BP^. Only participants who reported experiencing post-prandial hypotension were considered in the analysis (P3, P4, P5, P7, P10).

#### Questionnaires

Patient-Reported-Outcome-Measure questionnaires were sent to study participants via the electronic data capture system. Baseline questionnaires were sent once before surgery and then repeated every month after surgery. The following questionnaires are reported: Autonomic Dysreflexia following SCI questionnaire (ADFSCI), Medical Outcomes Study Sleep Scale (MOS-S)^52^, Neurogenic Bowel Dysfunction Score (NBDS), and Usability SUS^41^.

#### Spirometer measurements

The study participants were asked to perform a systematic set of breathing and cough tasks. Volume, flow and time data were recorded using the Spirometer (Spirobank II Smart Spirometer). During the coughing maneuver, EES^COUGHING^ was precisely timed with the expiratory phase after releasing the glottis blockage. All participants from the HemON study performed the respiratory and coughing assessments at 6 months without stimulation and with EES^TRUNK^ except P12 that could not perform the test at this timepoint.

#### Canadian Occupational Performance Measure (COPM)

The COPM was employed to assess the functional performance and perceived satisfaction with activities of daily living of each participant. The COPM was administered by trained occupational therapists to follow standard procedures. At baseline, participants identified up to five activities they perceived as important but difficult to perform and rated their perceived performance and satisfaction on a 10-point scale. The general scores were calculated by simply averaging the performance and satisfaction ratings across activities. The same activities were then rated again by the participants 3 and 6 months after implantation.

#### Spasticity assessment

Participants’ upper and lower limb spasticity was assessed using the Modified Ashworth Scale (MAS) in 2 conditions: without and with EES^SPASTICITY^. Participants were either asked to use their standard-of-care medication or to not use their standard-of-care medication the morning prior to the assessment. Participants were asked to maintain the stimulation off in the mornings prior to the assessment, which were conducted in the late morning.

#### Rehabilitation outcomes

In the HemON trial, physiotherapists and occupational therapists kept track of the 1-month rehabilitation participants’ sessions and documented whether the different activities were performed without or with EES through a standardized form (Mapping of Rehab Training - MART).

#### Muscle sympathetic nerve activity

In the HEMO trial, efferent postganglionic Muscle sympathetic nerve activity (MSNA) was acquired from the left common peroneal (fibular) nerve using microneurography^1,60,61^. The location of the common fibular nerve was located using palpation. A 2-MΩ tungsten microelectrode (Frederick Haer and Co, Bowdoin) was then inserted percutaneously alongside a subdermal low impedance reference electrode 2cm adjacent. The amplifier and head stage (NeuroAmpEx, AD Instruments) were also grounded to the participant with a surface electrode on the patella. The recording electrode was adjusted until entering the nerve. Crossing into the nerve and into different nerve bundles was identified audibly by the microneurographer. Once in the nerve, the recording microelectrode was confirmed to be near nerve fibers directed towards skeletal muscle by auditory feedback during tapping/palpation of the tibialis anterior/peroneal muscles, which has been shown previously to be indicative of microelectrode proximity to efferent postganglionic muscle sympathetic nerves1. If auditory feedback was present during light stroking of skin on the dorsal foot/lower leg, this indicated proximity to post-ganglionic skin sympathetic fibers. The search was stopped when the electrode was in proximity to muscle sympathetic fibers.

### Data processing

#### Intraoperative blood pressure data

During surgery (see **Neurosurgical intervention**), changes in blood pressure were recorded in response to different locations of EES using the arterial line (see **Post-operative hemodynamic monitoring**). Change in blood pressure or heart rate was defined as the difference in the average of 30-second windows before the start of the stimulation and at the end of the stimulation. For cross-subject analysis, the lower thoracic and lumbosacral stimulation locations in each subject were calculated by their relative distance to the center of the hemodynamic hotspot (T11) normalized by their respective spinal length (measured from T9 to the conus). Due to surgery time limits, P3 blood pressure mapping was not performed.

#### Postoperative blood pressure data

During a tilt-table test (See **Orthostatic challenge**), changes in blood pressure were recorded without stimulation or in response to different types of stimulation (continuous or closed-loop stimulation) using the Finometer (see **Post-operative hemodynamic monitoring**). Change in blood pressure or heart rate was defined as the difference in the average of a 60-second window before the start of the tilt and a 20-second window at 3 minutes of the challenge. If the participant could not tolerate at least 3 minutes of the test due to low blood pressure or other symptoms, an average of a 20-second window before the end of the tilt was used. All measurements in seated position were measured with stimulation on for 3 – 5 minutes. Change in blood pressure, or heart rate, was defined as the difference in the average of a 20-second window before the start of EES and the average of a 20-second window at 3 minutes, prior to stop stimulation. All signals were smoothed over a 10-second window for illustration. The same processing was used for post-operative, day 1 quantification. Data from continuous blood pressure monitoring was excluded if technical issues with the Finometer were observed including loss of the plethysmographic signal, incorrect calibration as measured by comparing continuous measuring to standard measurements from a brachial cuff, spasms, or due to study protocol deviations or adverse events.

#### Stimulation paradigms

The efficacy of continuous, closed-loop, and inertial measurement unit-based control of EES was measured by: 1) target error, defined as the error of systolic blood pressure with respect to a user-defined target on the systolic blood pressure, 2) error variability, defined as the standard deviation of the target error, to measure stability of the stimulation during dynamic orthostatic challenges. Both metrics were calculated from 1 minute before the dynamic challenges to 1 minute after the end of the challenge.

#### Cerebral Blood Flow (CBF) Calculations

Peak systolic and end-diastolic flow velocities were obtained from the Doppler spectrum analysis over three consecutive cycles in all arteries. Mean velocity was derived through the integration of the Doppler curve for each corresponding pair, with the mean value calculated across the three cycles. Arterial diameters were determined in B-mode for the left and right ICAs and VAs. The flow rate (Q) in each vessel, measured in milliliters per minute, was calculated using Equation 1, assuming a parabolic velocity profile. The total experimental cerebral blood flow (CBF_exp) was determined by summing the flow of the left and right ICAs and VAs, according to Equation 2^62^.

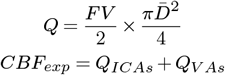

#### Autonomic Dysfunction Following Spinal Cord Injury (ADFSCI)

The ADFSCI is a 24-item questionnaire divided into four sections: demographics, medication, Autonomic Dysreflexia (AD), and hypotension. The hypotension section consists of 7 items. Each item employs a 5-point scale to measure the frequency and severity of symptoms related to hypotension, including headaches, goosebumps, dizziness, and light-headedness, across different situational contexts. The hypotension score corresponds to the sum of all the hypotension items. Participants were categorized as experiencing symptoms if the item score was higher or equal to 2, else they were categorized as not experiencing symptoms.

#### Analysis of kinematics

The acceleration and angular velocity collected by the Physilog 5 sensors were passed through a datafusion algorithm^63^ to extract sensor orientation with respect to the Earth in quaternion representation. Roll, pitch, and yaw angles were then calculated by using one of two representations (zyx or zyz), depending on the sensor placement with respect to its axis with the greatest motion, such that there would be no discontinuities in the pitch. The ratio of change in back curvature when turning the stimulation on is approximated as the ratio of the angles between the T6 and T10 vertebrae in the sagittal plane with and without stimulation.

#### Muscle sympathetic nerve activity

The MSNA signal was sampled at 20kHz, amplified (20,000×), and band-pass filtered (0.3–2.0 kHz) to obtain muscle sympathetic spike activity, and then rectified and integrated (0.1-s time constant) to obtain a multi-unit (mean voltage) neurogram (Labchart version 8, AD Instruments). After acquiring a stable recording site, a 3-min baseline commenced followed by step changes in epidural stimulation intensity for 1-2 minutes at each stage. Muscle sympathetic action potential spikes were identified as waveforms that matched a triphasic morphology with the main phase being negative. The negative deflections were only assessed if they were larger than 4.5x the standard deviation of the neurogram assessed during baseline (stimulator off).

#### Splanchnic impedance methods

In the HEMO trial, Bioelectrical Impedance (BEI) of the lower torso (abdominal cavity) was used to estimate fluid shifts within the splanchnic vascular regions during tilt-table testing^64^. BEI is measured by driving a small electrical current (0.7 mA with a frequency of 37 kHz) between electrodes and measuring the impedance with voltage-sensing electrodes on each end of the body segment of interest (Model 2994D/4 THRIM, UFI). For the lower torso, surface electrodes were placed at approximately the left 6th rib and around the left inguinal ligament. The impedance, measured in ohms, was low pass filtered at 0.5 Hz. Since body segment volume is inversely proportional to electrical resistance (V *∝* 1/R) such that an increase in resistance reflects a loss of body fluid, the impedance values were used to estimate body segment volume using the following equation: 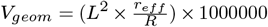, where L is the segment length in meters, r_eff_ is the effective resistance 1.0 Wm, and R is the segment impedance in W.

#### Statistics

All data are reported as mean values and individual data points. No statistical methods were used to predetermine sample sizes, but our sample sizes are similar to those reported in previous publications^65^. All statistical analysis was performed in R using the base package ‘stats’, with primary implementation through the ‘tidyverse’ and ‘broom’ packages. Tests used included one or two-tailed paired or independent samples Student’s t-tests, one-way ANOVA for evaluations with more than two conditions, and one- or two-way repeated-measures ANOVA for assessments, when data were distributed normally, tested using a Shapiro–Wilk test. Post hoc Tukey tests were applied when appropriate. For regressions, mixed model linear regression was used in cases of multiple observations, or else standard linear modeling. In cases where group size was equal to or less than three null hypothesis testing was not completed. The significance level was set as P < 0.05. Exclusions of data are noted in the relevant methods sections. Unless stated otherwise, experiments were not randomized, and the investigators were not blinded to allocation during experiments and outcome assessment.

## Data Availability

Software routines developed for the data analysis will be made available upon reasonable request to the corresponding authors.

## Acknowledgments

The authors would like to thank the study participants for their commitment and trust. All participants gave their informed consent for publication of their images. The authors would also like to thank M. Boulingre, J. Bourgeot, K. Charlong, M. Kameni-Hekam, C. Magrini, L. Mathews, L. McCracken, C. Parcheminey, M. Pedone, C. Perrin, M. Picek, K. Plesniar, C. Schiff, M. Smith and K. Tourigny for various contributions including data collection, support in patient logistics, patient care and regulatory affairs; I. Benrikane, A. Di Russo, and Y. Qin for their contributions in data analysis.

## Funding

This work was financially supported by: Eurostars project E!113969 PREP2GO, DARPA subaward A21-0795-S001 (P.O. 1083297), Eurostars E!1748 IMPULSE, Medtronic (ERP-2020-12543), Onward Medical, PHRT-279, the Defitech foundation, CIHR, Cumming Medical Research Fund, Edith Rodie Estate, Hotchkiss Brain Institute, Libin Cardiovascular Institute.

& **Clinical Study Team** Lorraine Aviolat^7^, Nadia Bérard^22^, Julie Brancato^22^, Krystel Bruyère^7^, Rebecca Charbonneau^1,2^, Chris Drummond-Main^2^, Sean Dukelow^1,2^, Grégoire Eberle^7^, John Gaudet^24^, Natacha Herrmann^7^, Julie Hervé^5,6,7^, Jamie Johnston^2^, Valentin Lupi^7^, John-Paul Miroz^5,6,7^, Aurélie Paley^5,6,7^, Julie Pillonel^7^, Carole Poulin-Kella^7^, Mélanie Ramirez^7^, Katrien Van Den Keybus^7^, Camille Varescon^5,6,7^, Molywan Vat^5,6,7^, Xavier Vo Pham^7^, Laurence Wenger^7^ and Patrick Koomen^9^

&& **Onward Team** Francesco Acquati^6,10^, Pierre Bessot^10^, Julien Dedelley^10^, Vincent Delattre^10^, Anahita Kyani^10^, Hendrik Lambert^10^, Sebastien Morand^10^, Chloé Picq^10^, Jared Pradarelli^10^, Francesca Stradolini^10^ and Rosanne van Dijsseldonk^10^

## Author contributions

**Writing of the manuscript** G.C. and A.Phillips., wrote the manuscript with J.W.S., L.Asboth., J.Bloch., N.H., R.D., A.P.G., and S.H., and all the authors contributed to editing the manuscript.

**Conducting therapy sessions or clinical evaluations** A.P.G., A.I., K.K., A. Phillips, R.M., J. Lee, J. Rimok, L.Asboth, M.B., J.B, L.B., K.D., N.H., N.I., J.Ledoux, E.Pralong, C.V., G.W., G.C., L.D.H., R.D.,, S.H., N.M., N.L., A.N., F.A., M.D., K.G., C.Picq, J.Pradarelli, M.Rieger, R.V., P.D., D.S, E.A., L.Aviolat, O.B., T.B., F.B., M.B., J.Bloch, J.Brancato, K.B., S. Carda, K.D., N.Hermann, J.H., N.I., J.Ledoux, V.L., J.Miroz, A.Paley, J.Pillonel, M.Ramirez, M.V., X.V., L.W., G.W., J.W.S., P.K. and E.B.

**Data analysis** A.P.G., A.I., K.K., R.M., P.D., D.S., J. Lee, J. Rimok, A. Phillips, L..Asboth, T.B., F.B., N.H., J.Ledoux, T.M., L.D.H., R.D., G.D., S.H., N.M., J.W.S., A.N., F.A., M.D., K.G., A.K., S.Morand, F.S., R.V. and E.B.

**Hardware/software development** A.P.G., L.Asboth, N.H., C.H., T.M., C.S., G.C., R.D., G.D., S.H., N.M., A.R., J. Garcia, N. Kuster, E.N., T.N., M.C., M.D., V.D., J.D., L.D., Y.D., D.G., K.G., H.L., J.Murphy, E.Paoles, J.F., T.V., E.M., S.Mandija and C.B.

**Regulatory affairs management** K.K., A.Phillips, L.Asboth, J.Bloch, L.B., N.H., G.C., L.D.H., R.D., A.P.G., I.V., P.B., M.D., L.D., K.G., C.J., H.L., C.Picq, M.Rieger, E.R., R.V. and A.W.

**Illustration preparation** L.Asboth, N.H., G.C., R.D., A. Phillips, K.K., A.P.G., M.G., S.H., F.M., J.Ravier and J.W.S.

**Funding acquisition and supervision** G.C., J.B. and A.Phillips.

**Team Supervision** K.K., A.Phillips, L.Asboth, J.Bloch, G.E., L.H., T.M., P.S., K.V., G.W., G.C., R.D., J.W.S., N. Kuster, E.N., N.Keijsers, N.L., I.V., M.D., H.L., J.Murphy, E.R., A.W., S.Mandija and C.B.

**Surgery** J.Bloch, S.Casha, F.G., E.K. and J.W.S.

## Code availability

Codes are available upon reasonable requests to G.C.

## Additional information

Correspondence and requests for materials should be addressed to G.C., J.Bloch and A.Phillips.

---

**Supplementary Fig. 1.**
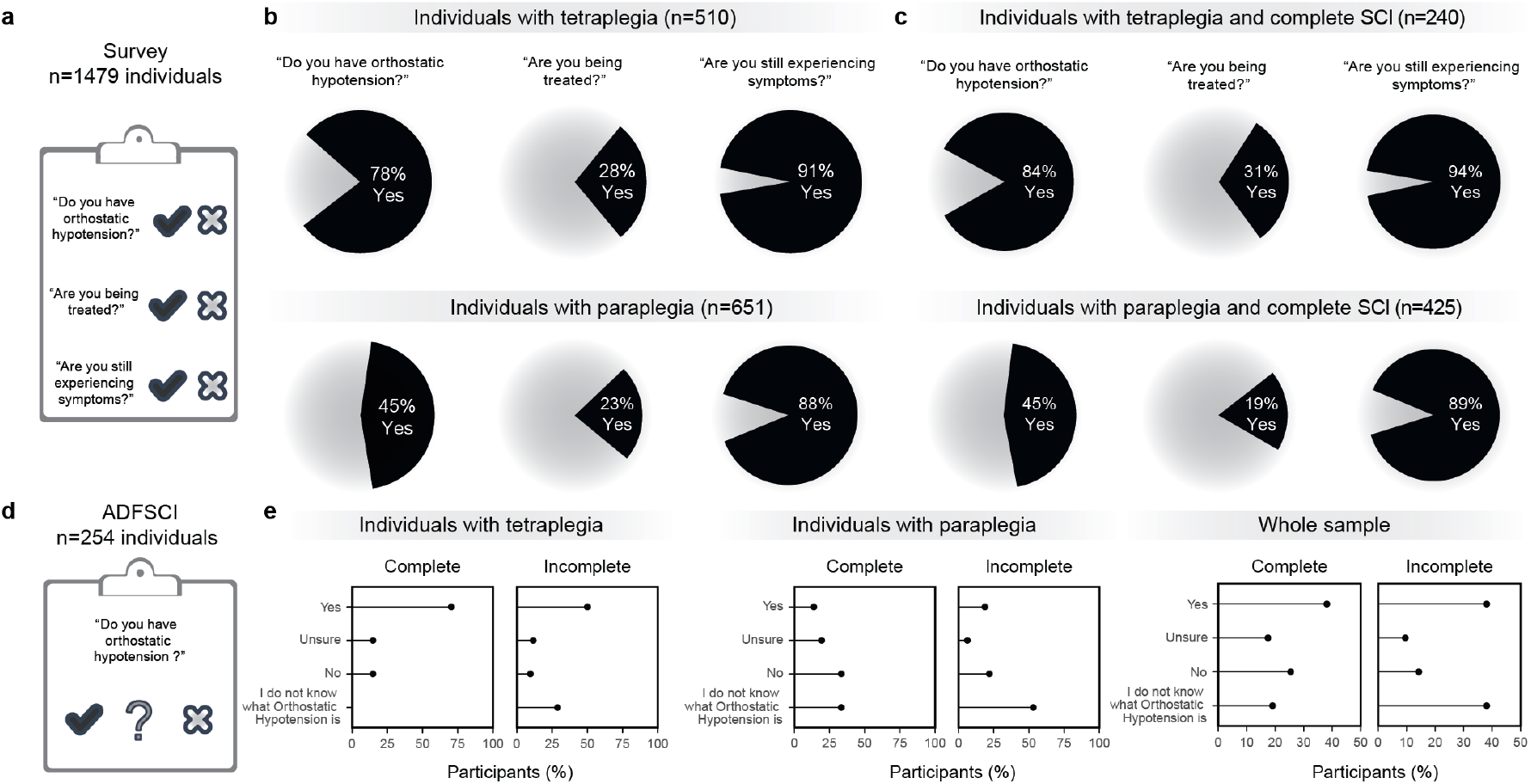
Orthostatic hypotension is a medically refractory condition in people with SCI. **a**, Survey on the presence of orthostatic hypotension and efficacy of its management conducted in a cohort of 1479 individuals with SCI as part of the Spinal Cord Injury Community Survey (SCICS). **b**, Prevalence of orthostatic hypotension and efficacy of its management quantified from the SCICS survey (n = 1161 of 1479 valid responses, of which 510 individuals with tetraplegia and 651 with paraplegia). **c**, Prevalence of orthostatic hypotension and efficacy of its management in individuals with functionally complete SCI quantified from the SCICS survey (n = 665 individuals with complete SCI, of which n= 240 individuals with tetraplegia and n= 425 with paraplegia). **d**, Survey on the presence of orthostatic hypotension conducted in 254 individuals with SCI, acquired with the Autonomic Dysfunction Following Spinal Cord Injury questionnaire (ADFSCI). **e**, Percentage of individuals who report orthostatic hypotension, segregated by the severity of the SCI in people with tetraplegia (n = 79), paraplegia (n = 68), and for the entire sample (n = 147).

**Supplementary Fig. 2.**
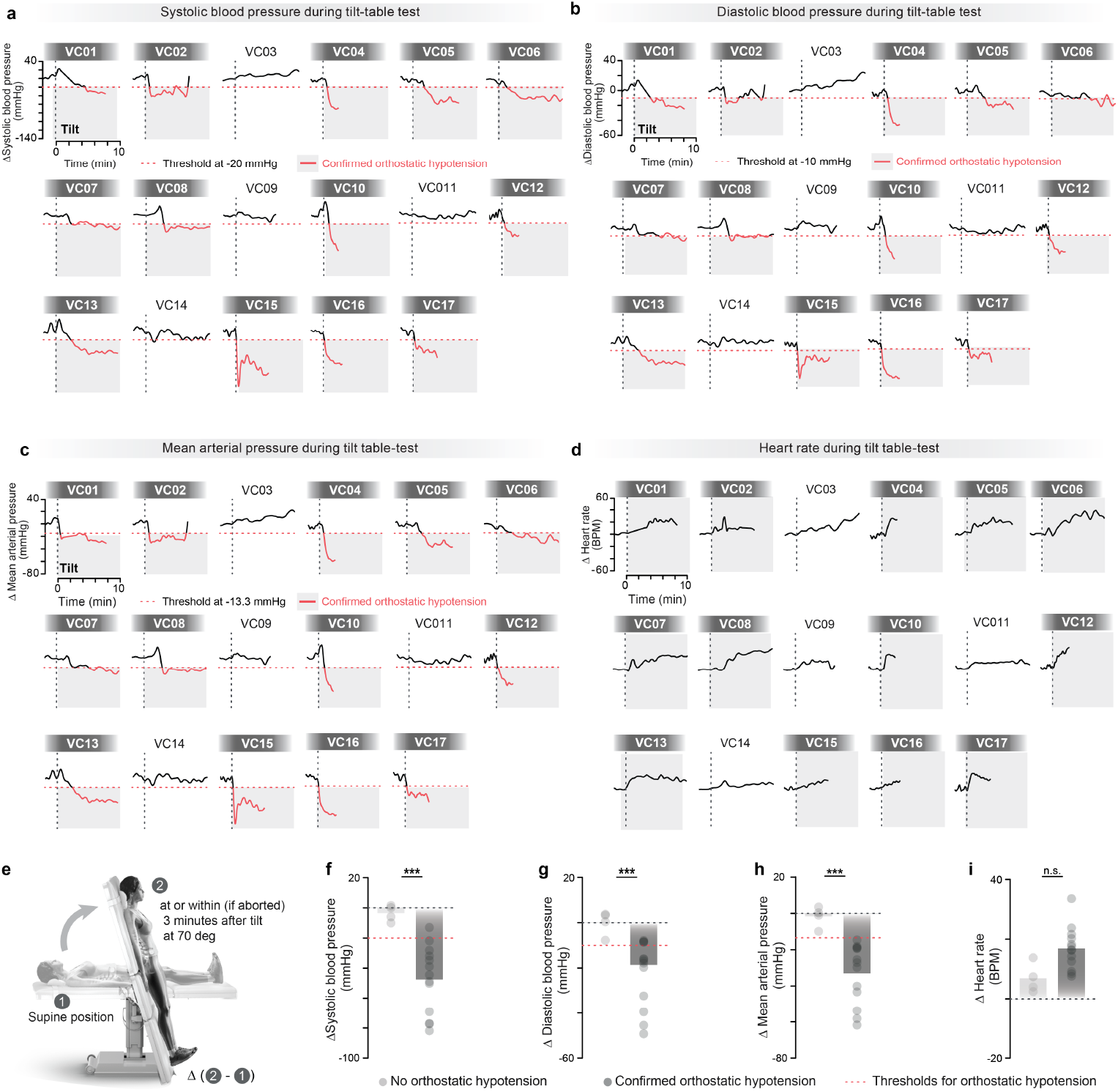
Validation of ADFSCI survey in a random cohort of individuals with SCI. **a**, Change in systolic blood pressure in response to an orthostatic challenge from a random sample of individuals who had filled out the Autonomic Dysfunction Following Spinal Cord Injury. The threshold crossing for confirmed orthostatic hypotension (red) was based on criteria established by the American Autonomic Society and the American Academy of Neurology^29^. **b**, As in **a**, for diastolic blood pressure. **c**, As in **a**, for mean arterial pressure. **d**, As in **a**, for heart rate. **e**, Scheme of the tilt table test described in **a-d. f**, Average change in systolic blood pressure within 3 minutes of a 10-minute tilt-table test (independent samples two-tailed t-test; t-value = 6.44, p = 1.19e-05). Each dot reports quantifications from a single participant who did not present with orthostatic hypotension (light grey) or who presented confirmed orthostatic hypotension (dark grey). **g**, Same as in **f**, for diastolic blood pressure (independent samples two-tailed t-test; t = 4.51; p = 4.78e-04). **h**, Same as in **f**, for mean arterial pressure (independent samples two-tailed t-test; t = 5.78; p = 4.27e-05). **i**, Same as in **f**, for heart rate (independent samples two-tailed t-test; t = -3.13; p = 1.60e-02).

**Supplementary Fig. 3.**
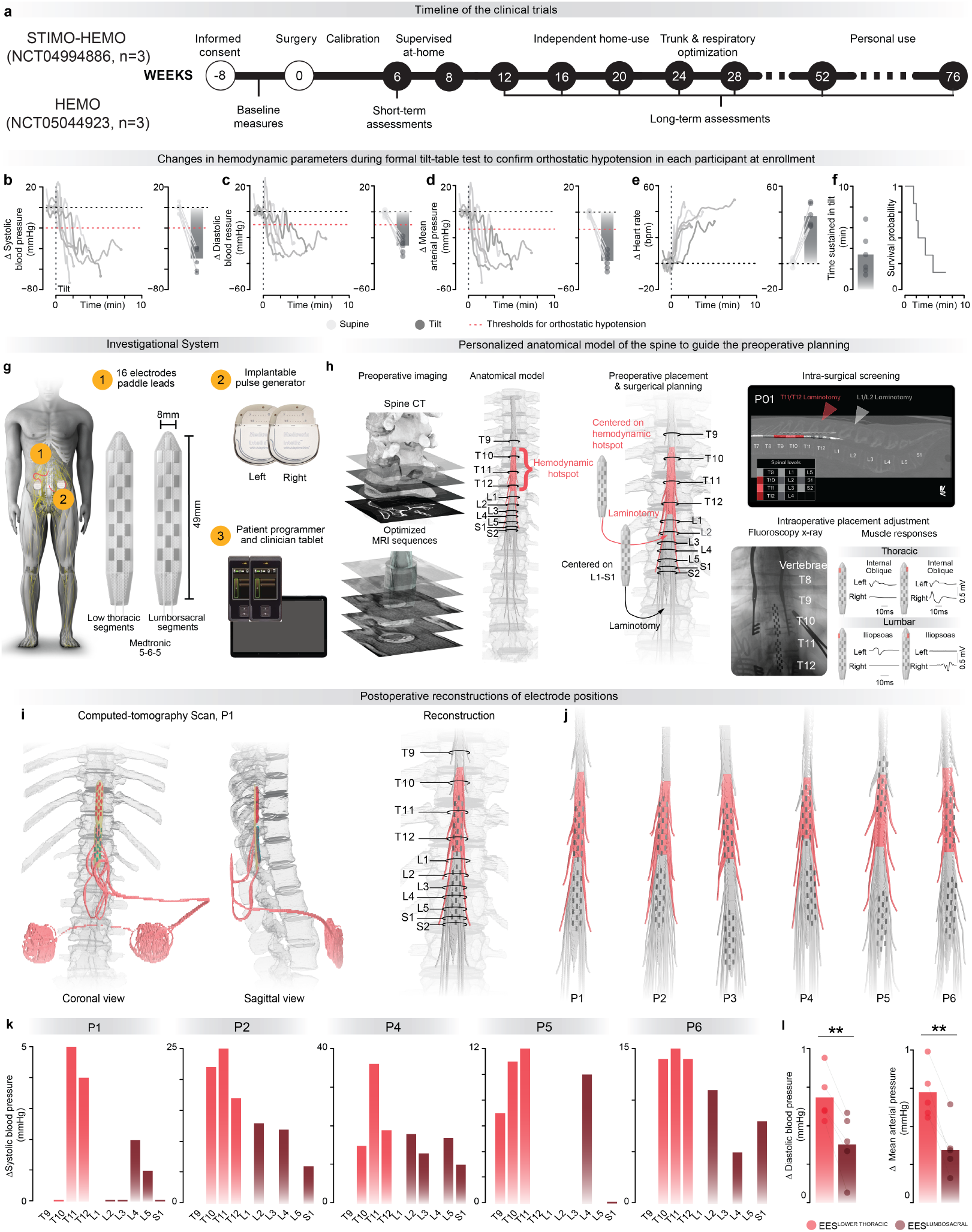
Mapping of the optimal location to modulate blood pressure with EES. **a**, Timeline of the clinical trial 1 (STIMO-HEMO) conducted in Lausanne, Switzerland and trial 2 (HEMO) in Calgary, Canada. **b**, Changes in systolic blood pressure during a formal 10-minute tilt-table test to verify that the six participants met the criteria for confirmed orthostatic hypotension. The bar graph reports the average drop in systolic blood pressure at or within (if the test had to be aborted before) 3 minutes of verticalization at a 70-degree tilt position. **c**, As in **b**, for diastolic blood pressure. **d**, As in **b**, for mean arterial pressure. **e**, As in **b**, for heart rate. **f**, Bar graph reporting the average duration of the tilt-table test shown in panels **b-e**. Each point denotes a single tilt for each of the six participants. The corresponding Kaplan-Meier plot of exposure status to time sustained in tilt is shown on the right. **g**, Schematic illustration of the investigational device employed to deliver EES in both trials, which includes 2 Medtronic Intellis Implantable Pulse Generators (IPG), 2 Medtronic Specify Surescan 5-6-5, and Medtronic clinical and patient programmers for home use. **h**, A personalized anatomical model of the spine is elaborated for each participant based on high-resolution magnetic resonance imaging and computed tomography. This model guides the determination of the desired lead placement and spinal entry levels (location of laminectomy). The final location of the leads is optimized intraoperatively based on the monitoring of electromyographic (EMG) signals from key trunk and leg muscles, as exemplified at the bottom right. **i**, The locations of each electrode are captured with postoperative computed imaging, which are then injected in the personalized anatomical model to visualize the final placement of both leads. **j**, Final location of the electrodes for the six participants of both trials. **k**, Changes in systolic blood pressure in response to EES applied over the segments of the lower thoracic and lumbosacral spinal cord during the intraoperative mapping for each participant. The location of the segments was determined postoperatively. Due to time constraints and electrode locations, only a subset of segments could be tested for each participant. **l**, Bar plots reporting the average change in diastolic blood pressure (*left*) (n = 5, paired samples two-tailed t-test; t = 6.17; p = 3.50e-03) and mean arterial pressure (*right*) (n = 5, paired samples two-tailed t-test; t = 7.55; p = 1.70e-03) in response to EES applied over the lower thoracic versus lumbosacral spinal segments during the intraoperative mapping.

**Supplementary Fig. 4.**
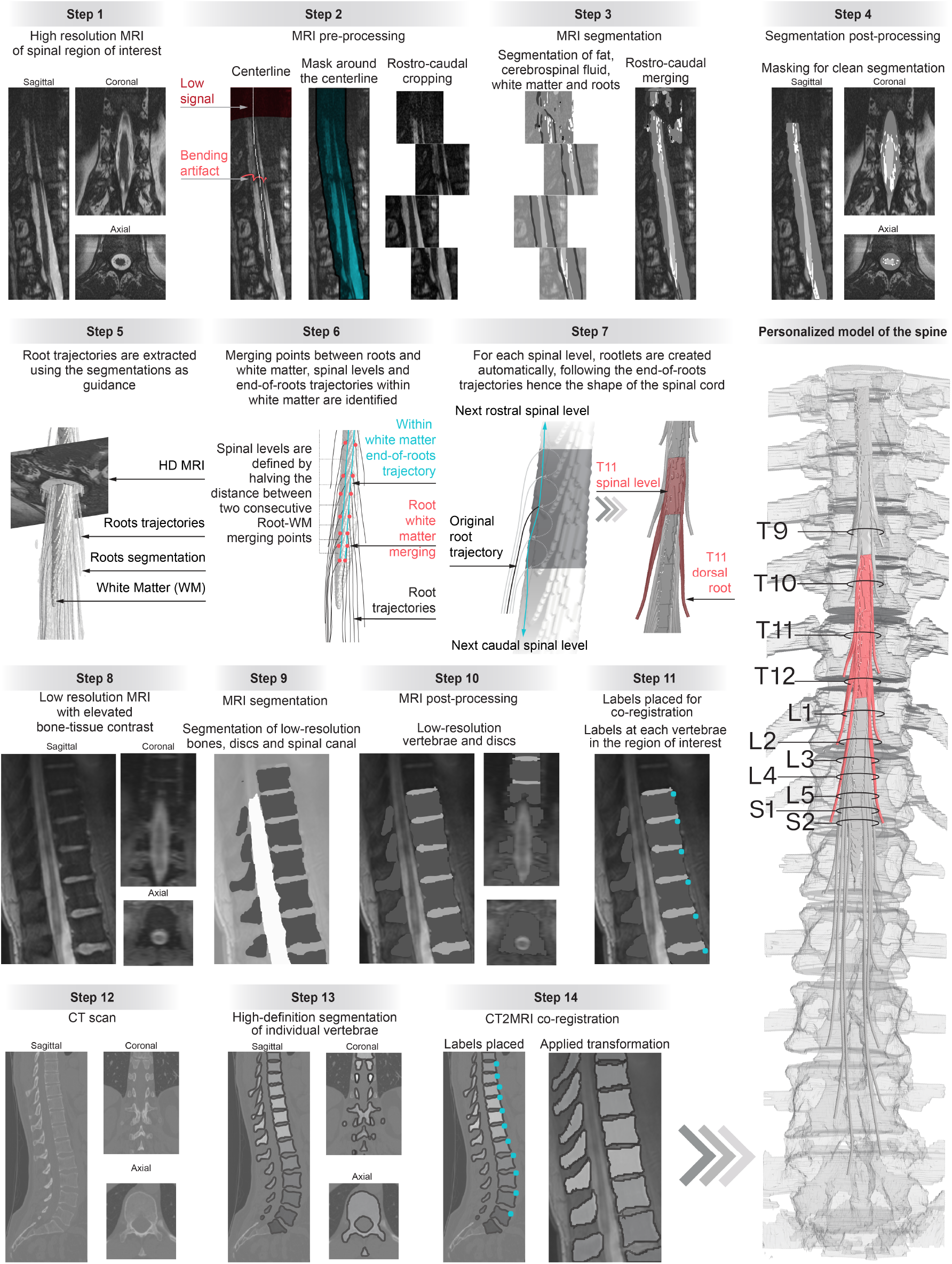
Personalized anatomical models of the spine. **Step 1**, Acquisition of high-definition magnetic resonance imaging (MRI) images of the relevant region of the spine with MRI sequences that have been optimized to augment the contrast between the dorsal root and cerebrospinal fluid. **Step 2**, Accurate detection of the centerline of the spinal cord, even in areas with low signal or artifacts. Creation of a 35 mm cylindric mask around the centerline and cropping of the image into boxes along the rostrocaudal axis. **Step 3**, A neural network trained on segmented data from 12 healthy volunteers is applied on the MRI acquisitions to detect the epidural fat, cerebrospinal fluid, white matter, and spinal root tissues. The segmented boxes are then merged, and manual corrections are performed. **Step 4**, A mask from the segmented epidural fat is used to refine the segmentation, with an option for manual adjustments. **Step 5**, The contour of ventral and dorsal roots are traced using the MRI and segmented tissues as guides, which enables the labeling of roots as they exit the vertebrae. **Step 6**, The trajectories of the spinal roots are trimmed within white matter tissues to 1 mm. These endpoints are used to determine the spinal segments and trajectories of the roots. This procedure is robust to the presence of scoliosis. **Step 7**, The trajectory of each spinal root is subdivided into 5 rootlets in order to generate anatomically realistic tridimensional models of the rootlets. **Step 8**, Acquisition of low-resolution MRIs with enhanced bone-tissue contrast. **Step 9**, Segmentation of bones, discs, and spinal canal using a neural network trained on low-quality data. **Step 10**, Removal of incomplete vertebrae/discs, discarding spinal canal segmentation. **Step 11**, Labeling of intervertebral discs for vertebrae identification to facilitate the coregistration of CT and MRI acquisitions. **Step 12**, Acquisition of high-definition computerized tomography scan. **Step 13**, Segmentation of individual vertebrae from the computerized tomography scan. **Step 14**, Alignment of computerized tomography and MRI data using labeled vertebrae and transformation with a two-step function. **Step 15**, Personalized anatomical model of the spine is generated based on the surfaces of the segmented tissues extracted from the MRI images. Preoperative and postoperative computerized tomography scans are used to update the model with the implanted system.

**Supplementary Fig. 5.**
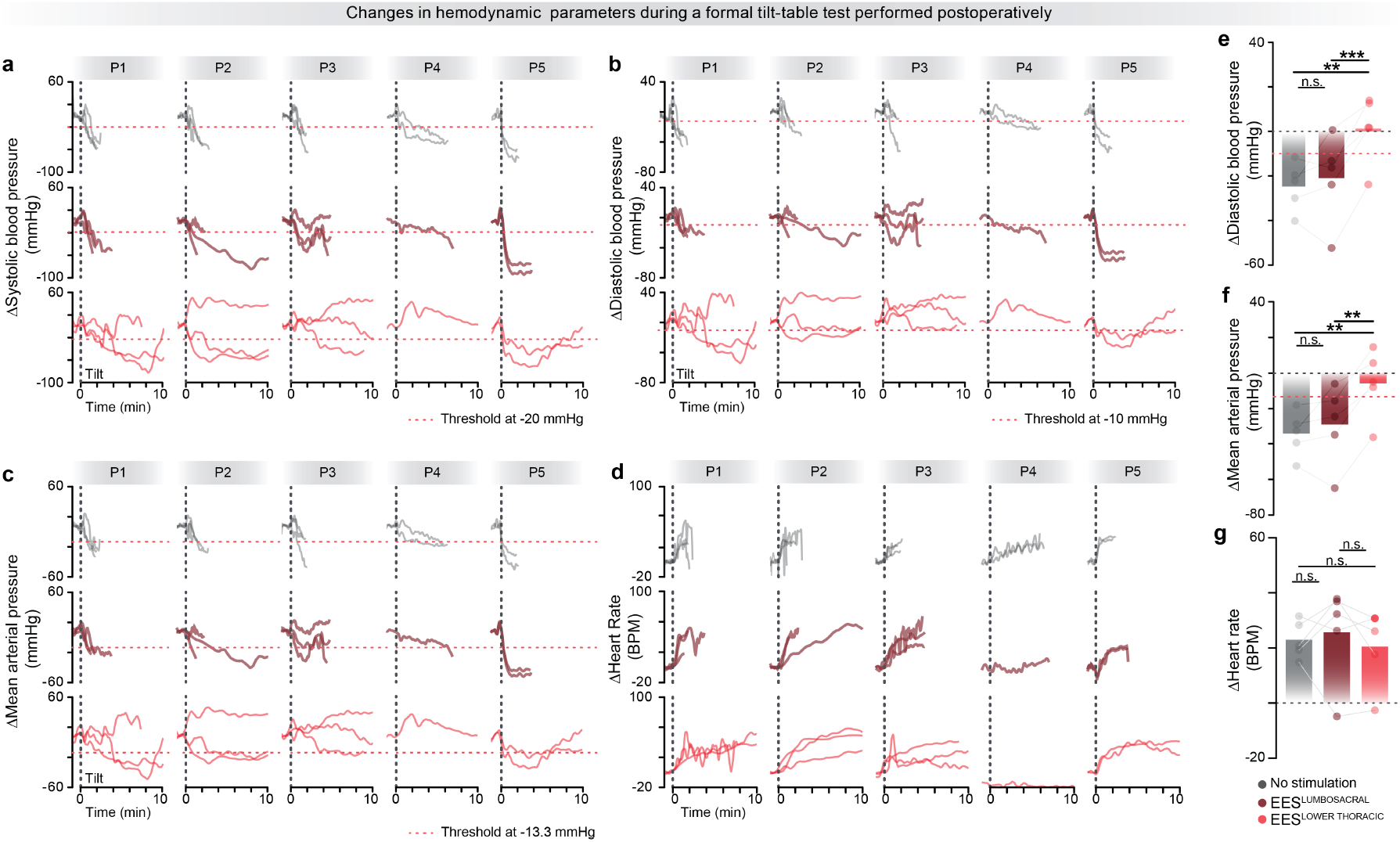
The hemodynamic hotspot to regulate blood pressure with EES is located in the low thoracic spinal cord, not the lumbosacral spinal cord. **a**, Changes in systolic blood pressure during a formal tilt-table test performed without EES, or with EES applied over the lower thoracic segments or the lumbosacral segments during postoperative evaluations. Each trace corresponds to a representative test under each experimental condition for the 5 participants. **b**, As in **a**,, for diastolic blood pressure. **c**, As in **a**,, for mean arterial pressure. **d**, As in **a**,, for heart rate. **e**, Bar graphs reporting the average drop in diastolic blood pressure at or within (if aborted) 3 minutes during a formal tilt-table test without EES, or with EES applied over the lower thoracic segments or the lumbosacral segments. 3 tilts were obtained for P1-P3 in each condition; 2 tilts were obtained for P4 without EES and 1 tilt in each condition with EES; 2 tilts were obtained for P5 in each condition (n = 5, repeated measures, two-tailed ANOVA with Tukey’s HSD; f = 19.6; p = 8.3e-04). **f**, As in **e**,, for mean arterial pressure (n=5, repeated measures, two-tailed ANOVA with Tukey’s HSD; f = 19.27; p = 8.7e-04). **g**, As in **e**, for heart rate (n=5, repeated measures, two-tailed ANOVA with Tukey’s HSD; f = 0.29; p = 0.76).

**Supplementary Fig. 6.**
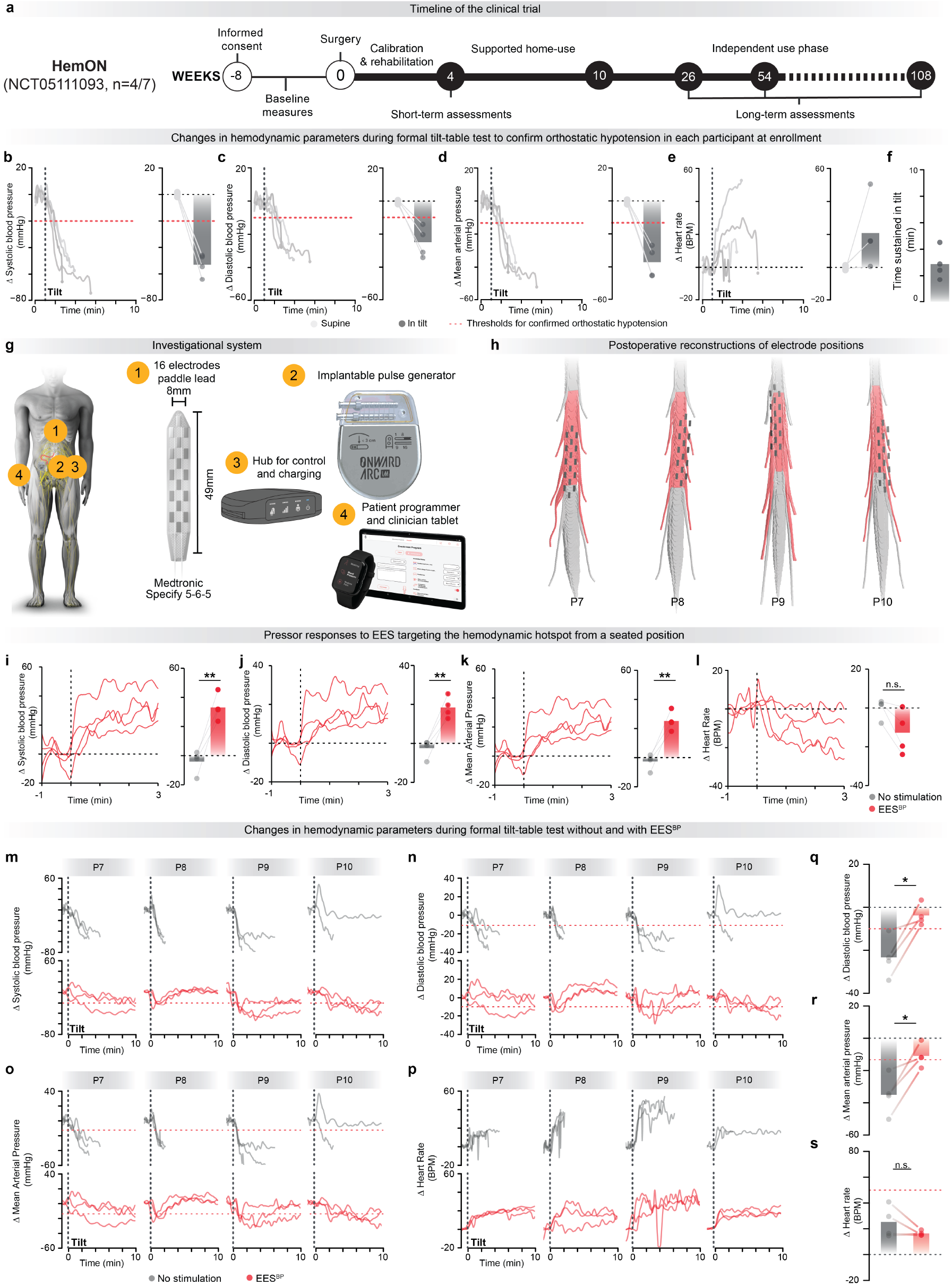
Validation of the purpose-built neurostimulation platform. **a**, Timeline of clinical trial 3 (HemON) conducted in Lausanne, Switzerland in 4 participants. **b**, Changes in systolic blood pressure during a formal 10-minute tilt-table test to verify that the four participants met the criteria for confirmed orthostatic hypotension (red dotted line). The bar graph reports the average drop in systolic blood pressure at or within (if the test had to be aborted before) 3 minutes of verticalization at a 70-degree tilt position. **c**, As in **b**, for diastolic blood pressure. **d**, As in **b**, for mean arterial pressure. **e**, As in **b**, for heart rate. **f**, Bar graph reporting the average duration of the tilt-table test shown in panels **b-e**. Each point denotes a single tilt for each of the 4 participants. **g**, Schematic illustration of the investigational device employed to deliver EES, including the ONWARD ARC^IM^ Implantable Pulse Generator (IPG), the Medtronic Specify Surescan 5-6-5, the ARC^IM^ Hub for therapy control and IPG charging, and the ONWARD ARC^IM^ clinician and personal programmer for home use. **h**, Final location of the electrodes with respect to the hemodynamic hotspot (red) for the 4 participants. **i**, Changes in systolic blood pressure when delivering EES targeting the hemodynamic hotspot (EES^BP^) in a seated position for each of the 4 participants. The bar graphs report average changes in systolic blood pressure (paired, t-test; t = 6.75, p = 6.65e-03). **j**, As in i, for diastolic blood pressure (paired, t-test; t = 7.56, p = 4.81e-03). **k**, As in i, for mean arterial pressure (paired, t-test; t = 7.12, p = 5.70e-03). **l**, As in i, for heart rate (paired, t-test; t = 3.10, p = 5.31e-02). **m**, Changes in systolic blood pressure without (grey) or with EES^BP^ (red) during a formal tilt-table test. **n**, As in **m**, for diastolic blood pressure. **o**, As in **m**, for mean arterial pressure. **p**, As in **m**, for heart rate. **q**, Bar plots reporting the average change in diastolic blood pressure during a formal tilt-table test without and with EES^BP^ (paired, t-test; t = 3.84; p = 3.1e-02). **r**, As in **q**, for mean arterial pressure (paired, t-test; t = 4.00; p = 2.8e-02). **s**, As in **q**, for heart rate (paired, t-test; t = 1.63; p = 0.20).

**Supplementary Fig. 7.**
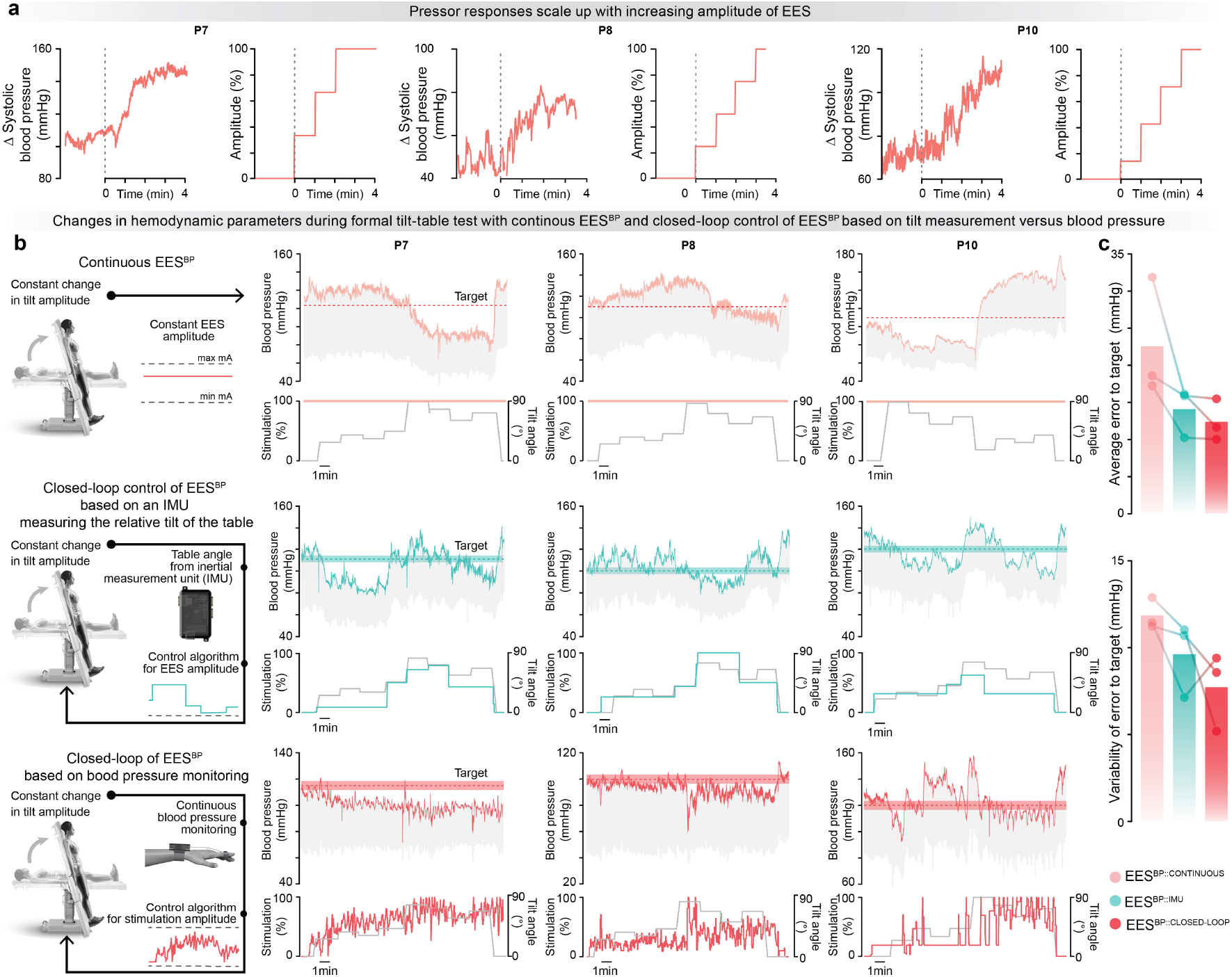
Closed-loop control of EES to stabilize blood pressure despite constant-changing orthostatic challenges. **a**, Changes in systolic blood pressure in response to graded increase in the amplitude of EES by 1 mA increment every minute. **b**, Schematic view of the three different paradigms used to control EES^BP^ during dynamic changes of the angle of the tilt-table (*left*). Changes in blood pressure while EES^BP^ is delivered continuously (*top*), with closed-loop control of EES^BP^ based on the amplitude of the tilt measured with an IMU (*middle*), and with closed-loop control of EES^BP^ based on continuous monitoring of systolic blood pressure (*bottom*) for the 3 participants. **c**, Bar plots reporting the average error to target (*left*) and variability of the error to target (*right*) for the 3 experimental conditions described in (**b**). The target was based on a predefined level of systolic blood pressure.

**Supplementary Fig. 8.**
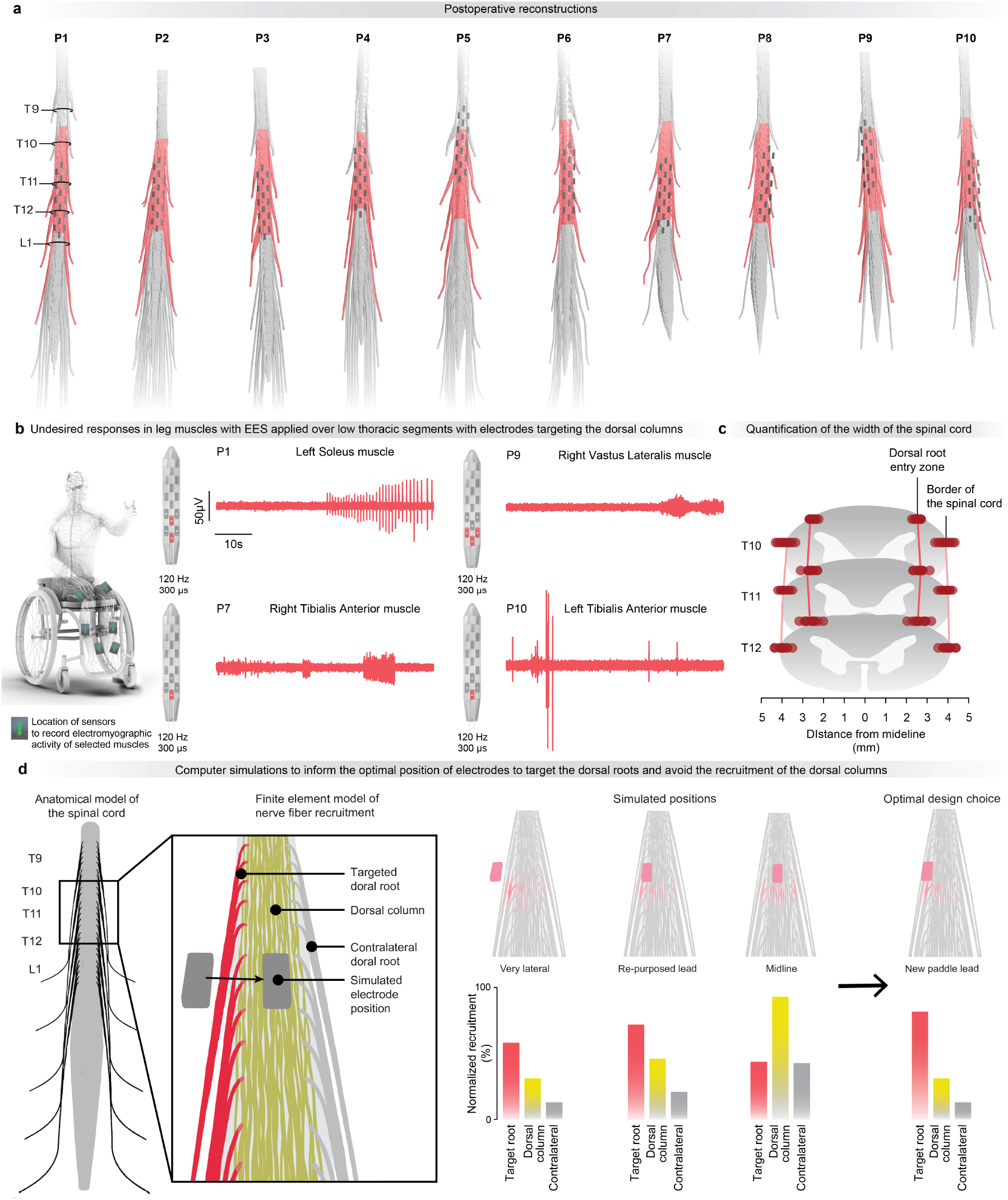
Informed design of the purpose-built paddle lead. **a**, Personalized anatomical models of the spinal cord updated with the final placement of the paddle lead for the 10 participants who were implanted with the Medtronic Surescan Specify 5-6-5 paddle lead (Supplementary Table 2). The hemodynamic hotspot is represented in red, thus informing the relative coverage of this hotspot with this specific lead. **b**, Examples of undesired responses in leg muscles when EES was applied with over electrode configurations that likely recruited the dorsal columns, and thus elicited muscle activity via antidromic afferent volleys evoked on ascending afferent fibers. **c**, Quantification of the location of the dorsal root entry zones and of the width of the spinal cord based on high-resolution magnetic resonance imaging acquired in the 10 participants shown in (**a**). **d**, Computer simulations to predict the relative recruitment of the targeted dorsal root, dorsal columns, and contralateral dorsal root based on the location of the electrode along the lateral directions. A canonical model of the lower thoracic spinal cord was implemented in silico for these simulations.

**Supplementary Fig. 9.**
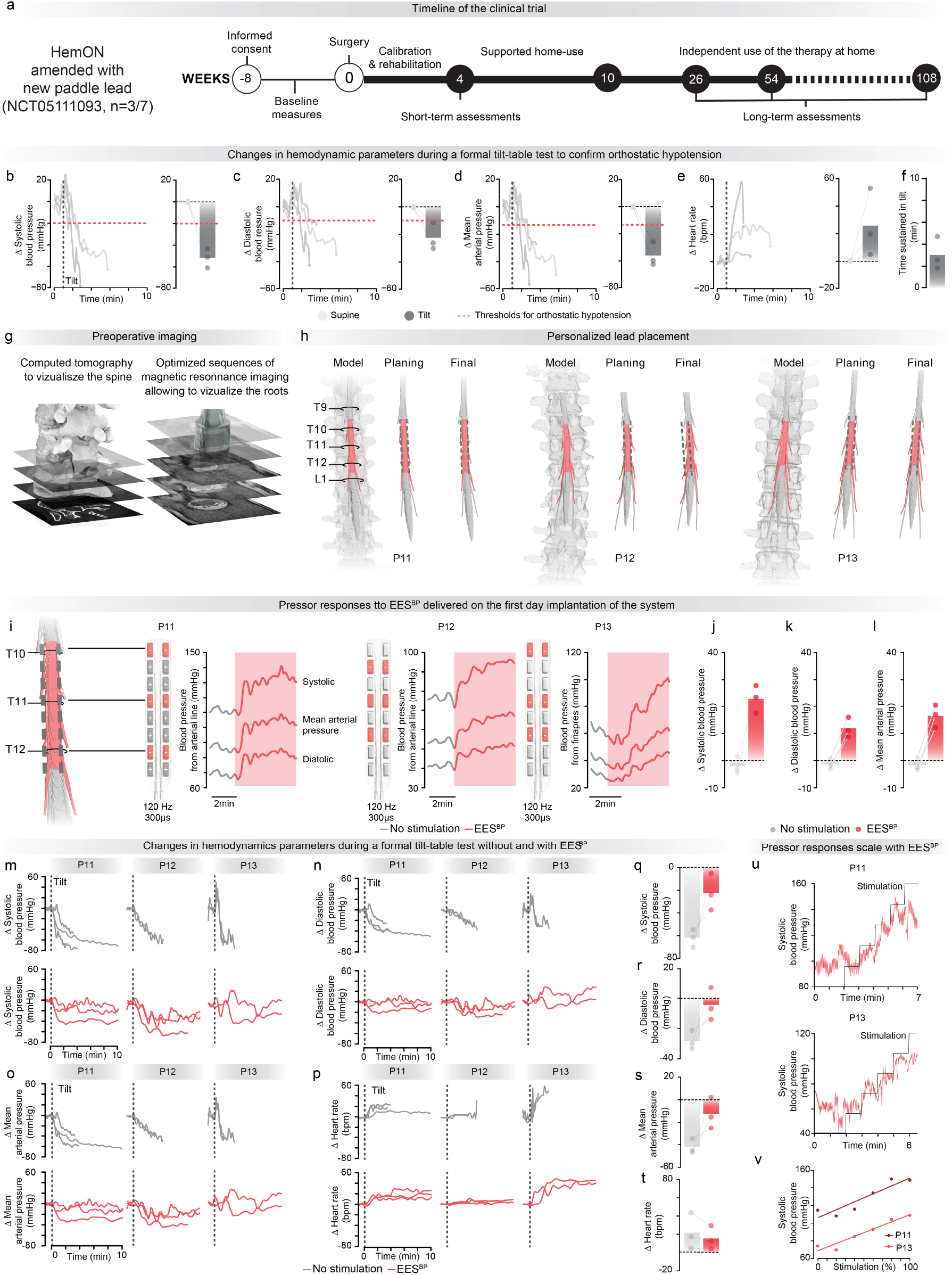
Validation of the complete purpose-built system to treat hemodynamic instability. **a**, Timeline of clinical trial 3 (HemON) amended for the new paddle lead conducted in Lausanne, Switzerland in 3 participants (Supplementary Table 2). **b**, Changes in systolic blood pressure during a formal 10-minute tilt-table test to verify that the four participants met the criteria for confirmed orthostatic hypotension (red dotted line). The bar graph reports the average drop in systolic blood pressure at or within (if the test had to be aborted before) 3 minutes of verticalization at a 70-degree tilt position. **c**, As in **b**, for diastolic blood pressure. **d**, As in **b**, for mean arterial pressure. **e**, As in **b**, for heart rate. **f**, Bar graph reporting the average duration in tilt shown in panels **b-d**. Each point represents a single tilt table test of a single participant (n = 3). **g**, Computed tomography and structural magnetic resonance imaging are acquired preoperatively to elaborate personalized models of the spine for each participant. **h**, Anatomical model, preoperative planning, and postoperative reconstruction of the final placement of the paddle lead for each participant (P11, P12, P13). **i**, Pressor responses elicited on the first day after the surgical implantation of the system when delivering EES^BP^ in a supine position (P11 - arterial line, P12 - arterial line, P13 - Finapres). **j**, Bar graph reporting the average change in systolic blood pressure without and with EES^BP^. **k**, As in **j**, for diastolic blood pressure. **l**, As in **j**, for the mean arterial pressure. **m**, Change in systolic blood pressure during a formal 10-minute tilt-table test without and with EES^BP^. Each line corresponds to a single test. **n**, As in **m**, for diastolic blood pressure. **o**, As in **m**, for mean arterial pressure. **p**, As in **m**, for heart rate. **q**, Bar graph reporting the average change in systolic blood pressure without and with EES^BP^ at or within (if the test had to be aborted before) 3 minutes of verticalization at a 70-degree tilt position. **r**, As in **q**, for diastolic blood pressure. **s**, As in **q**, for mean arterial pressure. **t**, As in **q**, for heart rate. **u**, Changes in systolic blood pressure with increases of EES^BP^ of 1 mA every 1 minute. Data are shown for P11 and P13. These recordings could not be collected in P12. **v**, Relationship between the amplitude of EES^BP^ and the modulation of systolic blood pressure for P11 and P13.

**Supplementary Fig. 10.**
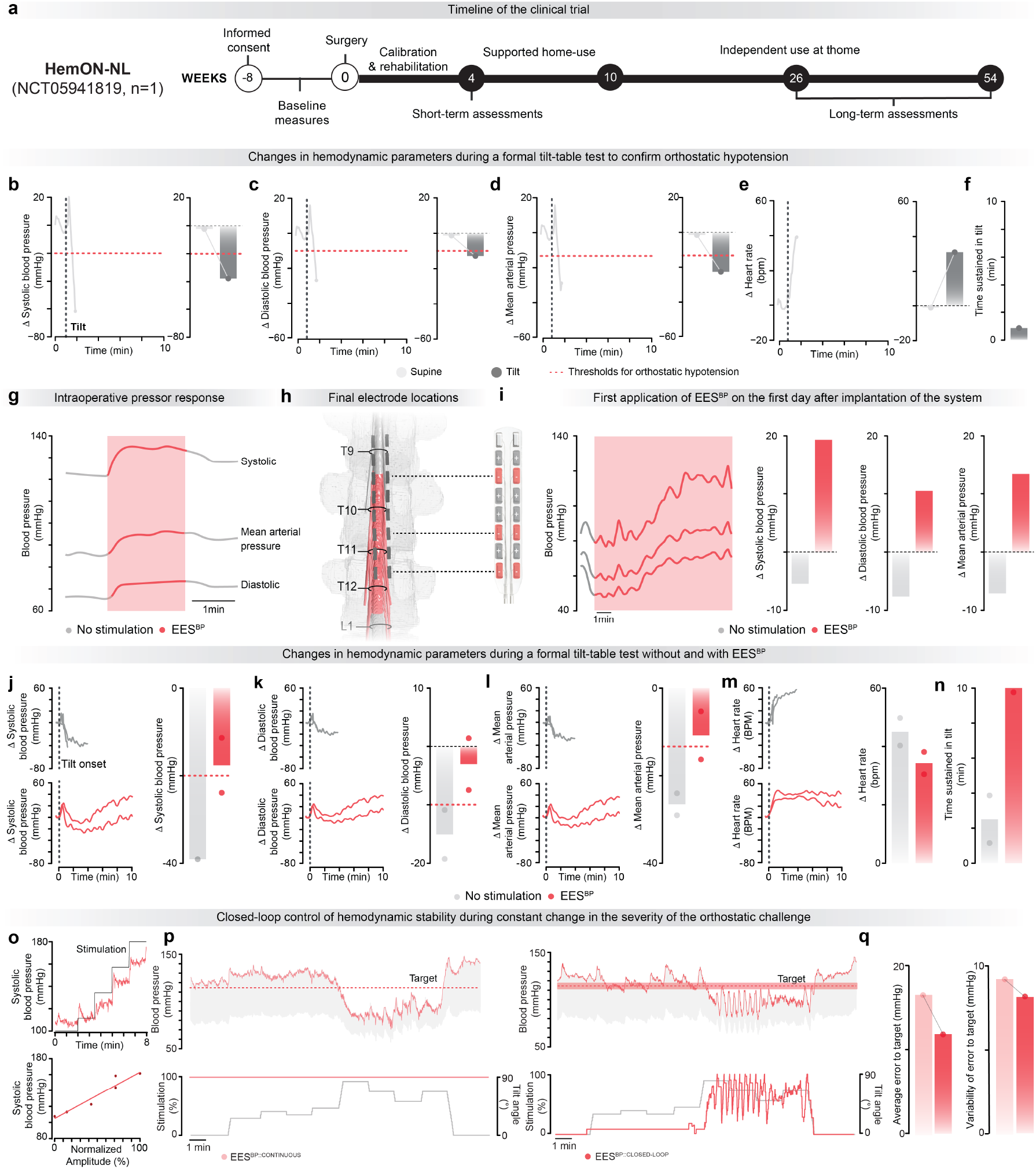
Independent validation of the complete system. **a**, Timeline of clinical trial 4 (HemON-NL) clinical trial conducted in Nijmegen, Netherlands. **b**, Changes in systolic blood pressure during a formal 10-minute tilt-table test to verify that the four participants met the criteria for confirmed orthostatic hypotension (red dotted line). The bar graph reports the average drop in systolic blood pressure at or within (if the test had to be aborted before) 3 minutes of verticalization at a 70-degree tilt position. **c**, As in **b**, for diastolic blood pressure. **d**, As in **b**, for mean arterial pressure. **e**, As in **b**, for heart rate. **f**, Bar graph reporting the average duration in tilt shown in panels **b-d. g**, Pressor response recorded intraoperatively using an arterial line. **h**, Final placement of the paddle lead represented into the personalized anatomical model of the spinal cord. **i**, Pressor responses recorded from a seated position during the first application of EES^BP^ after implantation of the system. The scheme of the paddle lead indicates the configuration of the electrodes, which was determined based on the location of the electrodes with respect to the targeted dorsal root entry zones. Bar graphs report the change in systolic blood pressure, diastolic blood pressure, and mean arterial pressure quantified 2 minutes after the application of EES^BP^. **j**, Change in systolic blood pressure without and with EES^BP^ during a formal 10-minute tilt-table test. Each line corresponds to a single test. The bar graph reports the average drop in systolic blood pressure at or within (if the test had to be aborted before) 3 minutes of verticalization at a 70-degree tilt position. **k**, As in **j**, for diastolic blood pressure. **l**, As in **j**, for mean arterial pressure. **m**, As in **j**, for heart rate. **n**, Bar graph reporting the average duration in tilt table test shown in panels **j-m. o**, Changes in systolic blood pressure with increases of EES^BP^ of 1 mA every 1 minute, and relationship between the amplitude of EES^BP^ and the modulation of systolic blood pressure (*R*^*2*^ = 0.96, p-value = 1.90e-02). **p**, Changes in diastolic and systolic blood pressure during a dynamic orthostatic challenge while EES^BP^ is applied continuously (*left*) or in closed-loop (*right*). **q**, Bar plots reporting the average error to target (*left*) and variability of the error to target (*right*) while EES^BP^ is applied continuously (*left*) or in closed-loop (*right*).

**Supplementary Fig. 11.**
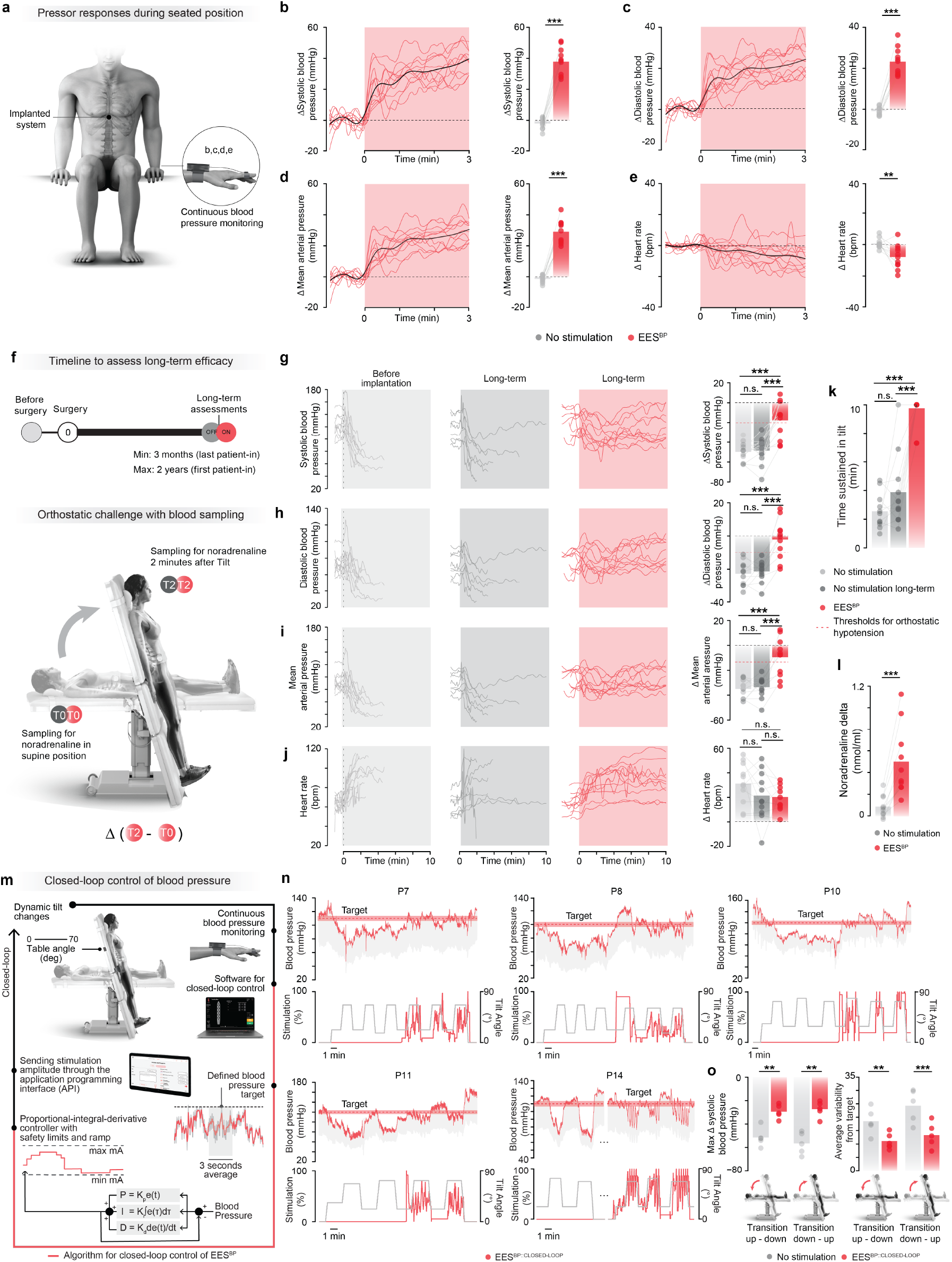
Long term efficacy of the system to regulate hemodynamic instability. **a**, Evaluation of pressor responses from a seated position in a wheelchair. **b**, Changes in systolic blood pressure when delivering EES^BP^ from a seated position (**a**). Each line corresponds to a response obtained in one of the 11 participants using the default EES^BP^ program after 3 months (n = 2) or at least 6 months after implantation of the system. The black line corresponds to the average response for all participants. The bar plots report the average change in systolic blood pressure in seated position before and 2 minutes after the onset of EES^BP^ (n = 11/11 - Trial 1, 3, 4; n = 9 measured at least 6 months after implantation, n = 2 measured with at least 3 months after implantation, paired, t-test; t = 16.63; p = 1.29e-08). **c**, As in **b**, for diastolic blood pressure (paired, t-test; t = 14.01; p = 6.75e-08). **d**, As in **b**, for mean arterial pressure (paired, t-test; t = 15.77; p = 2.16e-08). **e**, As in **b**, for heart rate (paired, t-test; t = 3.39; p = 6.9e-03). **f**, Timeline for long-term efficacy assessments, and time windows during which blood samples were taken during a formal 10-minute tilt-table test. **g**, Change in systolic blood pressure during a formal 10-minute tilt-table test conducted before implantation of the system (*left*), at least 3 months after implantation of the system but with EES^BP^ turned off (*middle*), and at the same time-point with EES^BP^ turned on (*right*) (n = 11/11 - Trial 1, 3, 4; participants of STIMO-HEMO, HemON and HemON-NL; n = 7 measured at least 6 months after implantation; n = 4 measured at least 3 months after implantation). Bar graph reports changes in systolic blood pressure during the formal tilt-table test. Each dot corresponds to a single test (n = 11/11 - Trial 1, 3, 4; repeated measures ANOVA with Tukey’s HSD). **h**, As in **g**,, for diastolic blood pressure (n = 11; repeated measures ANOVA with Tukey’s HSD). **i**, As in **g**,, for mean arterial pressure (n = 11; repeated measures ANOVA with Tukey’s HSD). **j**, As in **g**, for heart rate (n = 11; repeated measures ANOVA with Tukey’s HSD). **k**, Bar plots reporting the average duration in tilt-table test shown in panels g-j. Each dot corresponds to a single test per participant. **l**, Bar plots reporting changes in the concentration of endogenous norepinephrine in the blood between the supine position (T0) and after 2 minutes of verticalization at 70 deg (T2) without and with EES^BP^ (n = 10/13 - Trials 1,2 and 3; 3 participants could not be evaluated as described in methods, paired samples t-test, t = 5.49, p-value = 3.90e-04). **m**, Schematic of the experimental setup and algorithmic framework to control EES^BP^ in closed-loop in order to maintain systolic blood pressure within a user-defined target range. **n**, Changes in diastolic and systolic blood pressure during repeated transitions from tilts maintained at 20 or 70 degrees without EES^BP^ and with closed-loop control of EES^BP^. **o**, Bar plots reporting the average drop in systolic blood pressure during the tilts (n = 5, paired samples t-test, t = 5.78, p = 4.44e-03 average maximum drop during tilt down; t = 4.81, p = 8.57e-03 average maximum drop during tilt up), and average variability of error to target without EES^BP^ and with closed-loop control of EES^BP^ (n = 5, paired samples t-test, t = 5.75, p = 4.55e-03 average variability during tilt down; t = 10.39, p = 4.84e-03 average variability during tilt up). Each dot corresponds to the average values for a complete sequence per participant.

**Supplementary Fig. 12.**
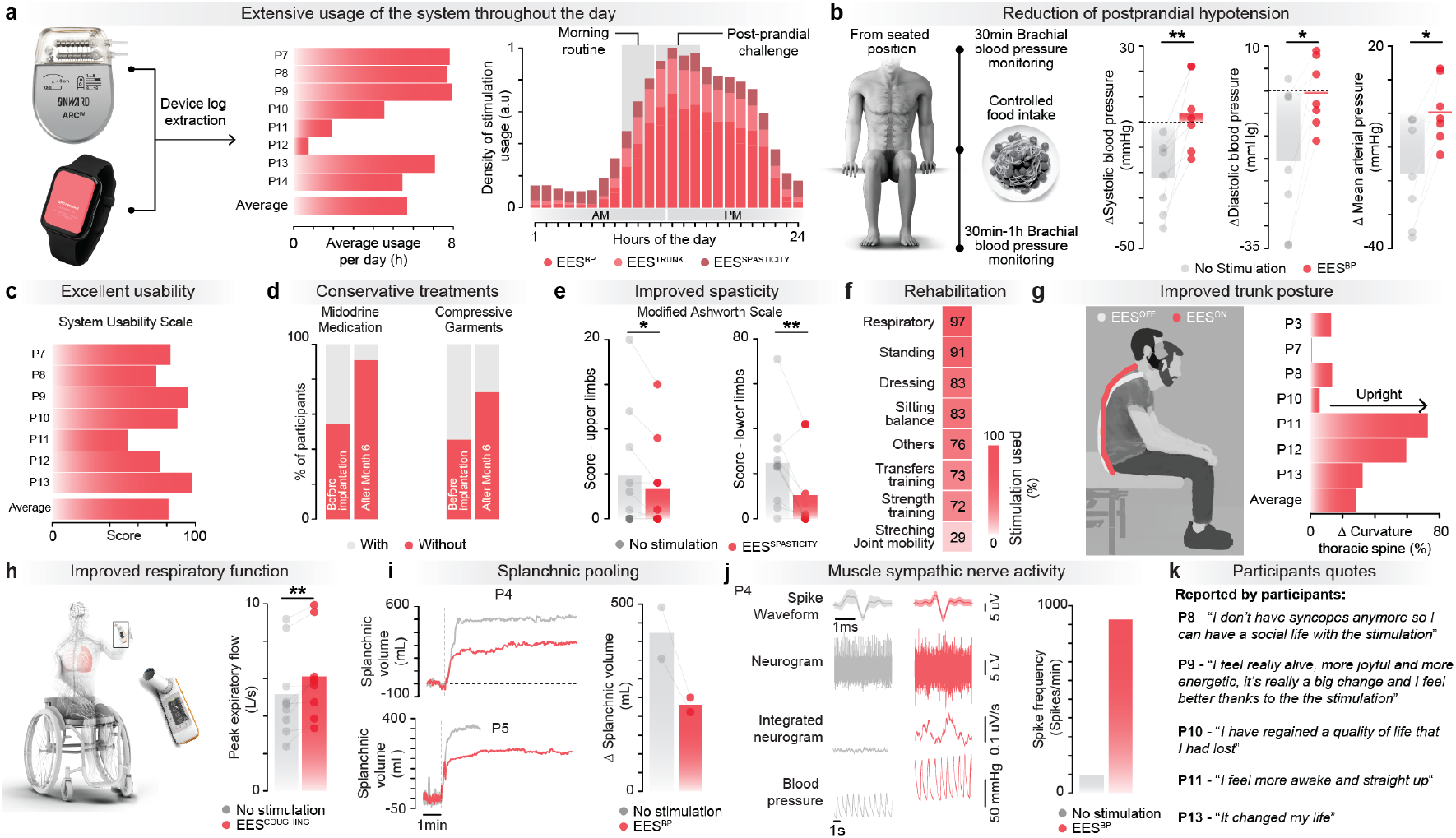
Long-term usability and efficacy of the system. **a**, Bar plots reporting the average daily usage of the system per participant (*left*), and the usage of the system throughout the hours of the day (*right*) for all participants implanted with at least part of the complete system (n = 8/8 - Trials 3-4). **b**, Bar plots reporting changes in hemodynamic parameters measured 30 minutes before and approximately every 5 minutes from 30 minutes to one hour after a controlled food intake blood pressure without and with EES^BP^ for participants presenting with postprandial hypotension (systolic, n = 7, paired samples t-test, t = 4.13, p-value = 6.12e-03; diastolic blood pressure, n = 7, paired samples t-test, t = 2.66, p-value = 3.78e-02; mean arterial blood pressure, n = 7, paired samples t-test, t = 3.37 p-value = 1.50e-02 - Trials 1-3). **c**, Bar plots reporting the score on the system usability scale during formal usability testing of the 7 participants implanted with at least the implanted pulse generator of the complete system (n = 7/7 - Trial 3). **d**, Bar plots reporting changes in the percentage of the 12 participants who took midodrine medication (*left*) and supportive garments (*right*) including compressive belts and/or compressive stockings, before implantation of the system and after six months of home use (*left*) (n = 12/14 - Trials 1-4, 2 participants in Trial 2 explanted prior to 6 months). **e**, Bar plots reporting spasticity score for the upper limb (*left*) and lower limb (*right*) without medication based on the Modified Ashworth Scale, quantified without and with programs to reduce muscle spasms, termed EES^SPASTICITY^ (n = 10/10 - Trials 1 and 3; left: paired samples t-test; t = 2.30; p-value = 4.8e-02, right: paired samples t-test; t = 3.30; p-value = 9.31e-03). **f**, Percentage of use of EES for various exercises measured over 1 month of rehabilitation for the seven participants who followed a personalized training program (n = 8/8 - Trials 3-4). **g**, Bar plot reporting changes in percentage of thoracic spine curvature without and with EES targeting the trunk musculature, termed EES^TRUNK^ (n = 7/10 - Trials 1 and 3, 3 participants not evaluated). **h**, Bar plot reporting the peak expiratory flow during a coughing maneuver without and with EES targeting trunk timed with coughing volition, termed EES^COUGHING^ (n = 9/10 - Trials 1 and 3, 1 incomplete assessment due to adverse event; paired samples t-test; t = 3.63; p-value = 6.70e-03). **i**, Changes in splanchnic volume without and without EES^BP^ (*left*), and bar plots reporting changes in splanchnic volume for 2 participants from the trial conducted in Calgary (2/3, Trial 2, 1 participant explanted prior to assessment). **j**, Muscle sympathetic nerve activity captured in the spike waveform, neurogram, integrated neurogram, and corresponding blood pressure without and with EES^BP^ for P4 (*left*). Bar plots reporting changes in neuronal spike frequency without and with EES^BP^ for P4 (*right*) (1/3, Trial 2, 2 participants explanted prior to assessment). **k**, Representative participant’s quotes extracted from semi-structured interviews expliciting changes in quality of life with the therapy.

**Supplementary Table 1.** Study information.

| Trial Number | Short Study Name | Study Title | Sponsor | Investigational Site | Manufacturer | Principal Investigator | Study Type | Primary Endpoint Description | ClinicalTrial.gov | Ethics Reference Number | Competent Authorities Number | National Registry Number | Number of Implanted Patients as of 31.01.2024 |
| --- | --- | --- | --- | --- | --- | --- | --- | --- | --- | --- | --- | --- | --- |
| 1 | STIMO-HEMO | Restoring Hemodynamic Stability Using Targeted Epidural Spinal Stimulation Following Spinal Cord Injury | CHUV | CHUV | MEDTRONIC | Jocelyne Bloch | Interventional, single group assignment | Investigate the preliminary safety of hemodynamic targeted epidural spinal stimulation (TESS) to modulate pressor responses and manage blood pressure instability in patients with chronic SCI located between C3 and T6 and who suffer from severe orthostatic hypotension (n=4). | NCT04994886 | CER-VD 2021-00588 | Swissmedic 10000882 | SNCTP000004406 | 3 |
| 2 | HEMO | Restoring Hemodynamic Stability Using Targeted Epidural Spinal Stimulation Following Spinal Cord Injury | University of Calgary | University of Calgary | MEDTRONIC | Aaron Phillips | Interventional, single group assignment | Investigate preliminary safety of hemodynamic targeted epidural spinal stimulation (TESS) to modulate pressor responses and manage blood pressure instability in patients with chronic SCI located between C3 and T6 who suffer from severe orthostatic hypotension (n=4) | NCT05044923 | REB21-0027 | not applicable | not applicable | 3 |
| 3 | HemON | Epidural Electrical Stimulation to Restore Hemodynamic Stability and Trunk Control in People With Spinal Cord Injury | EPFL | CHUV | ONWARD MEDTRONIC EPFL | Jocelyne Bloch | Interventional, single group assignment | Assess the safety of ARCIM Therapy at supporting the management of hemodynamic instability in participants with sub-acute or chronic spinal cord injury suffering from orthostatic hypotension. | NCT05111093 | CER-VD 2021-D0025 | Swissmedic 10000932 | SNCTP000004706 | 7 |
| 4 | HemON-NL | ARC Therapy to Restore Hemodynamic Stability and Trunk Control in People With Spinal Cord Injury | ONWARD | Sint Maartenskliniek | ONWARD | Ilse van Nes | Interventional, single group assignment | Assess the safety of ARC Therapy at supporting the management of hemodynamic instability in participants with sub-acute or chronic Spinal Cord Injury suffering from orthostatic hypotension. | NCT05941819 | 2023-16316 (METC) | NL83694.000.23 (CCMO) | not applicable | 1 |

**Supplementary Table 2.**
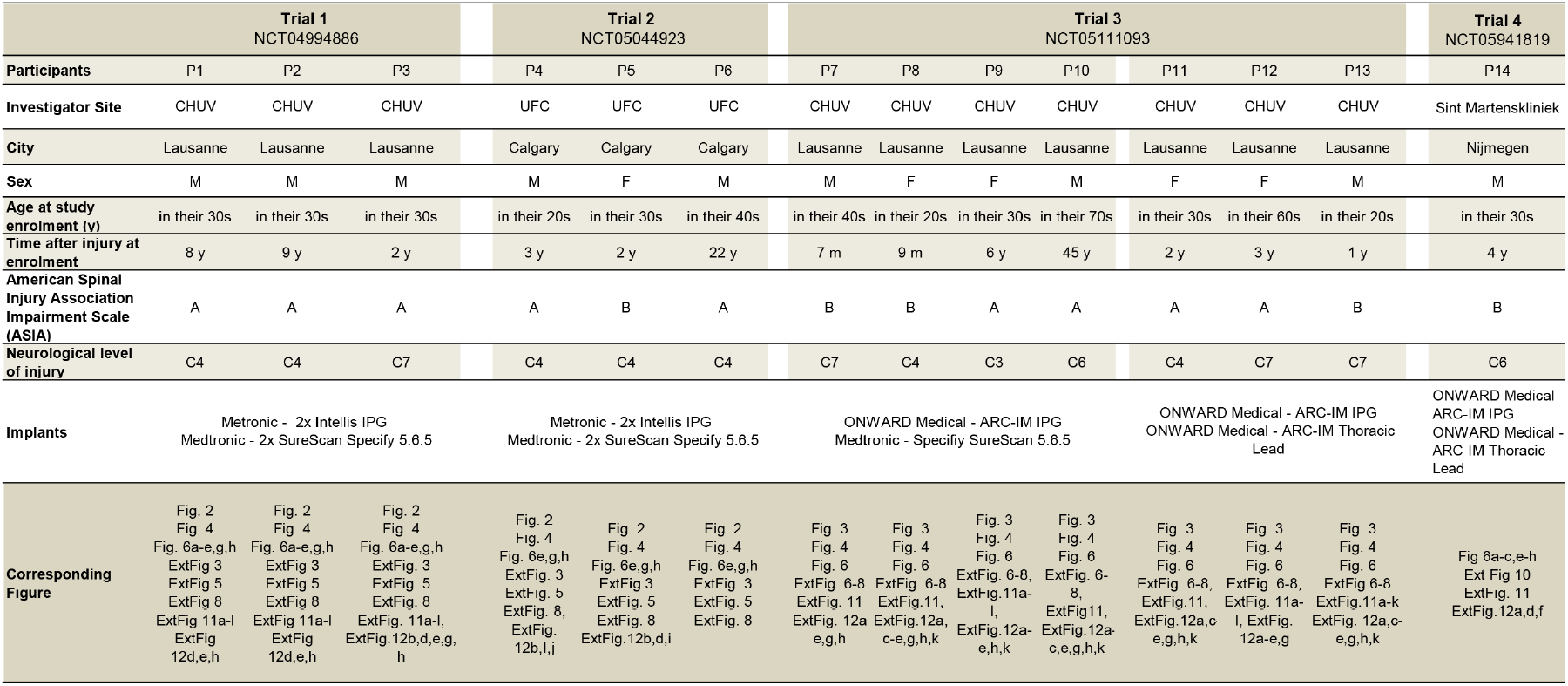
Patient Demographics. Demographic and neurological status of participants.

**Supplementary Table 3.** Validation cohort demographics. Participant demographics for validation cohort.

| Participant | Age at assessment (y) | Sex | Neurological Level of Injury | American Spinal Injury Association Impairment Scale (ASIA) | Time after injury at assessment (y) |
| --- | --- | --- | --- | --- | --- |
| VC01 | in their 40s | M | C7/C8 | A | 10 |
| VC02 | in their 40s | F | T4 | B/C | 25 |
| VC03 | in their 60s | M | C3-C6 | A | 5 |
| VC04 | in their 40s | F | Low cervical | C | 23 |
| VC05 | in their 60s | M | C4 | A | 11 |
| VC06 | in their 30s | F | Low cervical | C | 11 |
| VC07 | in their 40s | F | Low cervical | A | 26 |
| VC08 | in their 40s | F | T6 | A | 8 |
| VC09 | in their 20s | M | C4 | A | 5 |
| VC10 | in their 20s | M | C6/C7 | A | 2 |
| VC11 | in their 40s | F | C7 | A | 3.5 |
| VC12 | in their 40s | M | C4 | A | 21 |
| VC13 | in their 30s | F | C4 | B | 14 |
| VC14 | in their 30s | M | Low cervical | A | 1 |
| VC15 | in their 30s | M | C4 | B | 0.8 |
| VC16 | in their 40s | M | Low cervical | B | 1.5 |
| VC17 | in their 60s | M | High cervical | B | 2.5 |

**Supplementary Table 4.** Patient-by-patient safety data. Serious adverse device effects reported in all the participants of HemON and HemON-NL clinical trials.

|  | STIMO-HEMO |  |  | HEMO |  |  | HemON |  |  |  |  |  |  | HemON-NL |
| --- | --- | --- | --- | --- | --- | --- | --- | --- | --- | --- | --- | --- | --- | --- |
|  | P1<br>20<br>Months | P2<br>25<br>Months | P3<br>14<br>Months | P4<br>7<br>Months | P5<br>13<br>Months | P6<br>7<br>Months | P7<br>22<br>Months | P8<br>18<br>Months | P9<br>18<br>Months | P10<br>15<br>Months | P11<br>10<br>Months | P12<br>10<br>Months | P13<br>3<br>Months | P14<br>5<br>Months |
| Number of active months |  |  |  |  |  |  |  |  |  |  |  |  |  |  |
| **Number of Serious Adverse Device Effect (SADE)** | **0** | **2** | **0** | **1** | **1** | **1** | **0** | **0** | **1** | **0** | **0** | **0** | **0** | **0** |
| *Related to device/related to procedure* | 0 | 0 | 0 | 1 | 0 | 1 | 0 | 0 | 0 | 0 | 0 | 0 | 0 | 0 |
| *Only related to study procedure* | 0 | 2 | 0 | 0 | 1 | 0 | 0 | 0 | 1 | 0 | 0 | 0 | 0 | 0 |
| **Number of non-serious adverse device effect (ADE)** | **8** | **3** | **2** | **1** | **0** | **0** | **1** | **1** | **4** | **0** | **2** | **2** | **3** | **2** |
| *Related to device/related to procedure* | 3 | 2 | 0 | 1 | 0 | 0 | 0 | 0 | 1 | 0 | 2 | 0 | 1 | 2 |
| *Only related to study procedure* | 5 | 1 | 2 | 0 | 0 | 0 | 1 | 1 | 3 | 0 | 0 | 2 | 2 | 0 |

**Adverse device Effect (ADE)** (ISO 14155: 3.1): Adverse event possibly, probably, or causally related to the use of an investigational medical device. This definition includes adverse events resulting from insufficient or inadequate instructions for use, deployment, implantation, installation, or operation, or any malfunction of the investigational medical device. This definition includes any event resulting from use error or from intentional misuse of the investigational medical device. This includes comparator if the comparator is a medical device.
**Serious Adverse Device Effect (SADE)** (ISO14155): Adverse device effect that has resulted in any of the consequences characteristic of a serious adverse event.
Adverse events not related to device or procedure are not reported here.
When the (S)ADE is related to device, it has always extended to associated study procedure in this case. However, an adverse event might be related to study procedure while not being related to the device.

**Supplementary Table 5.**
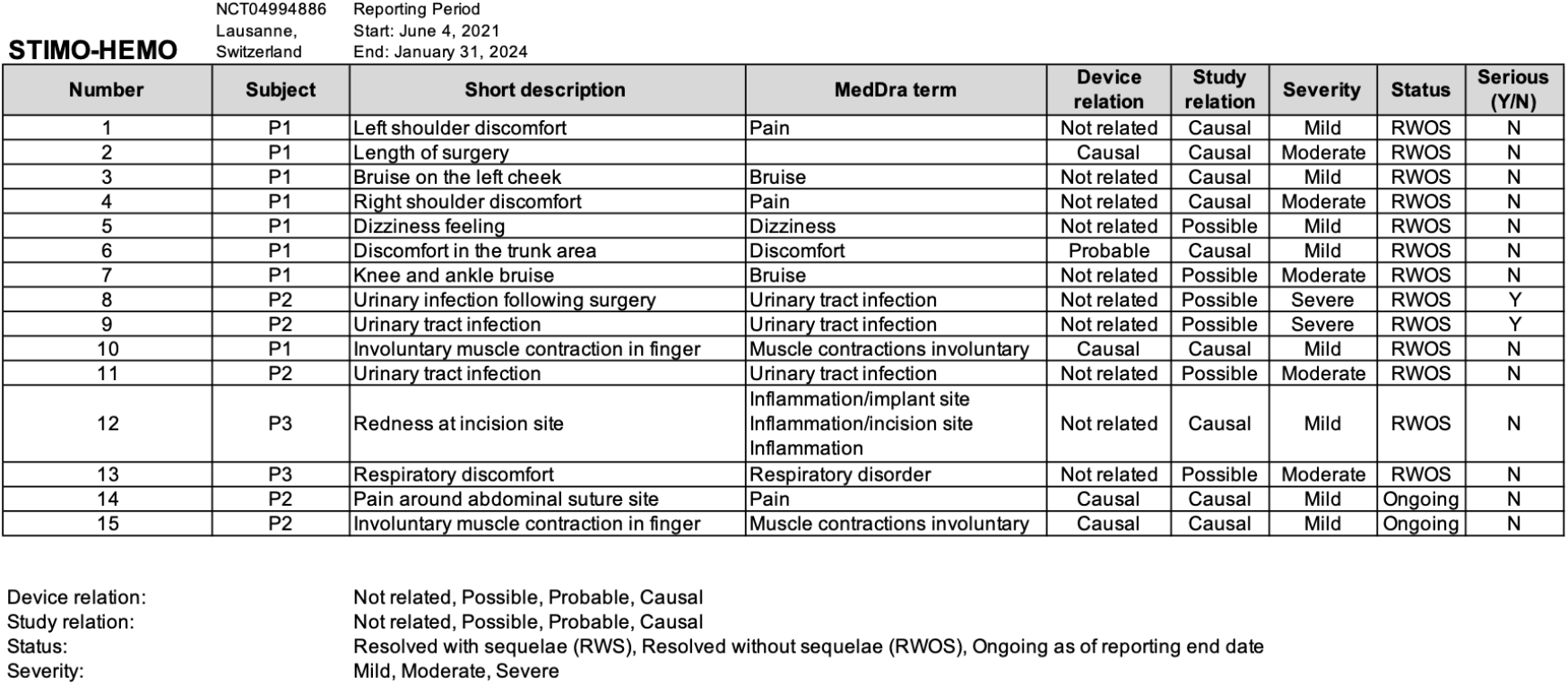
(Serious) Adverse Device Effects for STIMO-HEMO.

**Supplementary Table 6.** (Serious) Adverse Device Effects for HEMO.

| Number | Subject | Short description | MedDra term | Device relation | Study relation | Severity | Status | Serious (Y/N) |
| --- | --- | --- | --- | --- | --- | --- | --- | --- |
| 1 | P4 | Scar inflammation | Inflammation/incision site | Causal | Causal | Moderate | RWOS | N |
| 2 | P4 | Infection 4 months after surgery and device explant | Infection | Possible | Possible | Severe | RWOS | Y |
| 3 | P5 | Urinary tract infection following surgery | Urinary tract infection | Not related | Possible | Severe | RWOS | Y |
| 4 | P6 | Scar inflammation and systemic infection and device explant | Infection | Causal | Causal | Severe | RWOS | Y |

Device relation: Not related, Possible, Probable, Causal Study relation: Not related, Possible, Probable, Causal Status: Resolved with sequelae (RWS), Resolved without sequelae (RWOS), Ongoing as of reporting end date Severity: Mild, Moderate, Severe

**Supplementary Table 7.** (Serious) Adverse Device Effects for HemON and HemON-NL.

| Number | Subject | Short description | MedDra term | Device relation | Study relation | Severity | Status | Serious (Y/N) |
| --- | --- | --- | --- | --- | --- | --- | --- | --- |
| 1 | P8 | Neuropathic pain at puncture site during blood sampling, accompanied by insomnia | Pain | Not related | Possible | Mild | RWOS | N |
| 2 | P9 | Transient unexpected sensation due to overstimulation | Discomfort | Causal | Causal | Mild | RWOS | N |
| 3 | P9 | Blisters around stitches | Blister | Not related | Causal | Mild | RWOS | N |
| 4 | P9 | Urgency autonomic dysreflexia | Autonomic Dysreflexia | Not related | Probable | Severe | RWOS | Y |
| 5 | P9 | Supine hypertension after tilt and transient headache episode | Hypertension | Not related | Causal | Moderate | RWOS | N |
| 6 | P9 | Neuropathic pain at puncture site during blood sampling | Pain | Not related | Causal | Mild | RWOS | N |
| 7 | P7 | Skin irritation | Skin irritation | Not related | Causal | Mild | RWOS | N |
| 8 | P11 | Pain in abdominal region and constipation | Implant site pain | Possible | Probable | Moderate | RWOS | N |
| 9 | P12 | Malleolus fracture | Fracture | Not related | Probable | Moderate | RWOS | N |
| 10 | P12 | Bruises following surgery | Bruise | Not related | Causal | Mild | RWOS | N |
| 11 | P11 | High pressure accompanied by AD-like symptoms | Autonomic Dysreflexia | Causal | Causal | Mild | RWOS | N |
| 12 | P13 | Transient unexpected sensation due to stimulation | Discomfort | Causal | Causal | Mild | RWOS | N |
| 13 | P13 | Suture loosening | Suture rupture | Not related | Causal | Moderate | RWOS | N |
| 14 | P13 | Urinary Tract Infection accompanied by AD-like symptoms | Urinary Tract Infection/Autonomic Dysreflexia | Not related | Possible | Moderate | RWOS | N |

| Number | Subject | Short description | MedDra term | Device relation | Study relation | Severity | Status | Serious (Y/N) |
| --- | --- | --- | --- | --- | --- | --- | --- | --- |
| 1 | P14 | Fever | Fever | Possible | Possible | Mild | RWOS | N |
| 2 | P14 | Temperature elevation | Body temperature increased | Probable | Probable | Mild | RWOS | N |

Device relation: Not related, Possible, Probable, Causal Study relation: Not related, Possible, Probable, Causal Status: Resolved with sequelae (RWS), Resolved without sequelae (RWOS), Ongoing as of reporting end date Severity: Mild, Moderate, Severe

**Supplementary Note 1: Extended epidemiological results related to orthostatic hypotension in people with SCI** EES is a promising therapy to address the lack of satisfying management options for hemodynamic instability due to SCI. However, EES requires a neurosurgical intervention that must be weighed against the risks and benefits of the procedure. Establishing this balance requires an understanding of the prevalence, symptomatology, and effectiveness of current management strategies. While hypotension is a recognized medically refractory complication of SCI^2^, these epidemiological factors have not been quantified conclusively. To address this knowledge gap, we analyzed the Spinal Cord Injury Community Survey^26,27^ (SCICS), which includes self-reported information on symptoms of orthostatic hypotension and demographic information in 1,479 individuals living with chronic SCI (**Supplementary Fig. 1a**). This analysis revealed that 78% of individuals with tetraplegia had been told by a medical practitioner that they have orthostatic hypotension. Of this subset of individuals who were diagnosed, 28% of them were being treated for orthostatic hypotension, yet 91% still experienced symptoms (**Fig. 1a**). Orthostatic hypotension was still present in half the people with paraplegia (**Supplementary Fig. 1b**), and analysis of only individuals with complete (ASIA A) injuries led to comparable results (**Supplementary Fig. 1c**). Together these results suggested that conservative treatments for orthostatic hypotension are insufficient to manage the ongoing symptomatology. This analysis compelled us to characterize the pattern of hypotension-related symptoms due to SCI. For this purpose, we surveyed a diverse sample of 254 individuals with SCI from 26 different countries, which included responses to the Autonomic Dysfunction Following Spinal Cord Injury (ADFSCI) rating scale. When considering individuals with tetraplegia, we found that every individual who responded to the survey experienced symptoms of hypotension throughout the day. These symptoms were primarily characterized by lightheadedness, dizziness, fatigue, blurred vision, and weakness throughout the day (**Fig. 1b**). As expected, the frequency and severity of hypotension-related symptoms were higher in people with tetraplegia compared to people with paraplegia, and in those with complete SCI compared to those with incomplete SCI (**Supplementary Fig. 1e**). To link these outcomes to objective physiological quantifications, we conducted a formal 10-minute tilt table test in a subset of these individuals (**Fig. 1c, Supplementary Fig. 2a-d** and **Supplementary Table 3**). We found that individuals who met the criteria for orthostatic hypotension during the tilt-table test reported significantly more hypotensive symptoms (**Fig. 1d-f** and **Supplementary Fig. 2e-i**). These results established that i) the vast majority of individuals living with tetraplegia experience severe and persistent symptoms of hypotension; ii) hypotension is accompanied by quantifiable symptomatology that increases with the level and completeness of injury, iii) the ADFSCI is a valid self-reported measure of hypotension, and iv) the management of hypotension is insufficient to reduce the symptomatology. We concluded that these outcomes, combined with the long-term consequences of medically refractory hypotension, justified the consideration of therapeutic strategies involving surgical interventions to manage hypotension due to SCI.

**Supplementary Note 2: Requirements for a system to improve hemodynamic instability after SCI** Having identified the optimal location over which EES must be delivered to regulate blood pressure, we next sought to develop an implanted system that leveraged this understanding with the goal of establishing a therapy to improve the management of hemodynamic instability in people with SCI. We reasoned that such a system must combine the following features. First, the implantable neurostimulation platform should include the capacity to adjust EES in closed-loop. Indeed, while continuous EES is sufficient to stabilize blood pressure in static conditions, stabilization of blood pressure during dynamic changes in body orientation requires constant adjustment of EES amplitudes^1^. Moreover, constant adjustment of EES would be ideal in acute care settings wherein blood pressure lability impacts the neurological recovery of patients^5–8^. Second, the paddle lead must include an optimal configuration of electrodes to target the ensemble of dorsal root entry zones innervating the hemodynamic hotspot, while avoiding the undesired recruitment of sensory axons ascending in the dorsal columns. Since functional and anatomical experiments in mice, rats, nonhuman primates, and six humans revealed that the correct location to deliver EES is centered over the last three thoracic segments of the spinal cord, we concluded that the paddle lead must cover the entire extent of this hemodynamic hotspot. Third, the hardware and software components of the system must incorporate practical interfaces to support the rapid configuration of EES by physicians, and enable patients to operate the therapy safely and independently despite accessibility limitations. We aimed to develop a system that fulfilled these requirements and to establish the safety and efficacy of each component embedded in this new purpose-built implantable system through a series of stepwise clinical validations.

## References

1. Squair, J. W. et al. Neuroprosthetic baroreflex controls haemodynamics after spinal cord injury. Nature 590, 308–314. ISSN: 0028-0836 (2021).

2. Phillips, A. A. & Krassioukov, A. V. Contemporary Cardiovascular Concerns after Spinal Cord Injury: Mechanisms, Maladaptations, and Management. Journal of Neurotrauma 32, 1927–1942. ISSN: 0897-7151. 10.1089/neu.2015.3903 (2015).

3. Squair, J. W., Phillips, A. A., Harmon, M. & Krassioukov, A. V. Emergency management of autonomic dysreflexia with neurologic complications. Canadian Medical Association Journal 188, 1100–1103. ISSN: 0820-3946. 10.1503/cmaj.151311 (2016).

4. Wan, D. & Krassioukov, A. V. Life-threatening outcomes associated with autonomic dysreflexia: a clinical review. The Journal of Spinal Cord Medicine 37, 2–10. ISSN: 1079-0268. 10.1179/2045772313Y.0000000098 (2014).

5. Squair, J. W. et al. Empirical targets for acute hemodynamic management of individuals with spinal cord injury. Neurology 93, e1205–e1211. ISSN: 0028-3878. 10.1212/WNL.0000000000008125 (2019).

6. Squair, J. W. et al. Spinal cord perfusion pressure predicts neurologic recovery in acute spinal cord injury. Neurology 89, 1660–1667. ISSN: 0028-3878. 10.1212/WNL.0000000000004519 (2017).

7. Readdy, W. J. et al. Complications and outcomes of vasopres-sor usage in acute traumatic central cord syndrome. Journal of Neurosurgery. Spine 23, 574–580. ISSN: 1547-5654. 10.3171/2015.2.SPINE14746 (2015).

8. Inoue, T., Manley, G. T., Patel, N. & Whetstone, W. D. Medical and surgical management after spinal cord injury: vasopressor usage, early surgerys, and complications. Journal of Neurotrauma 31, 284–291. ISSN: 0897-7151. 10.1089/neu.2013.3061 (2014).

9. Wu, J.-C. et al. Increased risk of stroke after spinal cord in-jury: a nationwide 4-year follow-up cohort study. Neurology 78, 1051–1057. ISSN: 0028-3878. 10.1212/WNL.0b013e31824e8eaa (2012).

10. Cragg, J. J., Noonan, V. K., Krassioukov, A. & Borisoff, J. Cardiovascular disease and spinal cord injury: results from a national population health survey. Neurology 81, 723–728. ISSN: 0028-3878. 10.1212/WNL.0b013e3182a1aa68 (2013).

11. Illman, A., Stiller, K. & Williams, M. The prevalence of orthostatic hypotension during physiotherapy treatment in patients with an acute spinal cord injury. Spinal Cord 38, 741–747. ISSN: 1362-4393. 10.1038/sj.sc.3101089 (2000).

12. Carlozzi, N. E. et al. Impact of blood pressure dysregulation on health-related quality of life in persons with spinal cord injury: development of a conceptual model. Archives of Physical Medicine and Rehabilitation 94, 1721–1730. ISSN: 0003-9993. 10.1016/j.apmr.2013.02.024 (2013).

13. Rawlings, A. M. et al. Association of orthostatic hypotension with incident dementia, stroke, and cognitive decline. Neurology 91, e759–e768. ISSN: 0028-3878. 10.1212/WNL.0000000000006027 (2018).

14. National Institute of Biomedical Imaging and Bioengineering Consortium. Framework for a Research Study on Epidural Spinal Stimulation to Improve Bladder, Bowel, and Sexual Function in Individuals with Spinal Cord Injuries in (June 2015).

15. Pettigrew, R. I. et al. Epidural Spinal Stimulation to Improve Bladder, Bowel, and Sexual Function in Individuals With Spinal Cord Injuries: A Framework for Clinical Research. IEEE TRANSACTIONS ON BIOMEDICAL ENGINEERING 64, 253. https://www.nscisc.uab. (2 2017).

16. West, C. R. et al. Association of epidural stimulation with car-diovascular function in an individual with spinal cord injury. JAMA neurology 75, 630–632. ISSN: 2168-6149. 10.1001/jamaneurol.2017.5055 (2018).

17. Harkema, S. J. et al. Normalization of blood pressure with spinal cord epidural stimulation after severe spinal cord injury. Frontiers in Human Neuroscience 12, 83. ISSN: 1662-5161. 10.3389/fnhum.2018.00083 (2018).

18. Harkema, S. J. et al. Epidural spinal cord stimulation training and sustained recovery of cardiovascular function in individuals with chronic cervical spinal cord injury. JAMA neurology 75, 1569–1571. ISSN: 2168-6149. 10.1001/jamaneurol.2018.2617 (2018).

19. Darrow, D. et al. Epidural Spinal Cord Stimulation facilitates immediate restoration of dormant motor and autonomic supraspinal pathways after chronic neurologically complete spinal cord injury. English. Journal of neurotrauma 13, 158–167 (Jan. 2019).

20. Nightingale, T. E., Walter, M., Williams, A. M., Lam, T. & Krassioukov, A. V. Ergogenic effects of an epidural neuroprosthesis in one individual with spinal cord injury. Neurology 92, 338–340. ISSN: 0028-3878 (2019).

21. Aslan, S. C. et al. Epidural Spinal Cord Stimulation of Lumbosacral Networks Modulates Arterial Blood Pressure in Individuals With Spinal Cord Injury-Induced Cardiovascular Deficits. Frontiers in physiology 9, 565. ISSN: 1664-042X. 10.3389/fphys.2018.00565 (2018).

22. Strack, A. M., Sawyer, W. B., Marubio, L. M. & Loewy, A. D. Spinal origin of sympathetic preganglionic neurons in the rat. Brain Research 455, 187–191. ISSN: 0006-8993. 10.1016/0006-8993(88)90132-1 (1988).

23. Fink, G. D. & Osborn, J. W. Primer on the Autonomic Ner-vous System (Third Edition). Part III: Autonomic Physiology, 211–213 (2012).

24. Soriano, J. E. et al. Longitudinal interrogation of sympathetic neural circuits and hemodynamics in preclinical models. Nature Protocols, 1–34. ISSN: 1754-2189 (2022).

25. Soriano, J. E. et al. The neuronal architecture of autonomic dysreflexia. Under consideration.

26. Noreau, L. et al. Development and assessment of a community follow-up questionnaire for the Rick Hansen spinal cord injury registry. Archives of Physical Medicine and Rehabilitation 94, 1753–1765. ISSN: 0003-9993. 10.1016/j.apmr.2013.03.006 (2013).

27. Noreau, L., Noonan, V., Cobb, J., Leblond, J. & Dumont, F. Spinal Cord Injury Community Survey: A National, Comprehensive Study to Portray the Lives of Canadians with Spinal Cord Injury. Topics in Spinal Cord Injury Rehabilitation 20, 249–264. ISSN: 1082-0744 (2014).

28. Squair, J. W. et al. Implanted System for Orthostatic Hypotension in Multiple-System Atrophy. New England Journal of Medicine 386, 1339–1344. ISSN: 0028-4793 (2022).

29. Kaufmann, H. Consensus statement on the definition of orthostatic hypotension, pure autonomic failure and multiple system atrophy. Clinical Autonomic Research 6, 125–126. ISSN: 0959-9851. 10.1007/BF02291236 (1996).

30. Rowald, A. et al. Activity-dependent spinal cord neuromodulation rapidly restores trunk and leg motor functions after complete paralysis. Nature medicine 28, 260–271. ISSN: 1078-8956 (2022).

31. Joyner, M. J. & Casey, D. P. Regulation of Increased Blood Flow (Hyperemia) to Muscles During Exercise: A Hierarchy of Competing Physiological Needs. Physiological Reviews 95, 549–601. ISSN: 0031-9333 (2015).

32. Pickering, T. G., Harshfield, G. A., Kleinert, H. D., Blank, S. & Laragh, J. H. Blood Pressure During Normal Daily Activities, Sleep, and Exercise: Comparison of Values in Normal and Hypertensive Subjects. JAMA 247, 992–996. ISSN: 0098-7484 (1982).

33. Lavigne, G. & Kryger, M. Principles and Practice of Sleep Medicine (Sixth Edition). Part I: Principles of Sleep Medicine: Section 3Physiology in Sleep: Section 3: Physiology in Sleep, 115–117.e1 (2017).

34. Donaldson, H. H. & Davis, D. J. A description of charts showing the areas of the cross sections of the human spinal cord at the level of each spinal nerve. Journal of Comparative Neurology 13, 19–40. ISSN: 0092-7317 (1903).

35. Hynes, P. J., Fraher, J. P. & O’Sullivan, V. R. Segment length and root orientation in normal and sacrally attached spinal cords: A morphometric study. Clinical Anatomy 6, 167–172. ISSN: 0897-3806 (1993).

36. Ko, H.-Y., Park, J. H., Shin, Y. B. & Baek, S. Y. Gross quantitative measurements of spinal cord segments in human. Spinal Cord 42, 35–40. ISSN: 1362-4393. 10.1038/sj.sc.3101538 (2004).

37. Frostell, A., Hakim, R., Thelin, E. P., Mattsson, P. & Svensson, M. A Review of the Segmental Diameter of the Healthy Human Spinal Cord. Frontiers in Neurology 7, 238. ISSN: 1664-2295 (2016).

38. Mendez, A. et al. Segment-specific orientation of the dorsal and ventral roots for precise therapeutic targeting of human spinal cord. bioRxiv, 2020.01.31.928804 (2020).

39. Nunès, A. et al. Measurements and morphometric landmarks of the human spinal cord: A cadaveric study. Clinical Anatomy 36, 631–640. ISSN: 0897-3806 (2023).

40. Angeli, C. et al. Targeted Selection of Stimulation Parameters for Restoration of Motor and Autonomic Function in Individuals With Spinal Cord Injury. Neuromodulation: Technology at the Neural Interface. ISSN: 1094-7159 (2023).

41. Lewis, J. R. & Sauro, J. Item Benchmarks for the System Usability Scale. Journal of user experience 13, 158–167 (2018).

42. Vouga, T., Baud, R., Fasola, J., Bouri, M. & Bleuler, H. TWIICE-A Lightweight Lower-Limb Exoskeleton for Complete Paraplegics. 2017 International Conference on Rehabilitation Robotics (ICORR) 2017, 1639–1645 (2017).

43. Krassioukov, A. V., Johns, D. G. & Schramm, L. P. Sensitivity of sympathetically correlated spinal interneurons, renal sympathetic nerve activity, and arterial pressure to somatic and visceral stimuli after chronic spinal injury. Journal of Neurotrauma 19, 1521–1529. ISSN: 0897-7151. 10.1089/089771502762300193 (2002).

44. Skinnider, M. A. et al. Single-cell and spatial atlases of spinal cord injury in the Tabulae Paralytica. Nature 631, 150–163. ISSN: 1476-4687. 10.1038/s41586-024-07504-y (June 2024).

45. Rupp, R. et al. International Standards for Neurological Classification of Spinal Cord Injury. Topics in Spinal Cord Injury Rehabilitation 27, 1–22. ISSN: 1082-0744 (2021).

46. Isensee, F., Jaeger, P. F., Kohl, S. A. A., Petersen, J. & Maier-Hein, K. H. nnU-Net: a self-configuring method for deep learning-based biomedical image segmentation. Nature Methods 18, 203–211. ISSN: 1548-7091. eprint: 1904.08128 (2021).

47. Sekuboyina, A. et al. VerSe: A Vertebrae labelling and segmentation benchmark for multi-detector CT images. Medical Image Analysis 73, 102166. ISSN: 1361-8415. eprint: 2001. 09193 (2021).

48. Meng, D., Boyer, E. & Pujades, S. Vertebrae localization, segmentation and identification using a graph optimization and an anatomic consistency cycle. arXiv. eprint: 2110.12177 (2021).

49. Leener, B. D. et al. SCT: Spinal Cord Toolbox, an open-source software for processing spinal cord MRI data. NeuroImage 145, 24–43. ISSN: 1053-8119 (2017).

50. Bogert, L. W. J. & Lieshout, J. J. v. Non-invasive pulsatile arterial pressure and stroke volume changes from the human finger. Experimental Physiology 90, 437–446. ISSN: 1469-445X. 10.1113/expphysiol.2005.030262 (2005).

51. Jansen, J. R. et al. A comparison of cardiac output derived from the arterial pressure wave against thermodilution in cardiac surgery patients. British Journal of Anaesthesia 87, 212–222. ISSN: 0007-0912. 10.1093/bja/87.2.212 (2001).

52. Westerhof, B. E., Gisolf, J., Stok, W. J., Wesseling, K. H. & Karemaker, J. M. Time-domain cross-correlation barore-flex sensitivity: performance on the EUROBAVAR data set. Journal of Hypertension 22, 1371–1380. ISSN: 0263-6352. 10.1097/01.hjh.0000125439.28861.ed (2004).

53. Wieling, W., Ganzeboom, K. S. & Saul, J. P. Reflex syn-cope in children and adolescents. Heart 90, 1094–1100. ISSN: 1355-6037. 10.1136/hrt.2003.022996 (2004).

54. Whinnett, Z. I. et al. Multicenter randomized controlled crossover trial comparing hemodynamic optimization against echocardiographic optimization of av and VV delay of cardiac resynchronization therapy: the BRAVO trial. JACC. Cardiovascular Imaging 12, 1407–1416. ISSN: 1936-878X. 10.1016/j.jcmg.2018.02.014 (2019).

55. Ziegler, J. G. & Nichols, N. B. Optimum Settings for Automatic Controllers. Journal of Dynamic Systems, Measurement, and Control 115, 220–222. ISSN: 0022-0434 (1993).

56. Hasgall, P. et al. IT’IS Database for thermal and electromagnetic parameters of biological tissues Feb. 2022.

57. Rattay, F., Danner, S. M., Hofstoetter, U. S. & Minassian, K. Encyclopedia of Computational Neuroscience, 1–12 (2014).

58. Hines, M. L. & Carnevale, N. T. The NEURON simulation environment. Neural Computation 9, 1179–1209. ISSN: 0899-7667. 10.1162/neco.1997.9.6.1179 (1997).

59. Phillips, A. A., Krassioukov, A. V., Ainslie, P. N. & Warburton, D. E. R. Perturbed and spontaneous regional cerebral blood flow responses to changes in blood pressure after high-level spinal cord injury: the effect of midodrine. Journal of Applied Physiology 116, 645–653. ISSN: 8750-7587. 10.1152/japplphysiol.01090.2013 (2014).

60. Incognito, A. V. et al. Pharmacological assessment of the arterial baroreflex in a young healthy obese male with extremely low baseline muscle sympathetic nerve activity. Clinical Autonomic Research 28, 593–595. ISSN: 0959-9851. 10.1007/s10286-018-0559-2 (2018).

61. Incognito, A. V. et al. Evidence for differential control of muscle sympathetic single units during mild sympathoexcitation in young, healthy humans. American Journal of Physiology. Heart and Circulatory Physiology 316, H13–H23. ISSN: 0363-6135. 10.1152/ajpheart.00675.2018 (2019).

62. Yiallourou, T. I. et al. The effect of continuous positive airway pressure on total cerebral blood flow in healthy awake volunteers. Sleep and Breathing 17, 289–296. ISSN: 1520-9512 (2013).

63. Madgwick, S. O., Harrison, A. J. & Vaidyanathan, R. Estimation of IMU and MARG orientation using a gradient descent algorithm. 2011 IEEE International Conference on Rehabilitation Robotics 2011, 1–7 (2011).

64. Stewart, J. M., Medow, M. S., Glover, J. L. & Montgomery, L. D. Persistent splanchnic hyperemia during upright tilt in postural tachycardia syndrome. American Journal of Physiology. Heart and Circulatory Physiology 290, H665–73. ISSN:0363-6135. 10.1152/ajpheart.00784.2005 (2006).

65. Asboth, L. et al. Cortico-reticulo-spinal circuit reorganization enables functional recovery after severe spinal cord contusion. Nature Neuroscience 21, 576–588. ISSN: 1097-6256. 10.1038/s41593-018-0093-5 (2018).

